# MUTATE: A Human Genetic Atlas of Multi-organ AI Endophenotypes using GWAS Summary Statistics

**DOI:** 10.1101/2024.06.15.24308980

**Authors:** Aleix Boquet-Pujadas, Jian Zeng, Ye Ella Tian, Zhijian Yang, Li Shen, Andrew Zalesky, Christos Davatzikos, the MULTI consortium, Junhao Wen

## Abstract

Artificial intelligence (AI) has been increasingly integrated into imaging genetics to provide intermediate phenotypes (i.e., endophenotypes) that bridge the genetics and clinical manifestations of human disease. However, the genetic architecture of these AI endophenotypes remains largely unexplored in the context of human multi-organ system diseases. Using publicly available GWAS summary statistics from the UK Biobank, FinnGen, and the Psychiatric Genomics Consortium, we comprehensively depicted the genetic architecture of 2024 multi-organ AI endophenotypes (MAEs). We comparatively assessed the SNP-based heritability, polygenicity, and natural selection signatures of 2024 MAEs using methods commonly used in the field. Genetic correlation and Mendelian randomization analyses reveal both within-organ relationships and cross-organ interconnections. Bi-directional causal relationships were established between chronic human diseases and MAEs across multiple organ systems, including Alzheimer’s disease for the brain, diabetes for the metabolic system, asthma for the pulmonary system, and hypertension for the cardiovascular system. Finally, we derived polygenic risk scores for the 2024 MAEs for individuals not used to calculate MAEs and returned these to the UK Biobank. Our findings underscore the promise of the MAEs as new instruments to ameliorate overall human health. All results are encapsulated into the MUTATE genetic atlas and are publicly available at https://labs-laboratory.com/mutate.

## Introduction

Multi-organ research^1–9,10,11^ represents a pivotal frontier in advancing our understanding of human aging and disease. In particular, integrating artificial intelligence (AI) into multi-organ imaging genetics^1,4,12,3^ has emerged as a novel approach, offering potential promise in advancing precision medicine^13^. This integration introduces a new array of endophenotypes^14,15^, serving as intermediate, often quantitative, phenotypes, potentially reshaping how we perceive and approach medical AI^16^ in imaging and genetic research.

In recent years, three primary catalysts have significantly advanced the field of genetics. The first pivotal factor stems from the extensive collaborative efforts in consolidating large-scale multi-omics datasets, which has endowed researchers with unprecedented statistical power previously inaccessible. As an illustration, the UK Biobank (UKBB) study^17^ stands out for its comprehensive collection of multi-organ imaging^18^, genetics^19^, and proteomics^20,21^ data within the United Kingdom. Similarly, the FinnGen study^22^, conducted in Finland, has amassed extensive clinical and genetic data. Secondly, efforts toward open science have propelled the field, especially emphasizing the significance of publicly available resources, such as genome-wide association study (GWAS) summary statistics and widespread scientific dissemination. Notably, the FinnGen study and Psychiatric Genomics Consortium (PGC^23^) have publicly made all the GWAS summary statistics accessible^22^. Public GWAS platforms such as the GWAS Catalog^24^, OpenGWAS^25^, and GWAS ATLAS^26^ have consolidated and harmonized vast GWAS datasets, rendering them suitable for subsequent genetic analyses. Likewise, such good practice was also employed in the newly burgeoning field of brain imaging genetics^27^, including the BIG40 (https://open.win.ox.ac.uk/ukbiobank/big40/), the BIG-KP (https://bigkp.org/), BRIDGEPORT (https://labs-laboratory.com/bridgeport), and MEDICINE (https://labs-laboratory.com/medicine) knowledge portals. Finally, advanced computational genomics statistical methods using solely GWAS summary statistics, along with sufficient linkage disequilibrium information, have been developed, presenting an unparalleled chance to comprehend the genetic architecture of highly polygenic disease traits. For example, LDSC^28^ has been extensively utilized to estimate single-nucleotide polymorphism (SNP)-based heritability and genetic correlations. Mendelian randomization^29^ is a statistical method to dissect associations further, probing potential causal relationships among these complex human disease traits, although these methods often rely on several sensitive model assumptions^30^.

Despite these advancements, the intricate genetic foundation shaping these AI endophenotypes in the context of pleiotropic human disease endpoints (DE) within multi-organ systems remains largely uncharted. We previously applied AI to imaging genetic data and derived 2024 multi-organ AI endophenotypes (MAE). These encompassed 2003 multi-scale brain patterns of structural covariance (PSC) networks generated through a deep learning-analogy non-negative matrix factorization method^12^ (visualization for C32_1 encompassing deep subcortical structures: https://labs-laboratory.com/bridgeport/MuSIC/C32_1), 9 dimensional neuroimaging endophenotypes (DNE) quantifying neuroanatomical heterogeneity (also known as disease subtype) within 4 common brain diseases^4^, and 12 biological age gap (BAG) assessing the individual deviation in typical aging (i.e., acceleration or deceleration from the chronological age) across 9 human organ systems^1,3^ (**Supplementary eTable 1a**). The 2,024 MAEs were generated using data-driven AI and machine learning methods, distinguishing them from conventional imaging phenotypes (e.g., brain volume used in Elliott et al.^19^) and clinical diagnoses (e.g., binary Alzheimer’s disease (AD) diagnosis). For instance, we applied our Surreal-GAN model^31^ to neuroimaging data to derive two AD DNEs that capture the neuroanatomical heterogeneity of AD^32^. In contrast, traditional AD or AD-by-proxy approaches do not account for this heterogeneity and led to potential bias^33^, either inherently or in downstream GWAS analyses. To contribute to open science^34^, we made all the GWAS summary statistics derived from UKBB data publicly available at the MEDICINE knowledge portal: https://labs-laboratory.com/medicine. In addition, FinnGen analyzed genetic data for 2269 binary and 3 quantitative DEs from 377,277 individuals and 20,175,454 variants. They made these massive GWAS summary statistics publicly available to the community at https://finngen.gitbook.io/documentation/ (**Supplementary eTable 1b**). Finally, PGC consolidated GWAS results focused on neurological disorders worldwide and made the GWAS summary statistics accessible to the research community (https://pgc.unc.edu/, **Supplementary eTable 1c**) (**Fig. 1**).

**Figure 1:**
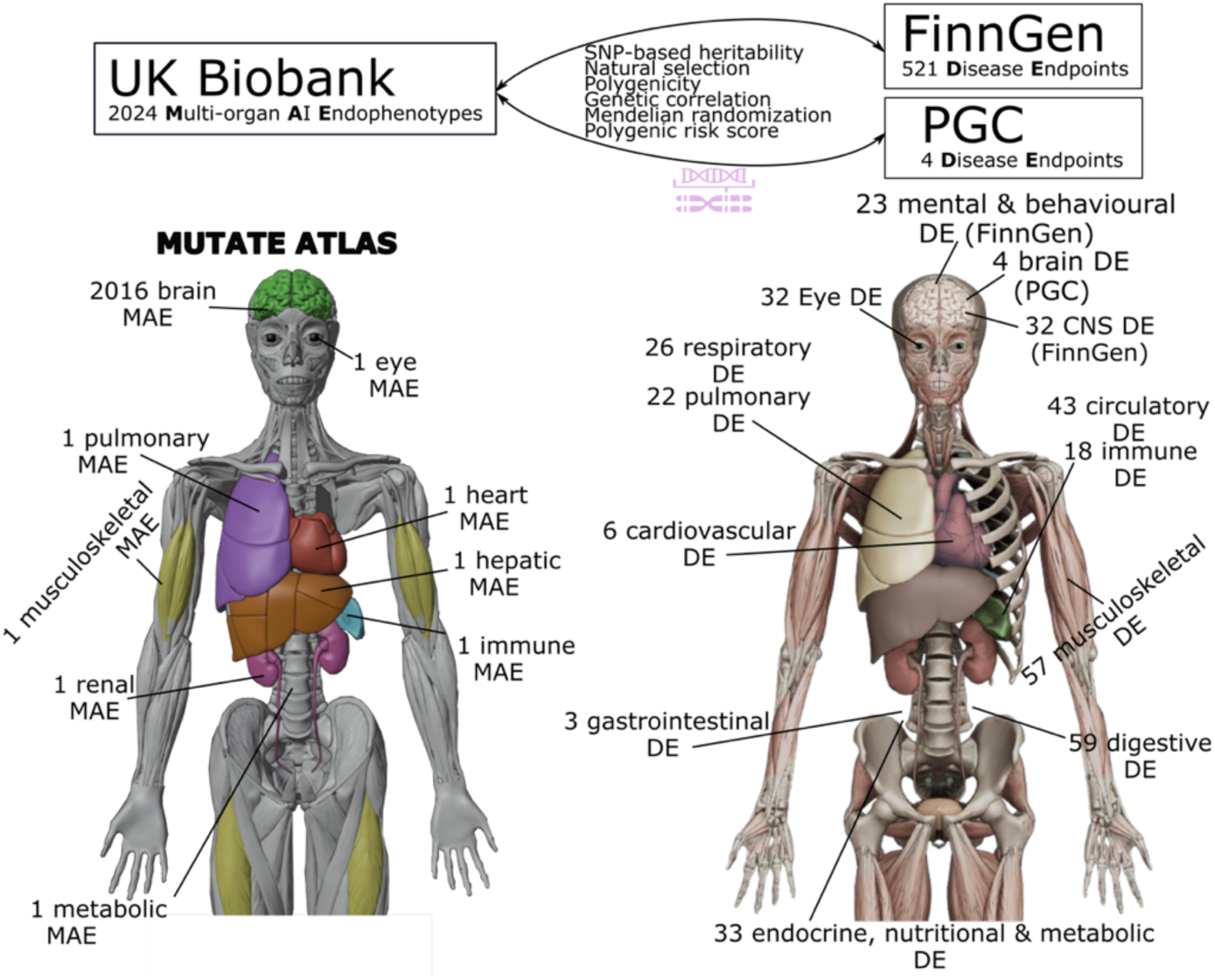
The 2024 MAEs and 525 Des. Using GWAS summary data from the UK Biobank, we generated 2024 MAEs (https://labs-laboratory.com/medicine/; https://labs-laboratory.com/bridgeport). Additionally, we harmonized GWAS summary data from FinnGen (https://www.finngen.fi/en/access_results) and PGC (https://pgc.unc.edu/for-researchers/download-results/) to incorporate 525 disease endpoints (DEs). All analyses in this study were conducted exclusively using GWAS summary data from these three public resources.

This study harnesses the extensive GWAS summary resources made publicly available by us on behalf of UKBB, FinnGen, and PGC (**Method 1**), along with the utilization of several advanced computational genomics statistical methods (refer to **Code Availability**), to thoroughly depict the genetic architecture of the 2024 MAEs (**Method 2**) and 525 DEs (>5000 cases) in the context of multi-organ investigations. Importantly, our previous research explored the genetic foundation of the 2024 MAEs but did not systematically encompass the FinnGen or PGC data. Specifically, we included 521 DEs released by the FinnGen study, accessible at https://finngen.gitbook.io/documentation/v/r9/, and 4 brain DEs (Alzheimer’s disease (AD), Attention-deficit/hyperactivity disorder (ADHD), bipolar disorder (BIP), and schizophrenia (SCZ)) from PGC (https://pgc.unc.edu/). This study expanded on this by systematically benchmarking the genetic analyses and comprehensively comparing various statistical methodologies^28,30,35–41^ (**Method 3**). Specifically, we aimed to compute the SNP-based heritability 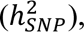 polygenicity (*π*), the relationship between SNP effect size and minor allele frequency (*S*: signature of natural selection, genetic correlation (*r*_*g*_), causality, and polygenic risk score (PRS) between the 2024 MAEs and 525 DEs. These findings were encapsulated within the MUTATE (**MU**l**T**i-organ **A**I endopheno**T**yp**E**) genetic atlas, which is publicly available at https://labs-laboratory.com/mutate.

## Materials and methods

### Method 1: The MULTI consortium

The `MULTI` consortium is an ongoing initiative to integrate and consolidate multi-organ and multi-omics biomedical data, including imaging, genetics, and proteomics. Building on existing consortia and studies, such as those listed below, `MULTI` aims to curate and harmonize the data to model human aging and disease across the lifespan at individual and summary levels. The present study solely utilized GWAS summary statistics; no individual-level data were used. We downloaded the GWAS summary statistics from three web portals for the 2024 MAEs, 521 DEs from FinnGen and 4 DEs from PGC.

### UKBB

UKBB is a population-based study of approximately 500,000 people recruited from the United Kingdom between 2006 and 2010. The UKBB study has ethical approval, and the ethics committee is detailed here: https://www.ukbiobank.ac.uk/learn-more-about-uk-biobank/governance/ethics-advisory-committee.

The GWAS summary statistics for all the 2024 MAEs are publicly available at the MEDICINE knowledge portal: https://labs-laboratory.com/medicine, which focuses on disseminating scientific findings on imaging genetics and AI methods in multi-organ science. Specifically, among the 2024 MAEs, 2003 PSCs – at varying scales from C32 to C1024 – were structural covariance networks derived via the sopNMF method^12^. 9 DNEs^4^ captured the neuroanatomical heterogeneity of four brain diseases (AD1-2, ASD1-3, LLD1-2, and SCZ1-2) using semi-supervised clustering or representation learning methods. 12 multi-organ BAGs (GM, WM, FC^3^, multimodal brain BAGs, cardiovascular BAG, eye BAG, hepatic BAG, immune BAG, musculoskeletal BAG, metabolic BAG, pulmonary BAG, and renal BAG^42^) were derived from various machine learning models to quantify the individual-level deviation from typical brain aging due to various pathological effects. Detailed AI methodologies are presented in **Method 2** for the MAEs, DNEs, and BAGs. All GWASs were performed within European ancestries and using the GRCh37 human genome assembly; the GWAS model (PLINK^43^ for linear model and fastGWA^44^ for linear mixed-effect model), sample sizes, and covariates included are detailed in the original papers and also in **Supplementary eTable 1a**.

### FinnGen

The FinnGen^22^ study is a research project based in Finland that explores combined genetics and health registry data to understand the underlying causes and mechanisms behind various disease endpoints. It particularly emphasizes the genetic basis of diseases in the Finnish population (>500,000) by conducting extensive GWAS and analyzing large-scale genomic data in collaboration with multiple research institutions and organizations. FinnGen has generously made their GWAS results publicly available to the community for research purposes (https://www.finngen.fi/en/access_results).

The present study used the GWAS summary statistics version R9 released to the public on May 11, 2022, after harmonization by the consortium. In the R9 release, FinnGen analyzed 2269 binary and 3 quantitative endpoints from 377,277 individuals and 20,175,454 variants. Regenie^45^ was used to run the GWAS models, including sex, age, 10 PCs, and genotyping batch as covariates. Genotype imputation was done with the population-specific SISu v4.0 reference panel. In our analysis, we concentrated solely on binary DEs with case numbers exceeding 5000 to ensure adequate statistical power, given the highly imbalanced case/control ratios. As the released data were based on the GRCh38 human genome assembly, we lifted the GWAS summary statistics to the GRCh37 version for all genetic analyses. **Supplementary eTable 1b** details the included 521 DEs. More details can be found at the FinnGen website: https://finngen.gitbook.io/documentation/v/r9/.

### Psychiatric Genomics Consortium

PGC^23^ is an international coalition of researchers exploring the genetic underpinnings of psychiatric disorders and beyond. This collaborative effort unites scientists globally to examine and decipher extensive genomic datasets concerning various brain diseases. The primary goal of PGC involves uncovering and comprehending the genetic elements that contribute to various psychiatric disorders, such as schizophrenia, bipolar disorder, and major depressive disorder. We downloaded GWAS summary statistics from the PGC website (https://pgc.unc.edu/for-researchers/download-results/) and manually harmonized the data to our Mendelian randomization analyses to replicate the FinnGen findings.

### Harmonization of the GWAS summary statistics from the 3 resources

Harmonization of GWAS summary statistics across different models and consortia for various software is crucial, such as aligning the effect allele and the direction of the effect size. There’s currently no established standard in the field for this process, although some advice has been proposed^46^. Certain software internally harmonizes data based on the allele frequency of the effect allele, such as the *TwoSampleMR* package^47^ for Mendelian randomization. We outlined the detailed procedures used to harmonize the GWAS summary data from the three sources.

For our UKBB MAE GWAS summary data, we harmonized the effect allele as the alternative allele from PLINK and A1 from fastGWA. Corresponding allele frequency was also needed for certain analyses, such as the *TwoSampleMR* package for Mendelian randomization. Due to privacy considerations, allele frequency information is not publicly available on the MEDICINE portal. However, it can be provided upon direct email request. P-value, effect sizes (e.g., BETA value and SE), and sample sizes are harmonized to respective to the effect allele. The variant identifier is based on the rs ID number, not the chromosome number and position number combination. All our GWAS summary data are on the human genome build assembly GRCh37.

The FinnGen team has systematically harmonized the GWAS summary data for the 521 DEs involved. The alternative allele serves as the effect allele. The rsID number represents the SNP; the chromosome number and position are also shared. The data includes P-values, effect sizes, and allele frequencies for both the alternative and reference alleles. Refer to the FinnGen documentation for more details: https://finngen.gitbook.io/documentation/data-description. We downloaded the original data and converted the data to GRCh37.

PGC did not systematically harmonize the GWAS summary statistics; the available data information and format depend on each study. **Supplementary eTable 1c** details the 4 DEs (AD, ADHD, bipolar disorder, and schizophrenia) included after the data filtering procedure. First, we ensured that the study population comprised individuals of European ancestry and, if necessary, lifted the data to GRCh37. Secondly, we excluded two studies where the allele frequency is unavailable because the *TwoSampleMR* package^47^ requires this information to harmonize the exposure and outcome data (e.g., flip the effect allele and effect size). Thirdly, we confirmed that the GWAS summary statistics didn’t overlap with UKBB data. Specifically, the AD GWAS summary data^48^ explicitly offered a version that excluded participants from UKBB. In addition, the original dataset lacked a column for the rsID number. To deal with this, we employed a mapping approach using the chromosome number and position to the dpSNP database (version 150), which allowed us to obtain the corresponding rsID numbers.

### Method 2: 2024 multi-organ AI endophenotypes

#### (a): The 2003 patterns of structural covariance of the brain

In our earlier study^12^, we utilized the sopNMF method on an extensive and varied brain imaging MRI dataset (*N*=50,699, including data from UKBB) to generate the multi-scale brain PSCs. The scale C ranges from 32 to 1024, progressively increasing by a factor of 2; 11 PSCs vanished during models.

Biologically, the 2003 PSCs represent data-driven structural networks that co-vary across brain regions and individuals in a coordinated fashion. Mathematically, the sopNMF method is a stochastic approximation ("deep learning-analogy") constructed and extended based on opNMF^49,50^. Consider an imaging dataset comprising *n* images, each containing *d* voxels. We represent the data as a matrix ***X***, where each column corresponds to a flattened image: 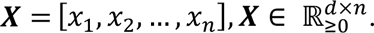 The method factorizes ***X*** into two low-rank matrices 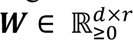 and 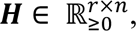 subject to two important constraints: *i*) non-negativity and *ii*) column-wise orthonormality. More mathematical details can be referred to the original references^12,49,50^ and **Supplementary eMethod 2a**.

#### (b): The 9 dimensional neuroimaging endophenotypes of the brain

The nine DNEs captured the neuroanatomical heterogeneity of four brain diseases, including AD1-2 for AD^32^, ASD1-3 for autism spectrum disorder^51^, LLD1-2 for late-life depression^52^, and SCZ1-2 for schizophrenia^53^. The underlying AI methodologies involved two different semi-supervised clustering or representation learning algorithms: Surreal-GAN^54^ and HYDRA^55^. Refer to a review for details of the semi-supervised learning^56^, which primarily seeks the so-called "*1- to-k*" mapping patterns or transformations from reference domains (like healthy controls) to target domains (such as patients).

Surreal-GAN^54^ was used to derive AD1-2^32^. It unravels the intrinsic heterogeneity associated with diseases through a deep representation learning approach. The methodological innovation, compared to its precentor Smile-GAN^57^, lies in how Surreal-GAN models disease heterogeneity: it interprets it as a continuous dimensional representation, ensures a consistent increase in disease severity within each dimension, and permits the simultaneous presence of multiple dimensions within the same participant without exclusivity. More mathematical details are presented in **Supplementary eMethod 2b.**

HYDRA^55^ was employed to derive the other 7 DNEs. It utilizes a widely adopted discriminative technique, namely support vector machines (SVM), to establish the "*1-to-k*" mapping. The model extends multiple linear SVMs to the nonlinear domain by piecing them together. This approach serves the dual purpose of classification and clustering simultaneously. Specifically, it creates a convex polytope by amalgamating hyperplanes derived from *k* linear SVMs. This polytope separates the healthy control group from the *k* subpopulations within the patient group. Conceptually, each face of this convex polytope can be likened to encoding each subtype (categorical trait) or dimension (continuous trait), capturing distinctive disease effects (Refer to **Supplementary eMethod 2c**).

#### (c): The 12 biological age gaps of nine human organ systems

The nine multi-organ BAGs (brain, cardiovascular, eye, hepatic, immune, musculoskeletal, metabolic, pulmonary, and renal) were derived from a previous study^2^ that used AI to predict the chronological age of healthy individuals without chronic medical conditions: AI-predicted age – chronological age. Using a 20-fold cross-validation procedure, we applied the model for each organ system, employing a linear support vector machine. Before training each model iteration, standardization was applied to measures (excluding categorical variables) within the training set. The model was solved using sequential minimal optimization with a gap tolerance of 0.001. The support vector regression settings were adjusted for optimization, adhering to established principles in the field^58^.

Alongside the nine organ BAGs, we previously derived three multimodal brain BAGs (GM, WM, and FC-IDP) using features from gray matter (GM), white matter (WM), and functional connectivity (FC) in MRI scans^3^. We systematically compared four machine learning models: SVR, LASSO regression, multilayer perceptron, and a five-layer neural network. We employed nested cross-validation (CV) and included an independent test dataset^59^ for a fair comparison across different models and MRI modalities. This process involved an outer loop CV with 100 repeated random splits: 80% for training and validation and 20% for testing. Within the inner loop, a 10-fold CV was utilized for hyperparameter tuning. Furthermore, we reserved an independent test dataset, which was kept unseen until the fine-tuning of the machine learning models^60^ (e.g., hyperparameters for SVR) was completed.

### Method 3: Genetic analyses based on GWAS summary statistics

#### (a): The genetic architecture of the 2024 MAEs and 525 DEs

Primarily, we used SBayesS^61^ to estimate three sets of parameters that fully unveil the genetic architecture of the 2024 MAEs and 525 DEs. SBayesS is an expanded approach capable of estimating three essential parameters characterizing the genetic architecture of complex traits through a Bayesian mixed linear model^62^. This method only requires GWAS summary statistics of the SNPs and LD information from a reference sample. These parameters include SNP-based heritability 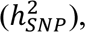 polygenicity (*π*), and the relationship between minor allele frequency (MAF) and effect size (*S*). We used the software pre-computed sparse LD correlation matrix derived from the European ancestry by Zeng et al.^61^. More mathematical details can be found in the original paper from Zeng et al.^61^. We ran the *gctb* command^62^ using the argument *--sbayes S*, and left all other arguments by default. When applying SBayesS to the 2025 MAEs and 525 DEs summary data, we found that 18 DEs failed to converge in the MCMC sampling, which may be due to LD differences between FinnGen and UKBB samples (the latter was used as the LD reference in SBayesS).

To benchmark different methods used in the field for SNP-based heritability estimates, we also employed two other methods based on GWAS summary data: *i*) LDSC^28^ and *ii*) SumHer^36^. LDSC relies on the principle that the correlation between SNP effect sizes and linkage disequilibrium with neighboring SNPs can be used to estimate the proportion of heritability explained by all SNPs using GWAS summary data. For LDSC, we used the precomputed LD scores from the 1000 Genomes of European ancestry. All other parameters were set to default in the software. After merging the GWAS summary statistics, we chose the 1000 Genomes reference panel for fair comparisons between the two studies and ensured that most SNPs were included in the analyses. For example, for the DE (RX_PARACETAMOL_NSAID), after merging with the reference panel LD, 1,171,361 remained. For the first MAE (C32_1), 1,092,510 SNPs remained after the same merging procedure. Furthermore, FinnGen didn’t provide the original genotype data; they only shared the LD information via the LDstore software but did not provide the allele information. Consequently, we cannot generate in-sample LD scores using the LDSC software. Finally, a prior investigation^63^ showcased the robustness of LDSC concerning the selection of LD reference panels – multi-ethnic European, Finnish-only, non-Finnish European from 1000 Genomes Phase 3 data, and FINRISK Finnish reference panel – regarding heritability estimates in four lipid traits within a Finnish population.

For SumHer, we used the BLD-LDAK model, as the software suggested. BLD-LDAK stands for "Bayesian LD-adjusted Kinship," where LD-adjusted kinship refers to the calculation of genetic relatedness between individuals using information about the correlation of alleles between nearby SNPs (linkage disequilibrium). We used the software-provided tagging file, generated from 2000 white British individuals, as a reference penal suggested by the software for European ancestry groups. The HapMap3 data (https://www.broadinstitute.org/medical-and-population-genetics/hapmap-3) merged with the tested GWAS summary SNPs. Similarly, we ensured sufficient SNPs remained after merging with the reference panel. All other parameters were set to default. SumHer differs from LDSC in several ways: *i*) it models inflation multiplicatively, whereas LDSC uses an additive approach; *ii*) it accounts for uneven LD patterns and incorporates MAF on SNP effect; and *iii*) it utilizes a restricted maximum likelihood solver rather than regression to estimate the 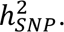

#### (b): Genetic correlation

We used three different methods to compute the MAE-DE pairwise (*N*=2024×525=1,062,600) genetic correlations (*r*_g_): *i*) LDSC^28^, *ii*) GNOVA^37^, and *iii*) HDL^41^.

An earlier study^65^ highlighted the significance of selecting an appropriate LD score reference panel for genetic correlation estimates based on summary statistics. We generated the same reference panel for LD scores across the three software for a fair comparison. For LDSC, we used the precomputed LD scores from the 1000 Genomes of European ancestry provided by the software. All other parameters were set by default. To employ GNOVA, we created the LD scores utilizing the 1000 Genomes of European ancestry using the *--save-ld* argument within the *gnova.py* script. For HDL, we used the provided scripts from HDL to generate the LD scores using the same 1000 Genomes of European ancestry (https://github.com/zhenin/HDL/wiki/Build-a-reference-panel).

Through our analysis, we found that the three packages have different levels of model convergence rates, which is critical for future applications as these open-source packages claim to advance genetic research. In particular, we found that LDSC (1,062,577/1,062,600) and GNOVA (1,062,600/1,062,600) converged for most of the tested MAE-DE pairs, whereas HDL failed a substantial proportion of the analyses, leading to only 59,291 out of the 1,062,600 MAE-DE pairs (refer to the raised issue: https://github.com/zhenin/HDL/issues/30). Therefore, in **Fig. 2**, we presented common significant results after Bonferroni corrections from the LDSC and GNOVA, resulting in 133 and 45 significant signals corrected on *i*) the number of MAEs and *ii*) the number of MAEs and DEs.

**Figure 2:**
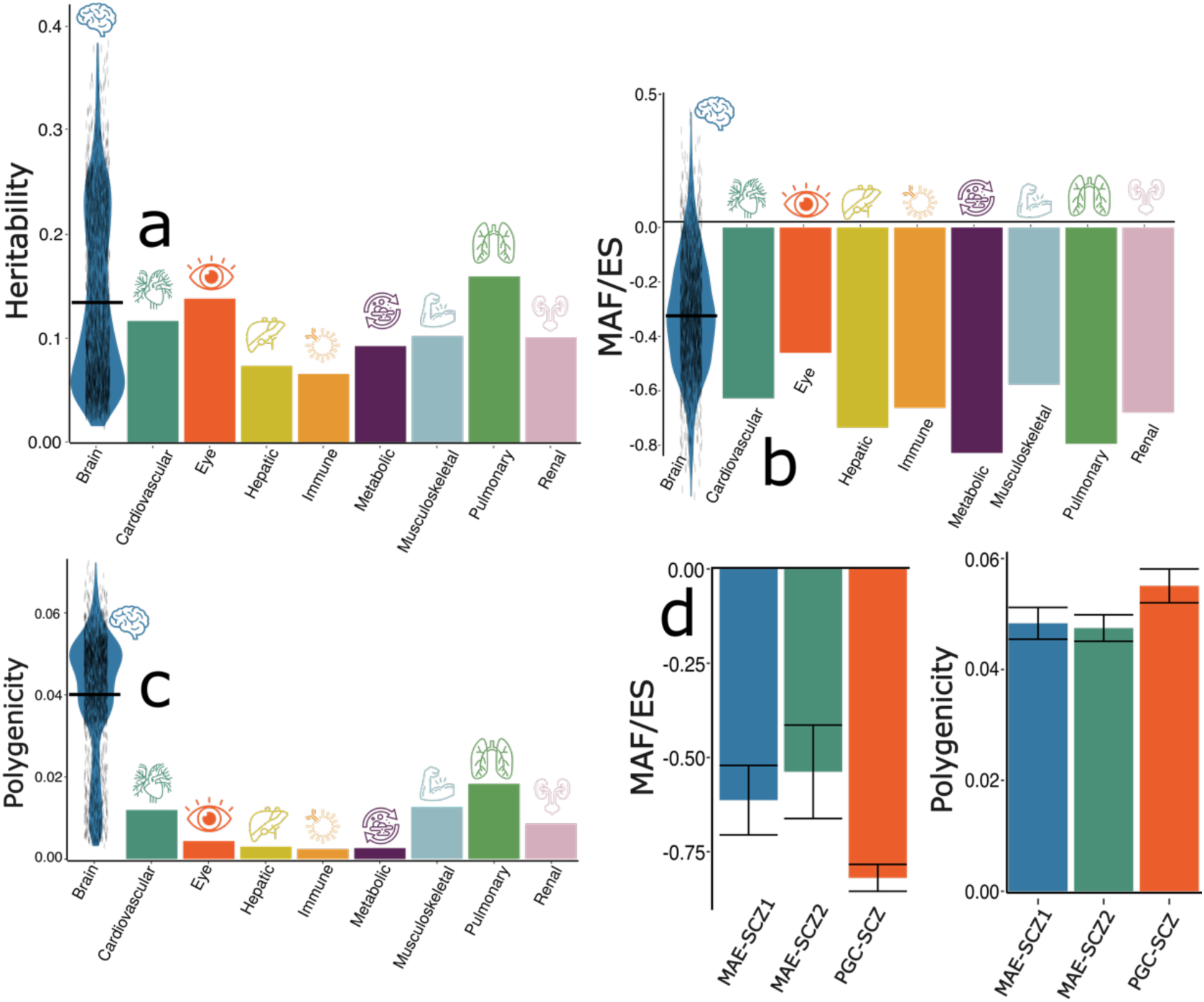
The genetic architecture of the 2024 MAEs. Three parameters are estimated by SBayesS to delineate the genetic architecture of the 2024 MAEs, including (**a**) the SNP-based heritability 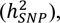 (**b**) the relationship between MAF and effect size (*S*), and (**c**) polygenicity (*π*). (**d**) We compared the *π* and *S* parameters using harmonized GWAS summary data for two AI- and imaging-derived subtypes (MAE-SCZ1 and MAE-SCZ2^4^) from UKBB and the disease endpoint of schizophrenia (PGC-SCZ^74^) from PGC. FinnGen data was not used due to bias stemming from the unavailability of FinnGen-specific linkage disequilibrium data (**Supplementary eMethod 1**). We present the distribution of the estimated parameters for the 2016 brain MAEs using a violin plot; the mean value is denoted by the black horizontal line. These results should be interpreted cautiously for comparative purposes due to limitations stemming from the lack of individual genotype data from FinnGen and PGC, differing linkage disequilibrium structures, and varying sample sizes.

#### (c): Two-sample bidirectional Mendelian randomization

We employed a bidirectional, two-sample Mendelian randomization using the *TwoSampleMR* package^47^ to infer the causal relationships between the 2024 MAEs, 521 DEs from FinnGen, and 4 brain DEs from PGC.

The forward Mendelian randomization examined causality from the 2024 MAEs to the 525 DEs, while the inverse analysis investigated causality from the 525 DEs to the 2024 MAEs. The *TwoSampleMR* package^47^ applied five different Mendelian randomization methods. We presented the significant findings after the Bonferroni correction using the inverse variance weighted (IVW) estimator, verifying that the correction remained significant in at least one of the other four estimators (Egger, weighted median, simple mode, and weighted mode estimators). For the significant signals, we performed several sensitivity analyses. First, a heterogeneity test was performed to check for violating the IV assumptions. Horizontal pleiotropy was estimated to navigate the violation of the IV’s exclusivity assumption^66^ using a funnel plot, single-SNP Mendelian randomization approaches, and Mendelian randomization Egger estimator. Moreover, the leave-one-out analysis excluded one instrument (SNP) at a time and assessed the sensitivity of the results to individual SNP.

Critically, to enhance transparency and reproducibility, we followed a systematic procedure guided by the STROBE-MR Statement^67^ in conducting all causality analyses. This comprehensive approach encompassed the selection of exposure and outcome variables, reporting full sets of statistics, and implementing sensitivity checks to identify potential violations of underlying assumptions. First, we performed an unbiased quality check on the GWAS summary statistics. Notably, the absence of population overlapping bias^29^ was confirmed, given that FinnGen and UKBB participants largely represent European ancestry populations without explicit overlap. For the four PGC DEs, we ensured that no UKBB participants were included in the GWAS summary data. Furthermore, all GWAS summary statistics were based on or lifted to GRCh37. Subsequently, we selected the effective exposure variables by assessing the statistical power of the exposure GWAS summary statistics in terms of instrumental variables (IVs), ensuring that the number of IVs exceeded 8 before harmonizing the data. Crucially, the function "*clump_data*" was applied to the exposure GWAS data, considering LD. The function "*harmonise_data*" was then used to harmonize the GWAS summary statistics of the exposure and outcome variables. This overall resulted in a smaller number (< 525 DEs or 2024 MAEs) of effective exposure/outcome variables in both forward and inverse Mendelian randomization analyses, as certain GWAS summary data did not have enough IVs.

#### (d): PRS calculation

PRS calculation used the GWAS summary statistics from the split-sample sensitivity analysis from our previous studies^12,3,1,4^. We established PRS weights using split1 GWAS data as the base/training set, while the split2 GWAS summary statistics were used as the target/testing data. Details of the quality control (QC) procedures are shown in our previous studies^12,3,1,4^. Following the QC procedures, PRS for the split2 group was computed using PRS-CS^68^. PRS-CS infers posterior SNP effect sizes under continuous shrinkage priors using GWAS summary statistics and an LD reference panel (i.e., UKBB reference). The shrinkage parameter was not set, and the algorithm learned it via a fully Bayesian approach.

After determining the optimal model, we applied the model to the entire UKBB sample (∼500k individuals). We then performed a PWAS to link the 2024 PRS-MAEs and 59 additional phenotypes (**Supplementary eTable 5)** not used to compute the PRS-MAE to avoid circular bias^69^. The 59 phenotypes include cognitive scores (e.g., fluid intelligence score; Field ID: 20016, mental traits (e.g., fed-up feelings; Filed ID: 1960), and lifestyle factors (e.g., tea intake; Filed ID: 1488). A linear regression was built considering the following covariates: sex (Field ID: 31), smoking status (Field ID: 20116), weight (Field ID: 21002), standing height (Field ID: 50), waist circumstance (Field ID: 48), age at recruitment (Field ID: 21022), and first 40 genetic principal components (Field ID: 22009).

## Results

### The genetic architecture of the 2024 MAEs and 525 DEs

We computed three parameters to fully depict the genetic architecture of the 2024 MAEs (**Method 3a**).

For the SNP-based heritability 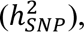 SBayesS^61^ obtained the highest 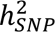 for the 2016 brain MAEs (mean 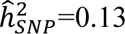 [0.01, 0.38]), followed by the pulmonary BAG (0.16±0.004), the eye BAG (0.14±0.009), the cardiovascular BAG (0.12±0.003), the renal BAG (0.10±0.003), and the musculoskeletal BAG (0.10±0.003) (**Fig. 2a** and **Supplementary eFile 1**). It is worth noting that SNP-based heritability varies across methods and depends on the input data, i.e., summary data or individual-level genotype data used in the method^70^. We aimed to benchmark the summary data-based methods by comparing the results from SBayesS with those of LDSC^28^ and SumHer^36^. Overall, while the estimates from the three methods were highly correlated (*r*=0.97 between LDSC and SumHer; *r*=0.99 between SBayesS and SumHer; *r*=0.99 between SBayesS and LDSC; **Supplementary eFigure 1**), SumHer (0.23±0.14) generally yielded larger 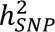 estimates than both LDSC (0.16±0.10) and SBayesS (0.13±0.08) (**Supplementary eFile 1**). This discrepancy across different data types (i.e., summary data vs. individual genotype data) and methods (e.g., LDSC vs. SBayesS) has been previously observed and compared in the literature, where the authors evaluated multiple methods designed for individual genotype data^70^. While we observed some differences in effect size magnitude across our three summary-data methods, the estimates remain highly correlated, ensuring internal validity within each method. However, we do not recommend directly comparing the numeric estimates across methods based on magnitude alone. Instead, greater emphasis should be placed on the confidence interval of standard errors of the estimates. We present the 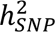 estimate of the 525 DEs and 2024 MAEs in **Supplementary eFigure 2**. **Supplementary eFile 2** presents the results of the 525 DEs. For the 525 DEs, we converted the 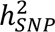 estimates from the observed scales to the liability scales, following the recommendations of Ojavee et al^71^. It’s important to clarify that we did not intend to compare the 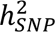 estimates of the two data sources due to differences in genotype coverage, sample sizes, allele frequencies, and other factors.

We then computed the natural selection signature (*S*) for the 2024 MAEs. The metabolic BAG showed a strong negative selection (S=-0.82±0.10), followed by the pulmonary BAG (S=-0.79±0.05), the hepatic BAG (S=-0.74±0.09), the renal BAG (S=-0.68±0.08), and the immune BAG (*S*=-0.66±0.11). For the brain MAEs (*S*=-0.33 [-1, 0.43]), the brain BAG and (S=-0.70±0.12) the subtype (ASD1) for autism spectrum disorder^51^ (S=-0.90±0.11) showed strong negative selection effects (**Fig. 2b** and **Supplementary eFile 3**).

Finally, we calculated the polygenicity (*π*) for the 2024 MAEs. We found that brain MAEs (0.040 [0.003, 0.072]) showed higher polygenicity than other organ systems (t-statistic=5.75; P-value=1.03×10^-8^), followed by the pulmonary BAG (0.018±0.001), the musculoskeletal BAG (0.013±0.001), and the cardiovascular BAG (0.011±0.001) (**Fig. 2c** and **Supplementary eFile 4**). The PSC (C128_115: https://labs-laboratory.com/bridgeport/MuSIC/C128_115) showed the highest polygenicity estimate (0.072±0.002).

### Potential evidence for the endophenotype hypothesis

Previous studies^72,73^ have found supporting evidence for the endophenotype hypothesis^14,15^ using traditional brain map-based signatures, showing that more genetic variants are associated with disease endpoints than imaging-derived signatures (i.e., endophenotypes). Of note, considering genetic differences between FinnGen and UKBB samples, SBayesS with the UKBB as LD reference may give biased estimates of *S* and *π* (LD from FinnGen not fully available; **Method 3a**). Therefore, we used the GWAS summary data for PGC schizophrenia (SCZ^74^) and two subtypes of SCZ (SCZ1 and SCZ2^4^) from our UKBB analysis to demonstrate this. The advantage of using PGC data is that the GWAS summary statistics are better powered (large sample sizes), and the data were from European ancestry groups across different countries. A data harmonization procedure is outlined in **Supplementary eMethod 1** to ensure a fair comparison of these estimates, which led to the utilization of a common set of SNPs and linkage disequilibrium information for computing the *S* and *π* parameters. Our results showed that MAE-SCZ1 (*π*=0.048±0.002; *S*=-0.61±0.09) and MAE-SCZ2 (*π*=0.047±0.002; *S*=-0.54±0.12) had lower polygenicity signals and weaker negative selection effects than PGC-SCZ (*π*=0.055±0.003; *S*=-0.82±0.04) (**Fig. 2d**). **Supplementary eFigure 3** shows the Manhattan plot of the harmonized summary data for MAE-SCZ1, MAE-SCZ2, and PGC-SCZ. These findings potentially support the endophenotype hypothesis^75^, which suggests that intermediate phenotypes (e.g., SCZ subtype MAEs) reside inside the causal pathway from genetics to exo-phenotypes (e.g., SCZ binary diagnosis), making them closer to the underlying etiology^72,73^.

### The genetic correlation shows organ-specific and cross-organ associations

We found 132 (P-value < 0.05/2024) and 45 (P-value < 0.05/2024/525) commonly significant positive genetic correlations (*r_g_*) after applying two levels of Bonferroni correction (**Fig. 3**) for the LDSC^28^ and GNOVA^37^ methods (**Method 3b**, **Supplementary eFile 5**, and **Supplementary eTable 2**). We noted that HDL encountered convergence issues with the models, as detailed in **Method 3b**.

**Figure 3:**
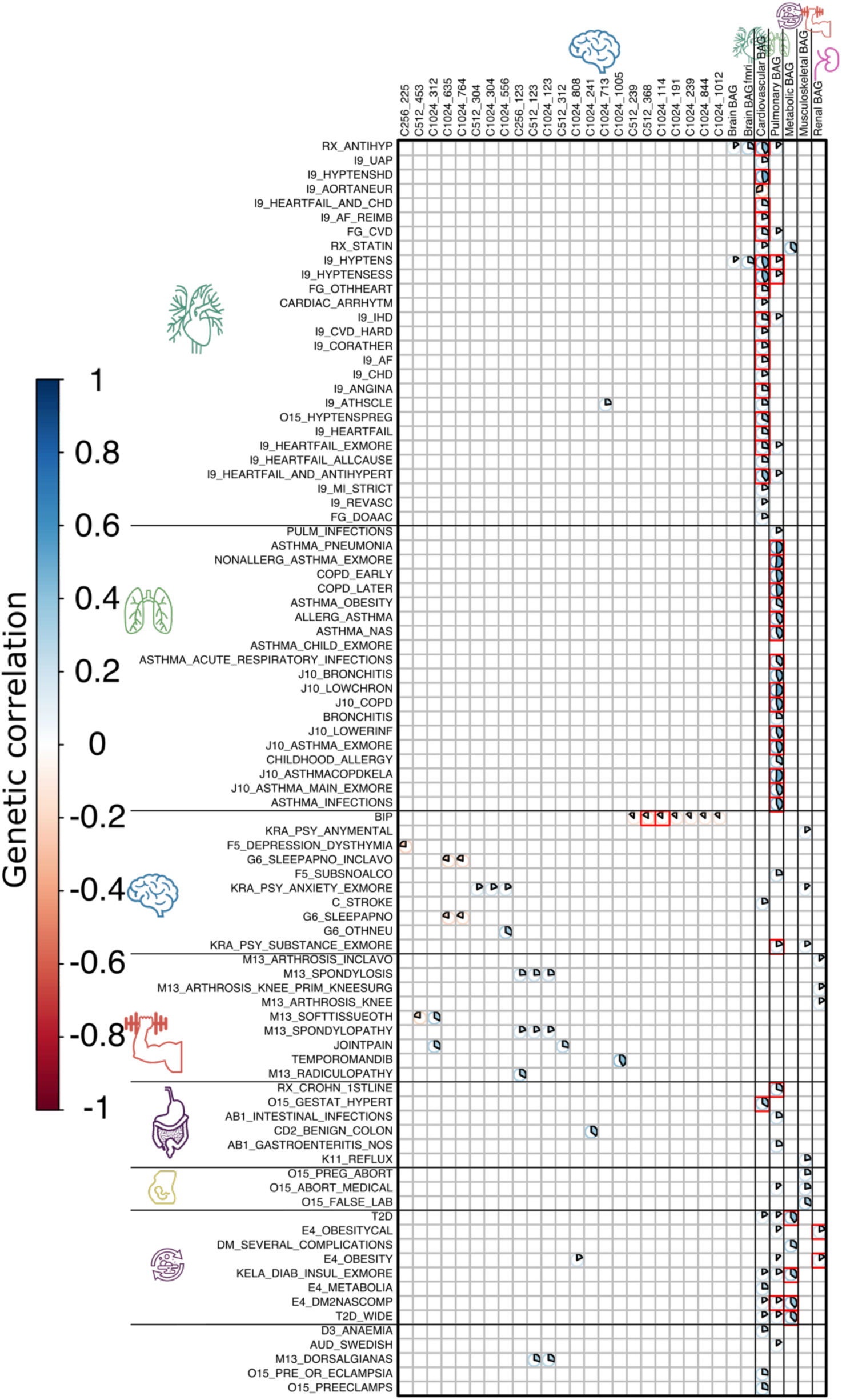
Genetic correlation between the 2024 MAEs and 525 Des. The significant genetic correlation estimates (*r_g_*) between 2024 MAEs and 525 DEs are depicted, considering two levels of corrections for multiple comparisons, considering the relatively smaller sample sizes (<40,000) for brain MAEs compared to other organ MAEs (>100,000). Initially, we reveal significant results shared between LDSC and GNOVA, employing Bonferroni correction based solely on the number of MAEs (P-value<0.05/2024), uncovering 133 MAE-AE pairs. Subsequently, a stricter correction based on both the number of MAEs and DEs is applied, leading to 45 unique MAE-AE pairs marked as red squares; the numeric results are displayed using results from LDSC. The genetic correlation for non-significant results was set to 0 for visualization purposes. For the MAEs, readers can explore the BRIDGEPORT portal for a visual representation of the 2003 brain PSCs (e.g., C256_225: https://labs-laboratory.com/bridgeport/MuSIC/C256_225) and the other BAGs at the MEDICINE portal: https://labs-laboratory.com/medicine.

Between these methods, the magnitude of the genetic correlations for the significant signals for both methods differed: mean 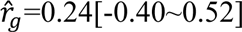 with 213 significant signals for LDSC, mean 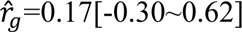 for GNOVA with 428 significant signals (**Fig. 2**). The three sets of converged estimates showed a strong correlation: *r*=0.77 (P-value<1×10^-10^; *N*=1,062,577) between LDSC and GNOVA, *r*=0.81 (P-value<1×10^-10^; *N*=59,289) between LDSC and HDL, and *r*=0.82 (P-value<1×10^-10^; *N*=59,289) between GNOVA and HDL. **Supplementary eFigure 4** shows the correlation of the three sets of estimates.

Within the significant signals identified, we observed *i*) organ-specific associations, in which the MAE showed a genetic association with the DE originating from the respective organ system, and *ii*) cross-organ connections, in which the MAE and DE were primarily involved from different organ systems. For example, two brain PSCs showed significant negative genetic correlations with BIP from PGC (C512_368 vs. BIP: -0.16±0.03; C1024_114 vs. BIP: - 0.15±0.03). At a less stringent level, the brain MAEs were also genetically associated with DEs from other organ systems, including the positive correlation between C1024_808 and obesity (E4_OBESITY: *r*_*g*_=0.17±0.13). The cardiovascular BAG was positively correlated with several DEs related to the cardiovascular system, including ischemic heart disease (I9_IHD: *r*_*g*_=0.26±0.03), coronary heart disease (I9_HEARTFAIL_AND_CHD: *r*_*g*_=0.26±0.03), angina (I9_ANGINA: *r*_*g*_=0.25±0.03) and atrial fibrillation (I9_AF: *r*_*g*_=0.22±0.04). Likewise, the pulmonary BAG was positively associated with multiple DEs related to the lung and respiratory system, including chronic obstructive pulmonary disease (COPD_EARLY: *r*_*g*_=0.47±0.04) and various forms of asthma (ASTHMA_NAS: *r*_*g*_=0.43±0.04). Cross-organ connections were established, such as between the pulmonary BAG and substance abuse (KRA_PSY_SUBSTANCE_EXMORE: *r*_*g*_=0.20±0.03) and hypertension (I9_HYPTENS: *r*_*g*_=0.17±0.03). Lastly, the metabolic BAG was largely linked to different forms of diabetes (T2D: *r*_*g*_=0.40±0.04).

### The brain, cardiovascular, and pulmonary MAEs are causally linked to DEs of multiple organ systems

Employing five distinct two-sample Mendelian randomization estimators, we identified 39 (P-value<0.05/633) and 15 (P-value<0.05/633/524) significant causal relationships, directed from the MAE to DE, that withstood the Bonferroni correction at two different levels of rigors, as per the inverse variance weighted (IVW) estimator and at least one of the other four estimators (**Method 3c** and **Supplementary eTable 3**).

Within the 15 significant causal relationships, the brain MAEs showed causal connections with DEs from the brain, as well as DEs from other organ systems. For example, the brain PSC (C1024_598) was causally linked to SCZ from PGC [P-value=9.89×10^-8^; OR (95% CI)=0.69 (0.59, 0.79); the number of IVs=7]. C1024_684 was causally linked to Ventral hernia from FinnGen [K11_VENTHER: P-value=1.09×10^-7^; OR (95% CI)=1.43 (1.25, 1.63); the number of IVs=18]. The pulmonary BAG was causally linked to multiple DEs related to the pulmonary system, including chronic obstructive pulmonary disease (COPD) [J10_COPD: P-value=2.70×10^-20^; OR (95% CI)=1.77 (1.56, 2.00); the number of IVs=59] and asthma [ASTHMA_PNEUMONIA: P-value=1.51×10^-14^; OR (95% CI)=1.67 (1.41, 1.96); the number of IVs=59]. The cardiovascular BAG was causally linked to ischemic heart disease (IHD) [ASTHMA_PNEUMONIA: P-value=1.09×10^-7^; OR (95% CI)=1.64 (1.36, 1.96); the number of IVs=37] (**Fig. 4**). **Supplementary eFile 6** presents the full set of results for the 521 FinnGen DEs and 4 PGC DEs. Mendelian randomization relies on stringent assumptions that can sometimes be violated. We showcased comprehensive sensitivity analyses for the significant signal from the cardiovascular BAG to ischemic heart disease (I9_IHD; **Supplementary eNote 1** and **Supplementary eFgiure 5**).

**Figure 4:**
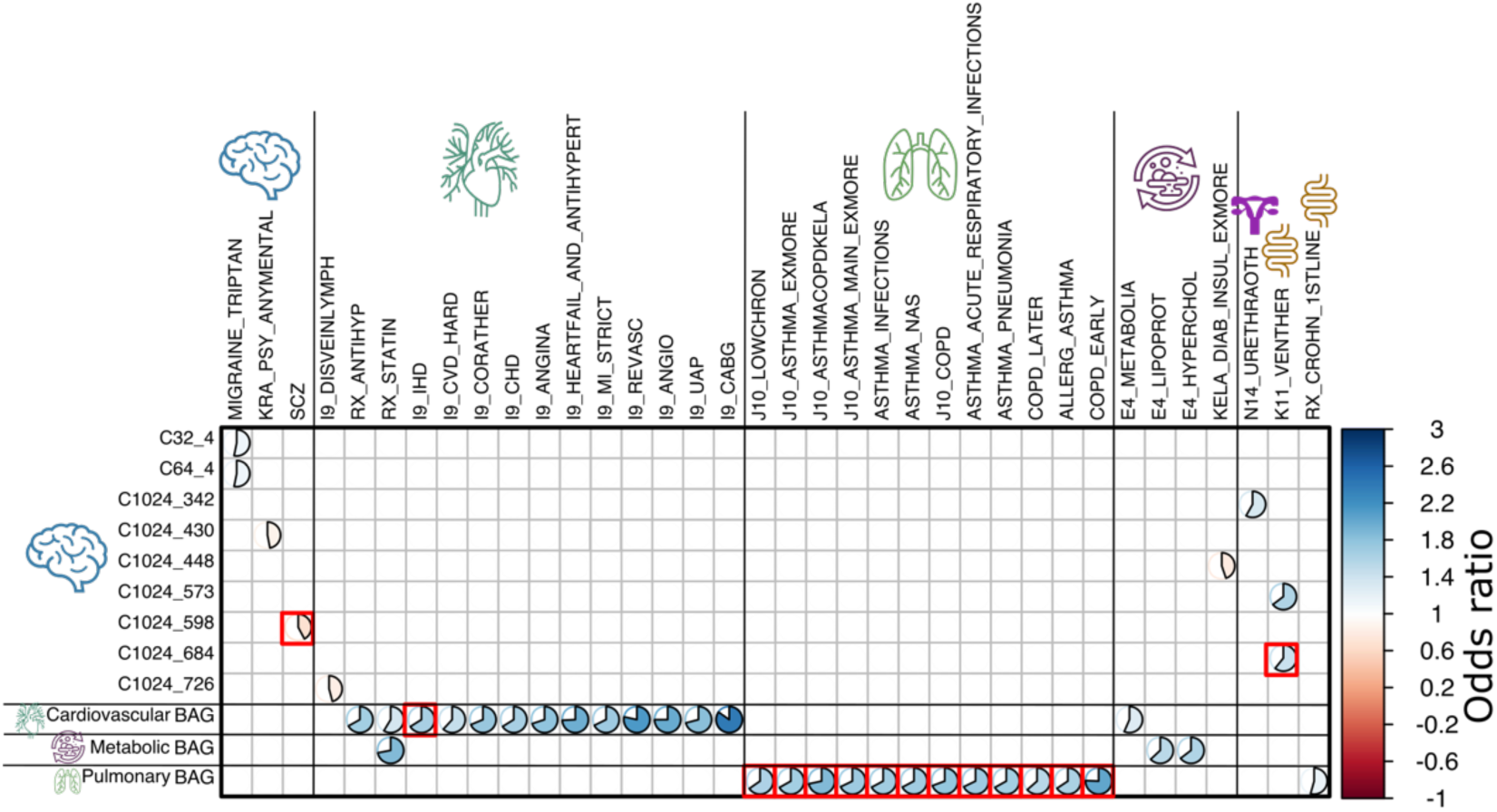
Causal relationship from the 2024 MAEs to the 525 Des. The causal relationship from the 2024 MAEs to the 525 DEs revealed 39 significant MAE-DE pairs, involving 633 MAEs as effective exposure variables (>8 instrumental variables before harmonization) and 525 DEs as outcomes. Bonferroni correction was applied to identify potential significant causal signals based on *i*) the 633 MAEs (P-value<0.05/633) and *ii*) the 633 MAEs and 525 DEs (P-value<0.05/633/524, denoted by the 15 red rectangles). Furthermore, we verified that the statistical significance attained for the IVW estimator was consistent and persisted across at least one of the other four Mendelian randomization estimators (Egger, weighted median, simple mode, and weighted mode estimators). For visualization purposes, the odds ratios for non-significant results were set to 1 and were left blank. For the MAEs, readers can explore the BRIDGEPORT portal for a visual representation of the 2003 brain PSCs (e.g., C32_4: https://labs-laboratory.com/bridgeport/MuSIC/C32_4) and the other BAGs at the MEDICINE portal: https://labs-laboratory.com/medicine. It is crucial to approach the interpretation of these potential causal relationships with caution despite our thorough efforts in conducting multiple sensitivity checks to assess any potential violations of underlying assumptions.

### The DEs involving Alzheimer’s disease, diabetes, asthma, and hypertension exert causal effects on multi-organ MAEs

We then tested the inverse causality by employing the DEs as exposure and MAEs as outcome variables. We identified 47 (P-value<0.05/787) and 23 (P-value<0.05/787/214) significant causal relationships, directed from the DE to MAE, that survived the Bonferroni correction at two different levels of rigors (**Method 3c** and **Supplementary eTable 4**).

Within the 23 significant causal relationships (P-value<0.05/787/214), various forms of Alzheimer’s disease were linked to the brain MAEs, including the brain BAG [G6_AD_WIDE: P-value=3.03×10^-7^; OR (95% CI)=1.10 (1.06, 1.13); the number of IVs=8] and metabolic BAG [G6_AD_WIDE: P-value=3.03×10^-7^; OR (95% CI)=1.07 (1.04, 1.09); the number of IVs=8]. Type 1 diabetes (E4_DM1NASCOMP) was also causally linked to multiple brain PSCs. In addition, the cardiovascular BAG was causally linked to multiple heart diseases, including hypertension [I9_HYPTENS: P-value=4.67×10^-31^; OR (95% CI)=1.23 (1.19, 1.27); the number of IVs=110]. Several forms of asthma were causally linked to the pulmonary BAG, such as allergic asthma [ALLERG_ASTHMA: P-value=2.38×10^-9^; OR (95% CI)=1.09 (1.06, 1.13); the number of IVs=14]. Finally, obesity was also linked to the renal BAG [E4_OBESITY: P-value=2.74×10^-8^; OR (95% CI)=1.11 (1.07, 1.15); the number of IVs=19] (**Fig. 5**). **Supplementary eFile 7** presents the full set of results for the 521 FinnGen DEs and 4 PGC DEs.

**Figure 5:**
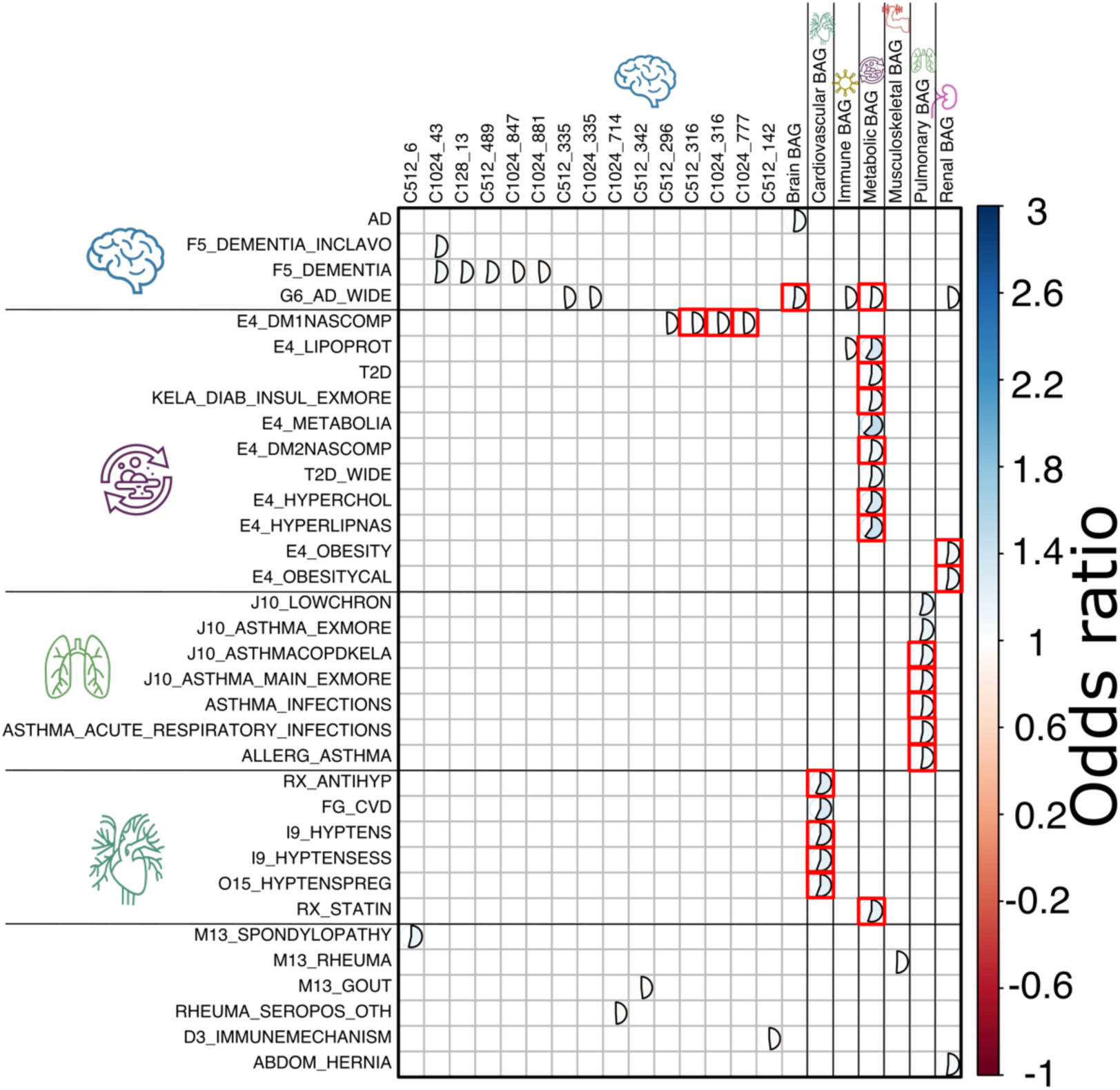
Causal relationship from the 525 DEs to the 2024 MAEs. The causal relationship from the 525 MAEs to the 2024 DEs revealed 47 significant DE-MAE pairs, involving 214 DEs as effective exposure variables (>8 instrumental variables before harmonization) and 787 DEs as effective outcomes after quality checks. Bonferroni correction was applied to identify potential significant causal signals based on *i*) the 787 MAEs (P-value<0.05/787) and *ii*) the 787 MAEs and 214 DEs (P-value<0.05/787/214, denoted by the 23 red rectangles). Furthermore, we verified that the statistical significance attained for the IVW estimator was consistent and persisted across at least one of the other four Mendelian randomization estimators (Egger, weighted median, simple mode, and weighted mode estimators). For visualization purposes, the odds ratios for non-significant results were set to 1 and were left blank. For the MAEs, readers can explore the BRIDGEPORT portal for a visual representation of the 2003 brain PSCs (e.g., C128_13: https://labs-laboratory.com/bridgeport/MuSIC/C128_13) and the other BAGs at the MEDICINE portal: https://labs-laboratory.com/medicine. It is crucial to approach the interpretation of these potential causal relationships with caution despite our thorough efforts in conducting multiple sensitivity checks to assess any potential violations of underlying assumptions.

### The polygenic risk scores of the 2024 MAEs

Using the PRS-CS^68^ method, we derived the PRS of the 2024 MAEs. We found that the 1799 MAEs could significantly (P-value<0.05/2024) predict the phenotypic BAGs in the test/target data (split2 GWAS; detailed in **Method 3d**). Among these, 1791 brain MAEs resulted in significant incremental *R^2^* ranging from 0.11% to 10.70% to predict the phenotype of interest. For example, the PSC (C1024_593 for part of the cerebellum: https://labs-laboratory.com/bridgeport/MuSIC/C1024_593) showed an incremental of *R^2^* 10.70%. The renal BAG showed an incremental *R^2^* of 5.92%, followed by the metabolic (*R^2^* = 5.67%) and pulmonary BAG (*R^2^* = 3.86%) (**Fig. 5a** and (**Supplementary eFile 8**).

We then applied the model to the entire UKBB population and performed a PRS-wide association study (PWAS), where the 2024 PRS-MAEs were linked to the 59 phenotypes that were not initially used to compute the respective PRS, to avoid the circular bias^69^ (**Supplementary eTable 5)**. Refer to **Method 3d** for details. We found 388 significant associations (P-value<0.05/2024/59) between 7 PRS-MAEs and 41 phenotypes. Among these, PSC C32_1 showed the most associations (94%); the lifestyle factor for only fish intake (Field ID: 16) was highly linked to multiple PRS-MAEs (16%). These results were expected because the 59 phenotypes (e.g., cognitive and mental traits) are primarily linked to the brain, and lifestyle factors were largely linked to multiple organ systems (**Fig. 5b** and **Supplementary eFile 9**). All derived PRS will be returned to UKBB and made available to the community.

**Figure 6:**
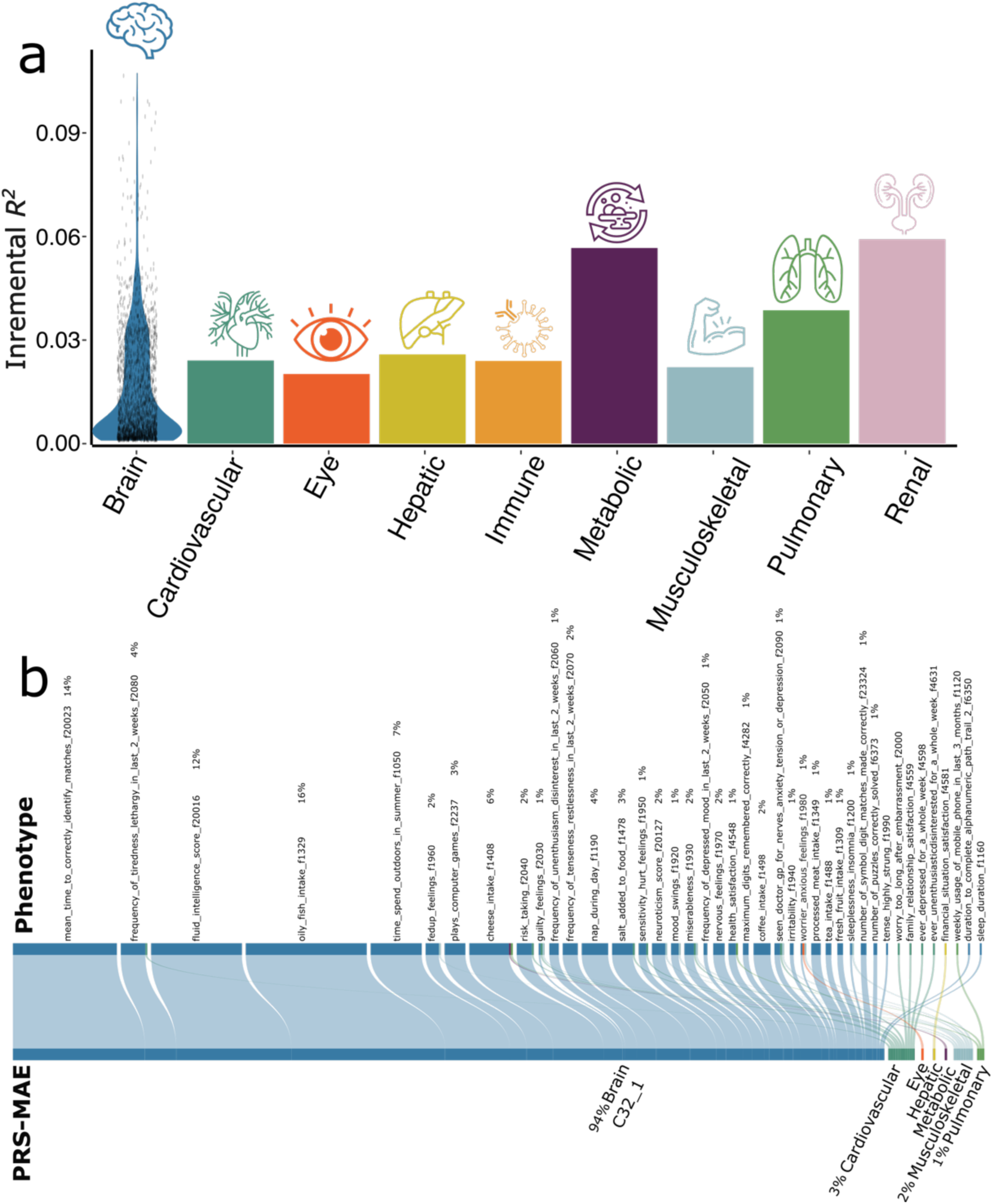
The polygenic risk score of the 2024 MAEs and PWAS. (**a**) The incremental *R^2^* of the PRS derived by PRC-CS to predict the 2024 MAEs in the target/test data (i.e., the split2 GWAS). The y-axis indicates the proportions of phenotypic variation that the PRS can significantly and additionally explain (i.e., incremental *R^2^*). The x-axis lists the 8 organ systems. For the brain, we showed the PRS distribution of the significant results from the 1791 brain PRS-MAEs; the other organ systems only have one PRS-MAE. (**b**) The PWAS links the PRS-MAEs to the 59 additional phenotypes not used to compute the PRS-MAE in the entire UKBB sample (P-value<0.05/2024/59).

## Discussion

This study expands previously established genetic atlases^76,35^ by integrating AI-derived endophenotypes via the 2024 MAEs within the multi-organ framework solely through GWAS summary statistics. We demonstrate a promising avenue for advancing imaging genetic research in two key aspects: *i*) integrating AI in imaging genetics and *ii*) exploring human aging and disease through a multi-organ perspective.

By comprehensively depicting the genetic architecture of the 2024 MAEs, we showcased that AI endophenotypes supported the endophenotype hypothesis^14,15^, in which they showed lower polygenicity and weaker negative selection effects than the disease diagnosis. First, it may suggest that these intermediate phenotypes exist along the causal pathway, bridging the gap between underlying genetics and "exo-phenotypes" like cognitive decline or disease diagnoses in case/control studies, thus positioned closer to the core etiology and pathology. Secondly, many of these 2024 MAEs originated from *in vivo* imaging methodologies like magnetic resonance imaging (MRI). Consequently, they tend to exhibit reduced noise levels (i.e., a higher SNR) in capturing disease-related effects and are less susceptible to biases, such as misclassification^77^, case/control-covariate sample bias (e.g., studies matching comorbidities and other factors), and imbalanced case/control ratios, as evidenced in many GWASs in FinnGen. Especially for the former, binary traits have a threshold for disease classification, leading to the dichotomization of individuals into affected and unaffected categories. Thirdly, the 525 DEs often represent complex diseases highly influenced by multiple genetic and environmental factors. Their multifaceted nature, involving numerous genes with modest effects and environmental interactions^78^, can lead to a higher vulnerability to disease onset and clinical symptoms. Consistent with this observation, we previously also found that one AI- and imaging-derived subtype of Alzheimer’s disease^32^ (AD1), but not the binary disease diagnosis, was genetically correlated with brain age (GM- and WM-BAG)^3^.

We observed that brain MAEs were overall more polygenic than MAEs from other organ systems. Brain disorders are highly polygenic^79^. First, the brain is a highly complex organ with intricate functions, and disorders affecting it are likely influenced by a larger number of genetic variants^12,80^. Second, many brain disorders are multifaceted, involving various aspects of brain structure, function, and connectivity, which can be influenced by various genetic factors^19^. Additionally, the brain regulates many physiological processes throughout the body, so disruptions in its function can have widespread effects, potentially involving interactions with multiple organ systems^1^. In addition, we found that most of the brain MAEs showed negative selection signatures, including the 9 disease subtype DNEs and 4 brain BAGs; some of the brain PSCs showed a positive *S* estimate (e.g., for the occipital lobe and subcortical structure, *S*=0.31±0.09: https://labs-laboratory.com/bridgeport/MuSIC/C32_18). The anticipated negative selection signatures of biological age across multiple organs and disease subtypes are expected to align with our prior findings, which revealed pervasive signatures of natural selection across a range of complex human traits and functional genomic categories. This negative selection signature prevents mutations with large deleterious effects from becoming frequent in the population^81^. The positive selection signatures identified in certain brain PSCs may suggest that positive selection may also play a role in shaping the genetic architecture of brain structural networks.

In previous studies on estimating SNP-based heritability, Elliott et al.^19^ used the SBAT (Sparse Bayesian Association Test) method with summary-level data to report heritability estimates for over 3,000 conventional imaging-derived phenotypes. For structural MRI-derived phenotypes – comparable to those derived from T1-weighted MRI in our 2,003 brain PSCs^12^ – they reported estimates of similar magnitude, if not lower for the mean, than ours. A potential reason for this difference is that we used the GCTA method^82^ with individual-level genetic data, which tends to provide more precise estimates in particular regarding LD patterns. This was further confirmed by Zhao et al., who also applied the GCTA method to estimate the heritability of these imaging-derived phenotypes^83^. Compared to previous studies using SBayesS to estimate polygenicity and selection signatures, Zeng and colleagues conducted two studies assessing these parameters across various traits, including disease endpoints (e.g., schizophrenia), cognitive traits (e.g., intelligence), and anthropometric (e.g., BMI) traits. Notably, in Zeng et al.^61^, the strongest negative selection signature was reported for dyslipidemia (*S*=-0.77), which we compare to our metabolic BAG in **Fig. 2b**.

The MUTATE atlas uncovered both established and previously undiscovered interactions concerning human systemic diseases within individual organs and across diverse organ systems. For example, within the cardiovascular system, the AI-derived MAE, cardiovascular BAG showed both substantial genetic correlation (**Fig. 2**) and bi-directional causality (**Fig. 3** and **4**) with multiple heart diseases, such as ischaemic heart disease^84^, heart failure^85^, and atrial fibrillation^86^. Similarly, pulmonary BAG was also causally linked to multiple diseases related to the lung and respiratory system, including COPD^87^ and various forms of asthma^88^. Another organ-specific connection was observed in neurologic diseases, encompassing conditions such as AD^89^ and various mental disorders^90^ linked to several MAEs associated with the brain, notably several PSCs and WM-BAG. Cross-organ interplay was evidenced for several novel connections. For instance, the brain PSCs exhibited causal connections to conditions extending beyond the brain, such as ventral hernia and vein diseases, as well as systemic conditions, like various forms of diabetes affecting the entire body. In contrast, AD appears to causally impact multiple BAGs across various human organ systems, including the renal, immune, and metabolic systems. It’s widely recognized that AD, being a complex condition, triggers detrimental effects that influence several human organ systems^89,91^. Our previous study used imaging genetics to investigate this multi-organ involvement along the disease continuum^92^. These results highlight the clinical relevance and interpretation of these AI endophenotypes to quantify individual-level organ health.

Emphasizing preventative strategies for specific chronic diseases is crucial to enhancing overall multi-organ health. Our MAEs present opportunities as novel instruments for selecting populations in clinical trials and facilitating therapeutic development. For example, one potential future avenue is to investigate whether these AI-driven disease subtype (e.g., AD DNEs) can be used to more effectively stratify patients for future AD clinical trial development and drug effect monitor. The hypothesis is that specific subtypes of AD patients may exhibit a better response to anti-amyloid therapies, with neuroimaging-derived MAEs serving as a selection tool to identify patients who are most likely to benefit, characterized by a high-risk, high-benefit profile. AD and various forms of diabetes exemplify disease endpoints significantly impacting multiple human organ systems. AD stands as the leading cause of dementia in older adults, presenting a persistent challenge in medicine despite numerous pharmacotherapeutic clinical trials. These trials have included interventions, such as anti-amyloid drugs^93,94^ and anti-tau drugs.^95^. The complexity and multifaceted nature of the underlying neuropathological processes may account for the lack of effective treatments. We call on the scientific community to embrace various mechanistic hypotheses to elucidate AD pathogenesis beyond amyloid and tau^96,97^. Likewise, the complexity of diabetes, with its various contributing factors, renders prevention challenging^98^. Moreover, diabetes often coexists with other chronic conditions affecting multiple organ systems, such as cardiovascular diseases, hypertension, and dyslipidemia^99^. Successful prevention strategies require a holistic approach, encompassing lifestyle adjustments, education, healthcare access, and societal considerations.

## Limitation

This study presents several limitations. Primarily, our analyses were centered solely on GWAS summary statistics derived from individuals of European ancestries. Future investigations should extend these findings to diverse ethnic groups, particularly those that are underrepresented, to ascertain broader applicability. This necessitates the research community’s commitment to embracing open science in AI and genetics. Secondly, the computational genomics statistical methods utilized in this research rely on several underlying statistical assumptions, which could potentially be violated and introduce bias. We mitigated bias by employing multiple methodologies to compute heritability, genetic correlation, and causality to address this concern. Additionally, we conducted thorough sensitivity checks, and the detailed results are provided accordingly. Additionally, our analysis was limited by the lack of individual-level genotype data from FinnGen and PGC, highlighting the need for future studies utilizing individual-level data to validate our empirical findings. Finally, our study recognizes a tradeoff between clinical interpretability and the detection of genetic associations when using AI-derived phenotypes.

## Outlook

In summary, we introduced the MUTATE genetic atlas to comprehensively comprehend the genetic architecture of AI endophenotypes and chronic diseases in multi-organ science. This investigation underscores the potential of integrating AI into genetic research and supports a comprehensive approach to investigating human diseases within a multi-organ paradigm.

## Data Availability

The results of the MUTATE atlas are disseminated at the MUTATE knowledge portal: https://labs-laboratory.com/mutate. The GWAS summary statistics for the 2024 MAEs can be accessed publicly through the MEDICINE knowledge portal: https://labs-laboratory.com/medicine and the BRIDGEPORT knowledge portal: https://labs-laboratory.com/bridgeport. The GWAS summary statistics for the 521 DEs from FinnGen are publicly available at: https://finngen.gitbook.io/documentation/v/r9/. The GWAS summary statistics for the 4 DEs from PGC are publicly available at: https://pgc.unc.edu/for-researchers/download-results/. The study used only GWAS summary statistics rather than individual-level data from the UK Biobank. However, the 2024 MAE GWAS data was initially derived from previous studies conducted under Application Numbers 35148 and 60698 from the UK Biobank.

## Code Availability

The software and resources used in this study are all publicly available:

- *GCTB*: https://cnsgenomics.com/software/gctb/#Overview, SNP-based heritability, polygenicity, and MAF/effect size ratio
- *LDSC*: https://github.com/bulik/ldsc, SNP-based heritability and genetic correlation
- *SumHer*: https://dougspeed.com/sumher/, SNP-based heritability
- *GNOVA*: https://github.com/xtonyjiang/GNOVA, genetic correlation
- *HDL*: https://github.com/zhenin/HDL, genetic correlation
- *TwoSampleMR*: https://mrcieu.github.io/TwoSampleMR/index.html, Mendelian randomization
- PRS-CS: https://github.com/getian107/PRScs, PRS
- Surreal-GAN: https://github.com/zhijian-yang/SurrealGAN, to derive AD1 and AD2
- HYDRA: https://github.com/anbai106/mlni, to derive LLD1-2, SCZ1-2, ASD1-3, and GM-, WM-, FC-BAG
- sopNMF: https://github.com/anbai106/SOPNMF, to derive the 2003 brain PSCs
- BioAge: https://github.com/yetianmed/BioAge, to derive the 9 multi-organ BAGs

## Authors’ contributions

Dr. Wen has full access to all the study data and is responsible for its integrity and accuracy.

*Study concept and design*: W.J

*Acquisition, analysis, or interpretation of data*: W.J

*Drafting of the manuscript*: W.J

*Critical revision of the manuscript for important intellectual content*: All authors

*Statistical analysis*: W.J

## Acknowledgment

The MULTI consortium (J.W) aims to integrate multi-organ and multi-omics biomedical data to advance our understanding of human aging and disease mechanisms. The current study does not use any individual data. The 2024 MAE GWAS summary data were derived from UK Biobank under Application numbers: 647044, 35148, and 60698. We sincerely thank the UK Biobank (https://www.ukbiobank.ac.uk/), FinnGen (https://www.finngen.fi/en), and PGC (https://pgc.unc.edu/) team for their invaluable contribution to advancing clinical research in our field.

