## Supplementary materials for "MUTATE: A Human Genetic Atlas of Multi-organ AI Endophenotypes using GWAS Summary Statistics"

#### eMethod 1: The harmonization procedure for the GWAS summary statistics of the PGC SCZ and UKBB SCZ subtypes

The GWAS summary statistics from UK Biobank, PGC, and FinnGen exhibit differences in genotype coverage and ethnicity, leading to variations in the estimation of linkage disequilibrium (LD). The SBayesS method necessitates precise LD estimation to compute the three parameters accurately. In our analyses, we observed biases in computing the *S* and *Pi* parameters, such as *Pi* values ~ 1 and *S* values below -1, for FinnGen when using an LD matrix computed from reference panel data from UK Biobank. For instance, SBayesS estimated a polygenicity (*Pi*) value of 0.98±0.02 for I9_HYPTENS (i.e., hypertension), suggesting that nearly all SNPs had non-zero effects in the GWAS. Similarly, H7_AMD (i.e., age-related macular degeneration) obtained a negative selection of *S*=-1.47. We contacted the FinnGen team to request the raw genotype data for the SuSie panel, which was originally utilized in FinnGen analyses. However, they were unable to share the data with us.

To test the endophenotype hypothesis, we used GWAS summary data from our UKBB MAEs (SCZ1 and SCZ2 subtypes) and the PGC SCZ. Ensuring that both datasets shared the same set of SNPs, we merged the SNPs common to both UKBB and PGC, resulting in 5,474,732 SNPs. Subsequently, we aligned these SNPs with the reference panel, flipped them according to the minor allele frequency, and removed outlier SNPs (>3 standard deviations from the median value), resulting in a final set of 736,053 SNPs located across the 22 chromosomes. By doing this, we can compute the three genetic parameters and compare them fairly.

**eMethod 2: AI methodologies employed in the current study**

- **a) sopNMF:**

The sopNMF algorithm is a stochastic approximation built and extended based on opNMF[1,2]. We consider a dataset of $n$ MR images and $d$ voxels per image. We represent the data as a matrix $\boldsymbol{X}$ where each column corresponds to a flattened image: $\boldsymbol{X} = [x_1, x_2, \ldots, x_n], \boldsymbol{X} \in \mathbb{R}_{\geq 0}^{d\times n}$. The sopNMF algorithm factorizes $\boldsymbol{X}$ into two low-rank ($r$) matrices $\boldsymbol{W} \in \mathbb{R}_{\geq 0}^{d\times r}$ and $\boldsymbol{H} \in \mathbb{R}_{\geq 0}^{r\times n}$ under the constraints of non-negativity and column-orthonormality. Using the Frobenius norm, the loss of this factorization problem can be formulated as

$$\|\boldsymbol{X} - \boldsymbol{WH}\|_F^2$$

$$\text{subject to } \boldsymbol{H} = \boldsymbol{W^T X}, \boldsymbol{W} \geq 0 \text{ and } \boldsymbol{W^T W} = \boldsymbol{I} \quad (1)$$

where $\boldsymbol{I}$ stands for the identity matrix. The columns $w_i \in \mathbb{R}^d$, $\|w_i\|^2 = 1, \forall\, i \in \{1..r\}$ of the so-called component matrix $\boldsymbol{W} = [w_1, w_2, \ldots, w_r]$ are part-based representations promoting sparsity in data in this lower-dimensional subspace. From this perspective, the loading coefficient matrix $\boldsymbol{H}$ represents the importance (weights) of each feature above for a given image. Instead of optimizing the non-convex problem in a batch learning paradigm (i.e., reading all images into memory) as opNMF,[1] sopNMF subsamples the number of images at each iteration, thereby significantly reducing its memory demand by randomly drawing data batches $\boldsymbol{X_b} \in \mathbb{R}_{\geq 0}^{d\times b}$ of $b \leq n$ images ($b$ is the batch size; $b$=32 was used in the current analyses); this is done without replacement so that all data goes through the model once ($\lceil n/b \rceil$). In this case, the updating rule can be rewritten as

$$\boldsymbol{W}_{t+1} = \boldsymbol{W}_t \frac{\left(\boldsymbol{X_b X_b^T W}\right)_t}{\left(\boldsymbol{W W^T X_b X_b^T W}\right)_t} \quad (2)$$

We calculate the loss on the entire dataset at the end of each epoch (i.e., the loss is incremental across all batches) with the following expression:

$$\sum_{i=1}^{\lceil n/b \rceil} \left\|\boldsymbol{X_{b\_i}} - \boldsymbol{W W^T X_{bi}}\right\|_F^2 \quad (3)$$

We evaluated the training loss and the sparsity of $\boldsymbol{W}$ at the end of each iteration. Moreover, early stopping was implemented to improve training efficiency and alleviate overfitting. We summarize the sopNMF algorithm in **SI Algorithm 1**.

We applied sopNMF to the training population (*N*=4000). The component matrix $\boldsymbol{W}$ was sparse after the algorithm converged with a pre-defined maximum number of epochs (100 by default) with an early stopping criterion. To build the MuSIC atlas, we clustered each voxel (row-wise) into one of the $r$ features/PSCs as follows:

$$\boldsymbol{M_j} = \text{argmax}_k(\boldsymbol{W_{j,k}}) \; (4)$$

where $\boldsymbol{M}$ is a $d$-dimensional vector and $j \in \{1..d\}$. The $j$-th element of $\boldsymbol{M}$ equals $k$ if $\boldsymbol{W_{j,k}}$ is the maximum value of the $j$-th row. Intuitively, $M$ indicates which of the $r$ PSCs each voxel belongs to. We finally projected the vector $\boldsymbol{M} \in \mathbb{R}_{\geq 0}^{d}$ into the original image space to visualize each PSC of the MuSIC atlas (**Fig. 1**). Of note, 13 PSCs have vanished in this process for *C*=1024: all 0 for these 13 vectors.

We present the algorithm as below:

**Algorithm 1:** Algorithm for sopNMF.

The source code of the Python implementation of sopNMF is available here: https://github.com/anbai106/SOPNMF

**Algorithm 1:** sopNMF

```
• Input: maximum number of epochs e, number of component C or r, batch
  size b, early stopping criteria θ (i.e., the loss without decreasing for a certain
  epochs) ;
• Output: W ∈ R^{d×r}, H ∈ R^{r×n} ;
• Initialization: W ;
if not θ or epoch ≠ e then
    for p ← 0 to e do
        for i ← 0 to t do
            Read mini-batch X_{bi}
            Update W_{i+1} via Eq. 2
        end
        loss = \sum_{i=1}^{\lceil n/b \rceil} ||X_{bi} − W W^T X_{bi}||_F^2 (Eq.3)
        if loss in θ then
            Stop
        else
            Shuffle X
            Continue
        end
    end
else
    Stop
end
```

- **b) Surreal-GAN**:

Surreal-GAN[3] is a novel deep representation learning method. Unlike the other semi-supervised methods which seek a categorical disease subtype, it dissects the neuroanatomical heterogeneity of brain diseases into *k* continuous variables. Compared to its precursor, the Smile-GAN model,[4] Surreal-GAN follows the same principle of semi-supervised clustering but solves several limitations of Smile-GAN. Semi-supervised clustering methods seek the so-called "*1-to-k*" mapping by learning distribution transformation from CN data to PT data. The following schematic figure demonstrates the principles of Smile-GAN and Surreal-GAN in the semi-supervised learning framework to disentangle AD neuroanatomical heterogeneity.

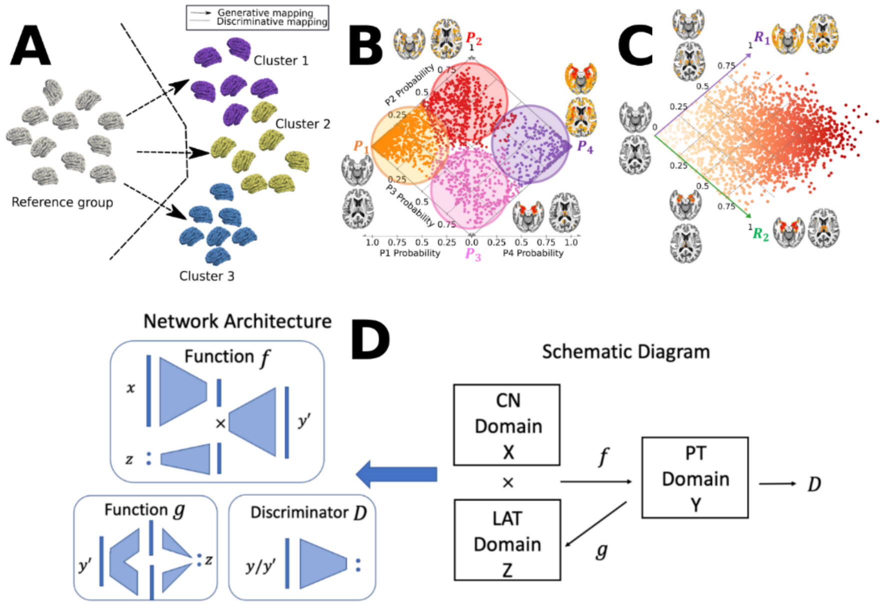


**Fig**. **A**) Schematic figures of semi-supervised clustering methods, including generative[3–5] and discriminative approaches.[6,7] Semi-supervised clustering methods dissect the neuroanatomical heterogeneity of brain diseases by seeking the so-called "*1-to-k*" mapping between the healthy control (CN) group as a reference and the patient (PT) group as a target. They sought to tease out clusters that are likely driven by distinct pathological trajectories instead of by global similarity/dissimilarity in data. The figure is adapted from our previous work.[8] **B**) Conceptual illustration of applying Smile-GAN ADNI to cluster MCI/AD participants into four hard-coded subtypes (P1, P2, P3, and P4) and two longitudinal pathways after applying the model to longitudinal data: P1→P2→P4; *ii*) P1→P3→P4. The figure is adapted from our previous work.[4] **C**) Conceptual illustration of applying Surreal-GAN to ADNI to recapitulate the neuroanatomical heterogeneity of MCI/AD participants into two continuous dimensional scores. Methodological advances are to capture disease severity and heterogeneity simultaneously using only cross-sectional data and allow the same individuals to express themselves along multiple dimensions. The figure is adapted from our previous work.[3] **D**) General network architectures (left) and schematic diagram of the semi-supervised learning (right, corresponding to Fig. A). The figure is adapted from our previous work.[3]

Surreal-GAN learns one transformation function $f$, which transforms CN data $\boldsymbol{x}$ to different synthesized PT data $\boldsymbol{y}' = f(\boldsymbol{x}, \boldsymbol{z})$, with latent variable $\boldsymbol{z}$ specifying distinct mapping directions. However, compared to Smile-GAN, Surreal-GAN considers that disease heterogeneity spatially and temporally (subtype) expands along a continuum (severity), similar to Sustain[9] to a certain extent. Thus, the latent variable $\boldsymbol{z}$ is modeled as a continuous variable, allowing infinite mapping directions from CN to PT data. $\boldsymbol{z} \sim p_{lat}(\boldsymbol{z})$ is sampled from a multivariate uniform distribution $U(0,1)^k$, rather than a categorical distribution as in Smile-GAN. Further, Surreal-GAN aims at disentangling spatial and temporal variations independently so that each dimension of the latent variable $\boldsymbol{z}$ is correlated with the severity of one relatively homogeneous imaging pattern. In contrast, different dimensions can be associated with spatially different imaging patterns.

To achieve the goals above, the objective function of Surreal-GAN consists of one adversarial loss and other regularization terms. The adversarial loss aims at matching the distribution of synthesized PT data, $p_{syn}$, and the distribution of real PT data, $p_{PT}$. Other regularization terms serve the following purposes: 1) encouraging sparse transformations (change loss); 2) reconstructing latent variables from synthesized or real PT data through the decomposer $g_1$ (decomposition loss) and reconstruction function $g_2$ (reconstruction loss); 3) boosting spatial separation of synthesized/captured patterns (orthogonality loss); 4) enforcing positive correlations between components of $\boldsymbol{z}$ and severity of synthesized patterns (monotonicity loss and cn loss).

Specifically, with distributions of CN, real PT, synthesized PT data denoted as $p_{CU}(\boldsymbol{x})$, $p_{PT}(\boldsymbol{y})$ and $p_{syn}(\boldsymbol{y}')$ respectively, the adversarial loss is defined as:

$$L_{GAN}(D,f) = E_{\boldsymbol{y}\sim p_{PT}(\boldsymbol{y})}\left[log\left(D(\boldsymbol{y})\right)\right] + E_{\boldsymbol{z}\sim p_{Lat}(\boldsymbol{z}),\boldsymbol{x}\sim p_{CN}(\boldsymbol{x})}\left[1 - log\left(D\left(f(\boldsymbol{x},\boldsymbol{z})\right)\right)\right]$$
$$= E_{\boldsymbol{y}\sim p_{PT}(\boldsymbol{y})}\left[log\left(D(\boldsymbol{y})\right)\right] + E_{p_{syn}(\boldsymbol{y}')}\left[1 - log\left(D(\boldsymbol{y}')\right)\right]$$

The transformation function attempts to synthesize PT data $\boldsymbol{y}'$, so that they follow similar distributions as real PT data. The discriminator, $D$, distinguishes the synthesized PT data from real PT data. Therefore, the discriminator is updated to maximize the adversarial loss, while the transformation function is optimized to minimize it.

Other regularization terms are introduced to further regularize the transformation function $f$. With the assumption that the disease process will not change brain anatomy dramatically and primarily only affect certain regions throughout most of the disease stages, the change loss is introduced to control sparsity and distance of transformations:

$$L_{change}(f) = E_{\boldsymbol{z}\sim p_{Lat}(\boldsymbol{z}),\boldsymbol{x}\sim p_{CU}(\boldsymbol{x})}[\|f(\boldsymbol{x},\boldsymbol{z}) - \boldsymbol{x}\|_1]$$

The decomposer $g_1$ serves to reconstruct changes synthesized by each component: $\boldsymbol{q}_i = f(\boldsymbol{x},\boldsymbol{a}^i) - \boldsymbol{x}$, where $\boldsymbol{a}_i^i = \boldsymbol{z}_i$ and $\boldsymbol{a}_j^i = 0$ for $j \neq i$. With $\widehat{\boldsymbol{q}}_{f(x,z)} = [\boldsymbol{q}_1^T, \boldsymbol{q}_2^T, \cdots, \boldsymbol{q}_M^T]^T$, the decomposition loss is defined as:

$$L_{decom}(f,g_1) = E_{\boldsymbol{z}\sim p_{Lat}(\boldsymbol{z}),\boldsymbol{x}\sim p_{CN}(\boldsymbol{x})}\left[\left\|g_1(f(\boldsymbol{x},\boldsymbol{z})) - \widehat{\boldsymbol{q}}_{f(x,z)}\right\|_2\right]$$

The reconstruction function $g_2$ serves to further reconstruct each component of the sampled $\boldsymbol{z}$ variable from $g_1\left(f(\boldsymbol{x},\boldsymbol{z})\right)$. The function $g$ is defined as a composition of $g_1$ and $g_2$, with $g\left(f(\boldsymbol{x},\boldsymbol{z})\right) = \left[g_2\left(g_1\left(f(\boldsymbol{x},\boldsymbol{z})\right)_{0:S}\right), \cdots, g_2\left(g_1\left(f(\boldsymbol{x},\boldsymbol{z})\right)_{S*(M-1):S*M}\right)\right]^T$ (S = number of ROIs), and the reconstruction loss is defined as

$$L_{recons}(f,g) = L_{recons}(f,g_1,g_2) = E_{\boldsymbol{z}\sim p_{Lat}(\boldsymbol{z}),\boldsymbol{x}\sim p_{CN}(\boldsymbol{x})}\left[\left\|g\left(f(\boldsymbol{x},\boldsymbol{z})\right) - \boldsymbol{z}\right\|_2\right]$$

The orthogonality loss aims to boost changes led each component, $\boldsymbol{q}_i$, to be relatively orthogonal to each other. For this purpose, a matrix $\boldsymbol{A}_{f(\boldsymbol{x},\boldsymbol{z})}$ is constructed with the $i_{th}$ column $A_{f(\boldsymbol{x},\boldsymbol{z})_{:,i}} = \boldsymbol{q}_i/||\boldsymbol{q}_i||_2$, and the orthogonality loss is defined as:

$$L_{ortho}(f) = E_{\boldsymbol{z}\sim p_{Lat}(z),\boldsymbol{x}\sim p_{CU}(\boldsymbol{x})}[||\boldsymbol{A}_{f(\boldsymbol{x},\boldsymbol{z})}^T\boldsymbol{A}_{f(\boldsymbol{x},\boldsymbol{z})} - I||_F]$$

To encourage a positive correlation between the severity of the synthesized pattern and the value of each component $\boldsymbol{z}_i$, another latent variable $\boldsymbol{z}'\sim p_{sev}(\boldsymbol{z}'|\boldsymbol{z})$ is sampled conditioned on previously sampled $\boldsymbol{z}$ variables, such that $\boldsymbol{z}_i' \geq \boldsymbol{z}_i$ for any $1 \leq i \leq k$. With these two sampled latent variables, the monotonicity loss is defined as:

$$L_{mono}(f) = E_{\boldsymbol{z}\sim p_{Lat}(\boldsymbol{z}),\boldsymbol{z}\sim p_{sev}(\boldsymbol{z}'|\boldsymbol{z}),\boldsymbol{x}\sim p_{CU}(\boldsymbol{x})}[||max\ (|f(\boldsymbol{x},\boldsymbol{z}) - \boldsymbol{x}| - |f(\boldsymbol{x},\boldsymbol{z}') - \boldsymbol{x}|,0)||_2]$$

The cn loss is introduced to further ensure that a small $\boldsymbol{z}$ variable lead to mild patterns. By letting $p_{cn}(\boldsymbol{z}) = U(0,0.05)^k$ to be a multivariate uniform distribution, the cn loss is defined as:

$$L_{cn}(f) = E_{\boldsymbol{z}^{cn} \sim p_{cn}(\boldsymbol{z}), \boldsymbol{x} \sim p_{CU}(\boldsymbol{x})}[\|f(\boldsymbol{x}, \boldsymbol{z}^{\boldsymbol{cn}}) - \boldsymbol{x}\|_1]$$

With all loss functions introduced above, the full objective of Surreal-GAN can be written as

$$L(D, f, g_1, g_2) = L_{GAN}(D, f) + \gamma L_{change}(f) + \kappa L_{decom}(f, g_1) + \zeta L_{recon}(f, g_1, g_2) + \lambda L_{ortho}(f) + \mu L_{mono}(f) + \eta L_{cn}(f)$$

With $\gamma$, $\kappa$, $\zeta$, $\lambda$, $\mu$, and $\eta$ being hyperparameters that control the relative importance of each loss function during the training process. More details of parameter selections can be found in the Surreal-GAN paper.[3]

Through the training process, parametrized functions $f$ , $g_1$, and $g_2$ are updated to satisfy that:

$$f, g_1, g_2 = arg \min_{f,g} \max_{D} L(D, f, g_1, g_2)$$

More importantly, after the training process, the function $g$, a composition of $g_1$ and $g_2$, can be applied to unseen PT data to infer the latent variable, which is referred to as the R-indices (i.e., dimensions) of PT data.

- **c) HYDRA**:

HYDRA leverages multiple ($k$) support vector machines (SVM) to seek the "*1-to-k*" mapping. It extends the $k$ linear SVMs' hyperplanes to the non-linear case piecewise, thereby constructing a $k$-face polytope for classification and clustering. This polytope, therefore, separates the CN group and the $k$ subpopulation of the PT group. Intuitively, each face of the convex polytope can be regarded to encode each subtype to derive each DNE – the distance of each participant to his/her nearest face of the $k$-face polytope.

To solve this joint optimization problem, the convex polytope is estimated by sequentially solving each linear SVM as a sub-problem under the principle of the sample-weighted SVM. The optimization stops until the sample weights get stable, i.e., the polytope was stably established. The objective of maximizing the polytope's margin can be summarized as:

$$\min_{\{w_j, b_j\}_{(j=1)}^{k}} \sum_{j=1}^{k} \frac{\|w_j\|_2^2}{2} + \mu \sum_{i|y_i=+1} \frac{1}{k} \max\{0, 1 - w_j^T X_j^T - b_j\} + \mu \sum_{i|y_i=-1} S_{i,j} \max\{0, 1 + w_j^T X_j^T + b_j\}$$

where $w_j$ and $b_j$ are the weight and bias for each hyperplane, respectively. $\mu$ is a penalty parameter on the training error, and $\boldsymbol{S}$ is the subtype membership matrix of dimension $n * k$ deciding whether a patient sample $i$ belongs to subtype $j$. The DNE indices are derived as:

$$DNE = w_j^T X_j^T + b_j$$

**eNote 1: Sensitivity check analyses for the MR results for the causal pathway of "Cardiovascular BAG→I9_IHD"**

As Mendelian randomization is sensitive to underlying IV assumptions, we performed sensitivity analyses to investigate the potential violation, exemplified by the potential causal relationship: cardiovascular BAG→I9_IHD.

In the original analysis, we observed a potential outlier instrumental variable (IV; i.e., independent SNPs: rs77870048) for the effect sizes on the exposure (i.e., cardiovascular BAG) and outcome variables (i.e., I9_IHD) (**eFig. 5 a**). We excluded those two outliers to rerun the analysis; the causal effect persisted, and the *beta* value slightly increased with a more significant P-value. We then performed a leave-one-IV-out analysis (**eFig. 5 b**) and showed the forest plot for the individual-SNP level of the causal effect sizes (**eFig. 5 c**), indicating that the outlier SNP contributed most to the originally observed causal effect. Finally, we showed a slight asymmetry funnel plot that indicates slightly asymmetry (**eFig. 5 d**). This heterogeneity is further supported by the heterogeneity analysis, evidenced by the Cochran's Q test ($P < 1x10^{-10}$) with the outlier SNP and Cochran's Q test ($P < 1x10^{-5}$) without the outlier SNP.

We further investigate whether the observed heterogeneity is driven by potential horizontal pleiotropy or if it arises from SNP-specific variability rather than directional pleiotropy. To this end, we applied MR-Egger regression with MAF-corrected weights to the summarized data, yielding an intercept estimate of 0.007 with an associated P-value of 0.45 (with the outlier SNP) and -0.009 with an associated P-value of 0.37 (without the outlier SNP). This further supports the absence of directional pleiotropy.

In summary, the MR-Egger intercept being close to zero and not statistically significant suggests no strong evidence of directional pleiotropy, indicating that the overall causal estimate is unlikely to be systematically biased by horizontal pleiotropy. The presence of an outlier SNP suggests that this variant may be contributing to the observed heterogeneity, potentially due to a strong pleiotropic effect or a violation of instrumental variable assumptions. Removing this outlier led to a stronger and more significant causal effect, suggesting that it may have introduced noise or bias that weakened the association. These findings indicate that the causal effect is robust and becomes more pronounced after accounting for potential outliers. The observed heterogeneity (Cochran's Q) is likely driven by SNP-specific variability rather than widespread pleiotropy, as the MR-Egger intercept does not suggest systematic bias.

**eFigure 1: The SNP-based heritability estimates between the three methods are highly correlated**

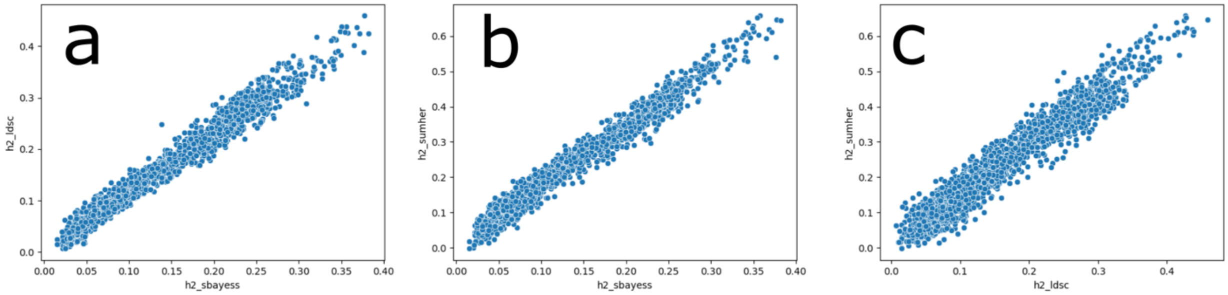


The scatter plot of the $h^2$ estimates between LDSC[10] and SBayesS[11] (**a**), SumHer[12] and SBayesS[11] (**b**), and LDSC[10] and SumHer[12].

**eFigure 2: The SNP-based heritability estimates for LDSC and SumHer for the 2024 MAEs and 525 DEs**

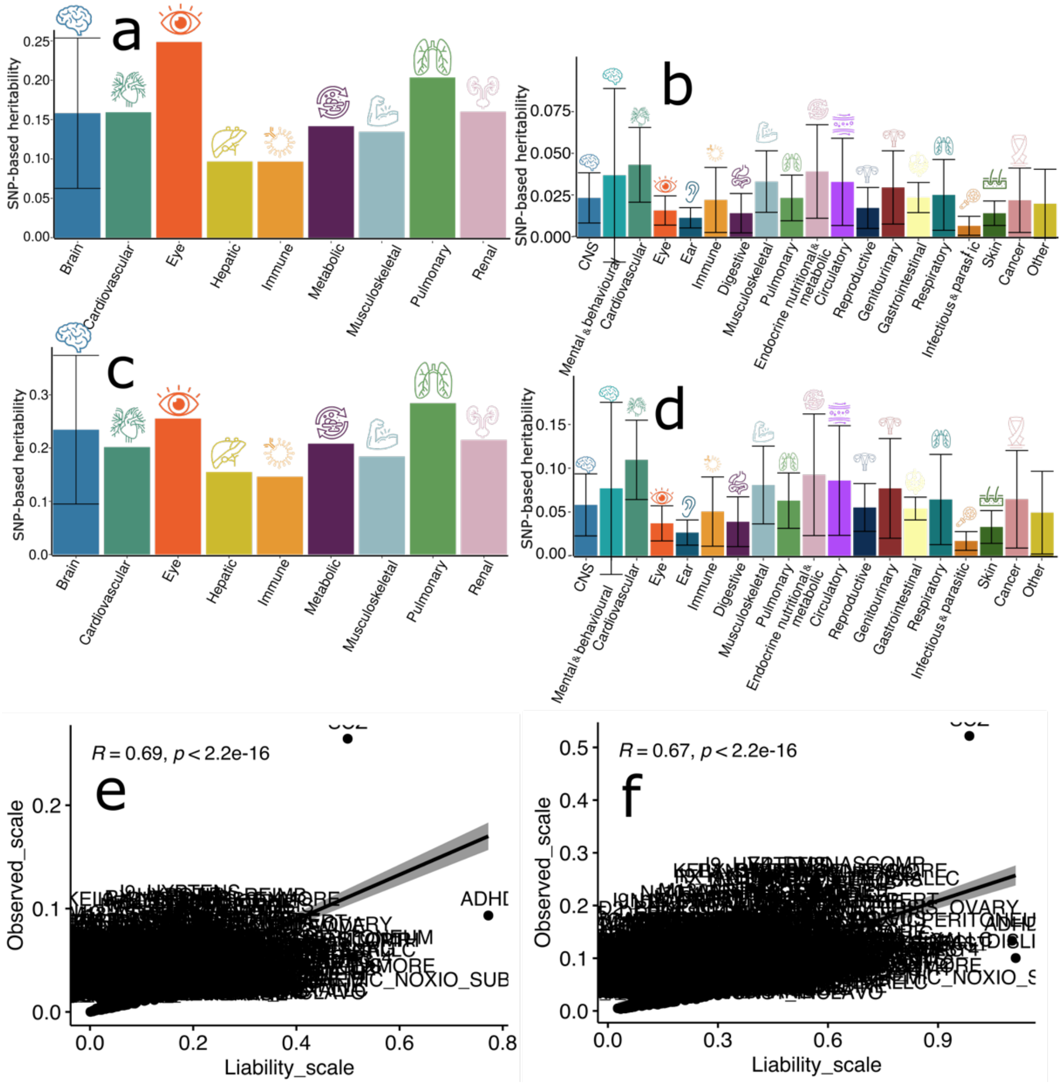


**a**) the $h^2$ of the 2024 MAEs using the LDSC software. **b**) the $h^2$ of the 525 DEs using the LDSC software. **c**) the $h^2$ of the 2024 MAEs using the SumHer software. **d**) the $h^2$ of the 525 DEs using the SumHer software. **e**) the scatter plot of the $h^2$ estimates of the 525 DEs at the observed scale and the liability scale using LDSC. **f**) the scatter plot of the $h^2$ estimates of the 525 DEs at the observed scale and the liability scale using SumHer. Abbreviations: CNS: central nervous system.

**eFigure 3: Manhattan plot for the UKBB SCZ subtypes vs. the PGC SCZ after data harmonization**

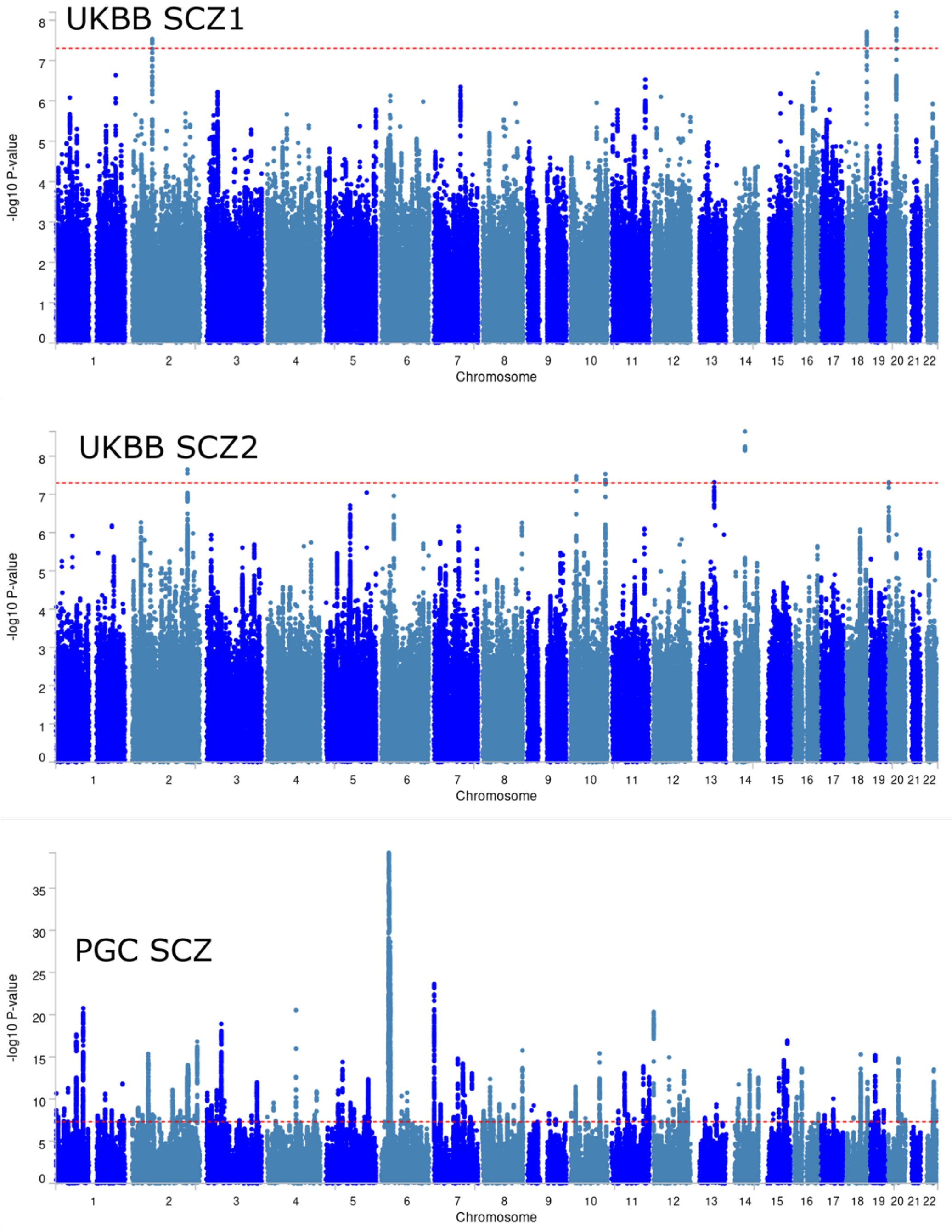


We harmonized the GWAS summary statistics from the UKBB SCZ subtypes and the PGC SCZ, identifying a total of 5,474,732 shared SNPs between the two datasets. Subsequently, we applied SBayesS on the harmonized data to calculate the three genetic parameters. Notably, polygenicity (*Pi*) is not influenced by sample sizes, ensuring the robustness of our finding that PGC SCZ exhibited higher *Pi* and *S*.

**eFigure 4: The genetic correlation estimates between the across software are highly correlated**

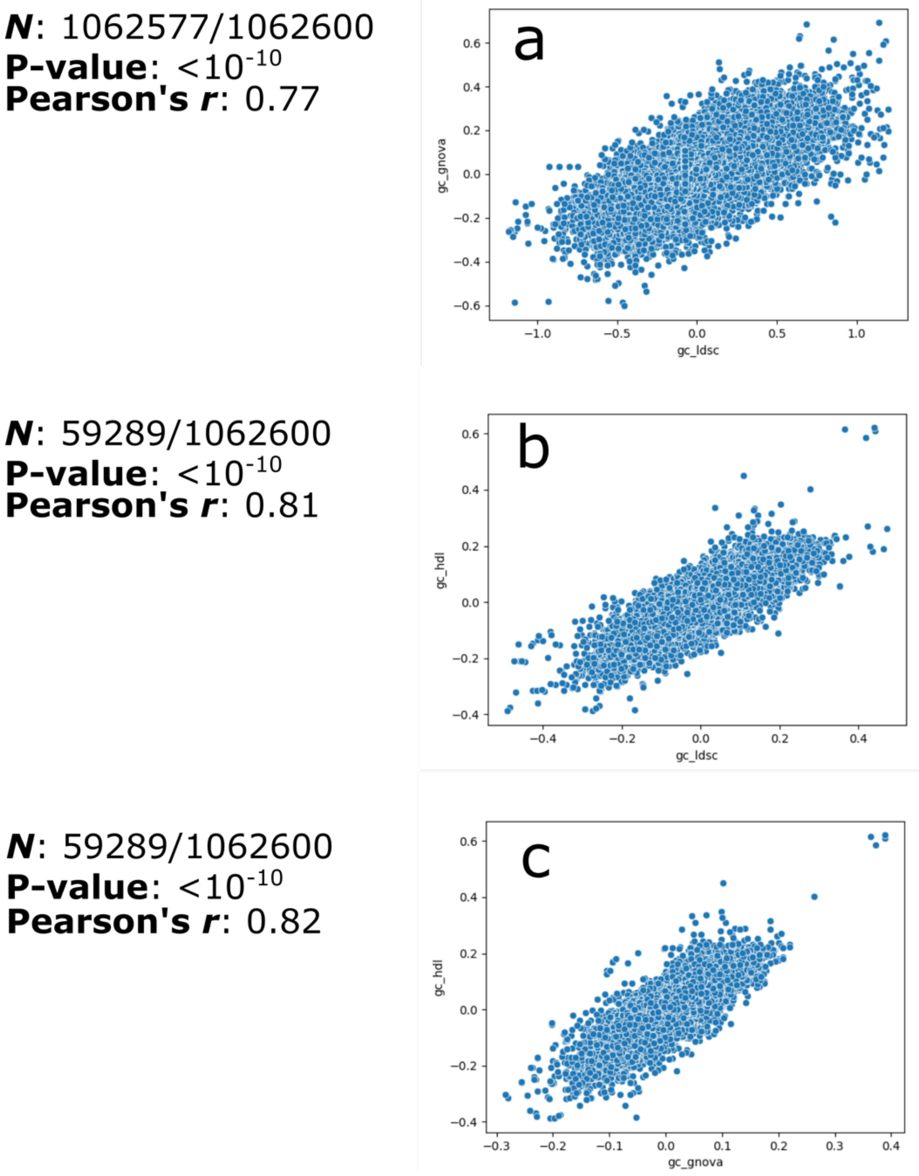


**a**) The scatter plot of the $r_g$ estimates between HDL and GNOVA; **b**) The scatter plot of the $r_g$ estimates between HDL and LDSC; **c**) The scatter plot of the $r_g$ estimates between GNOVA and LDSC.

**eFigure 5: Sensitivity check analyses for one example of our Mendelian randomization analysis: Cardiovascular BAG→I9_IHD**

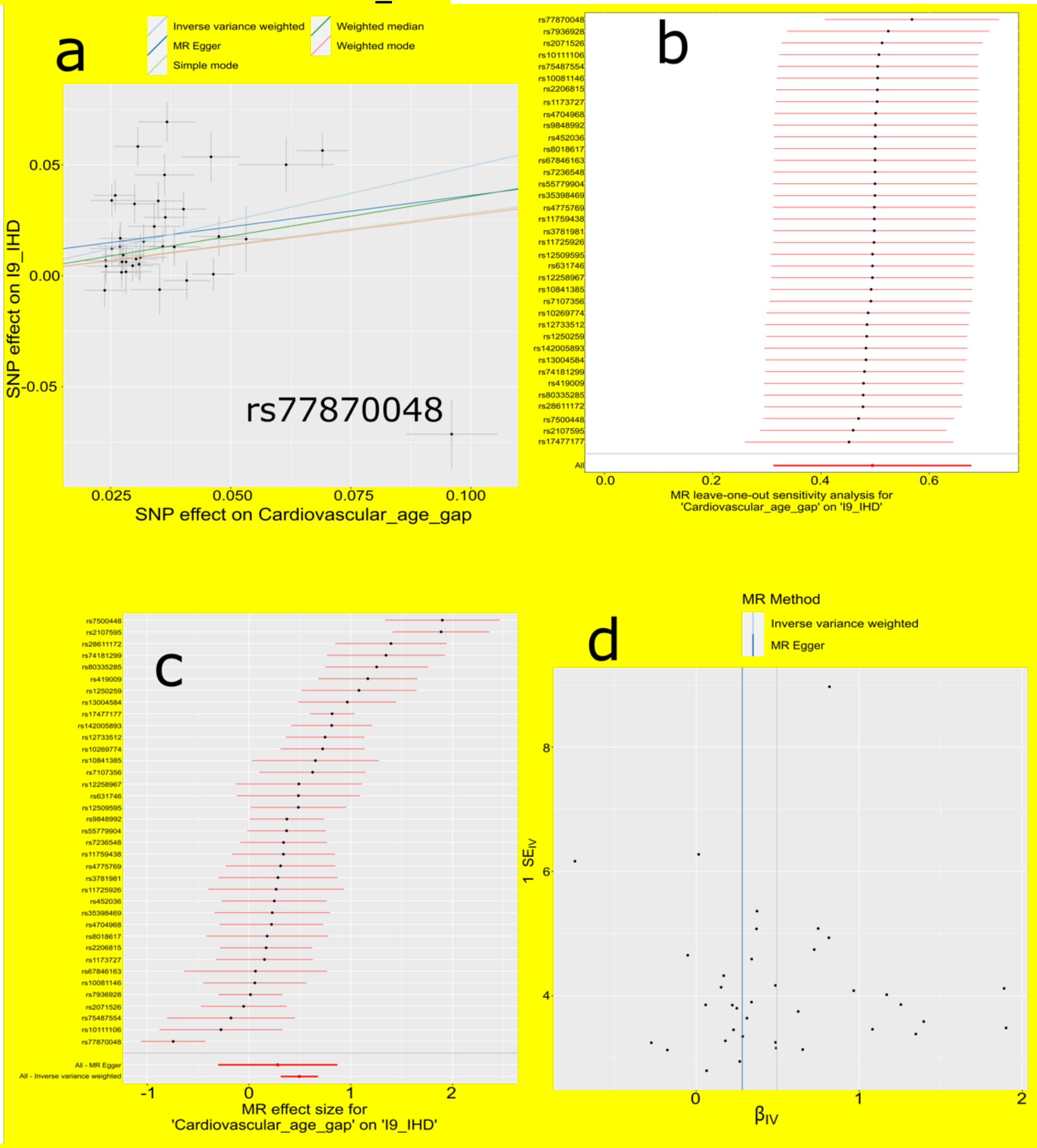


**a**) Scatter plot for the MR effect sizes of the SNP-Cardiovascular BAG association (*x*-axis, log OR) and the SNP-I9_IHD associations (*y*-axis, log OR) with standard error bars. The slopes of the five lines correspond to the causal effect sizes estimated by the five MR estimators, respectively. One obvious outlier IV was identified (rs77870048). We re-performed the MR by excluding this SNP and obtained a slightly stronger effect size than the original model. **b**) Leave-one-SNP-out analysis. Each dot represents the MR effect (log OR), and the error bar displays the 95% CI by excluding that SNP from the analysis. The red line depicts the IVW estimator using all SNPs. **c**) Forest plot for the single-SNP MR results. Each dot represents the MR effect (log OR)), and the error bar displays the 95% CI using only one SNP; the bottom lines show the MR effect using all SNPs together for IVW and MR Egger estimators. **d**) Funnel plot for the causal relationship. Each dot represents MR effect sizes estimated using each SNP as a separate instrument against the inverse of the standard error of the causal estimate.

#### eTable 1: The 2024 MAEs and 525 DEs

a) **2024 MAEs**: The 2003 brain PSCs originated from our prior investigation utilizing six scales of the MuSIC atlas[13] (C=32, 64, 128, 256, 512, and 1024, with 11 PSCs at C1024 omitted during optimization). The 9 brain DNEs were generated from our semi-supervised clustering and representation learning methods[14]. The 12 multi-organ BAGs were generated from a support vector regression model using different organ-specific features in our previous studies[15,16].

| Group | Scale (C) | Number of PSCs | MAE | N | Pubmed ID |
|---|---|---|---|---|---|
| MuSIC PSC | 32 | 32 | C32_*N* | 33,351 | 38127979 |
| | 64 | 64 | C64_*N* | | |
| | 128 | 128 | C128_*N* | | |
| | 256 | 256 | C256_*N* | | |
| | 512 | 512 | C512_*N* | | |
| | 1024 | 1013 | C1024_*N* | | |
| DNE | NA | NA | AD1 | 33,592 | 37662256 |
| | | | AD2 | | |
| | | | ASD1 | | |
| | | | ASD2 | | |
| | | | ASD3 | | |
| | | | LLD1 | | |
| | | | LLD2 | | |
| | | | SCZ1 | | |
| | | | SCZ2 | | |
| BAG | NA | NA | GM-BAG | 31,557 | 37333190 |
| | | | WM-BAG | 31,674 | |
| | | | FC-BAG | 32,017 | |
| | | | Brain BAG | 30,062 | 37398441 |
| | | | Cardiovascular BAG | 111,386 | |
| | | | Eye BAG | 36,004 | |
| | | | Hepatic BAG | 111,386 | |
| | | | Immune BAG | 111,386 | |
| | | | Musculoskeletal BAG | 111,386 | |
| | | | Metabolic BAG | 111,386 | |
| | | | Pulmonary BAG | 111,386 | |
| | | | Renal BAG | 111,386 | |

b) **521 DEs**: The 521 DEs were downloaded directly from the FinnGen study website.

| DE | Name | Category | Num_cases | Num_controls |
|---|---|---|---|---|
| RX_PARACETAMOL_NSAID | Paracetamol of NSAID medication | Drug purchase endpoints | 314977 | 62300 |
| RX_ANTIHYP | Antihypertensive medication - note that there are other indications | Drug purchase endpoints | 199546 | 177731 |
| FG_CVD | Cardiovascular diseases (excluding rheumatic etc) | Cardiometabolic endpoints | 185353 | 191924 |
| PAIN | Pain (limb, back, neck, head abdominally) | Miscellaneous, not yet classified endpoints | 171922 | 204598 |
| K11_CARIES_1_ONLYAVO | Dental caries 1, only avohilmo | XI Diseases of the digestive system (K11_) | 161113 | 216164 |
| M13_ARTHROPATHIES | Arthropathies | XIII Diseases of the musculoskeletal system and connective tissue (M13_) | 136415 | 240862 |
| RX_STATIN | Statin medication | IX Diseases of the circulatory system (I9_) | 127169 | 250108 |
| K11_ORAL | Diseases of oral cavity, salivary glands and jaws | XI Diseases of the digestive system (K11_) | 118043 | 259234 |
| PULM_INFECTIONS | COPD/asthma/ILD related infections | Interstitial lung disease endpoints | 117312 | 259965 |
| I9_HYPTENS | Hypertension | IX Diseases of the circulatory system (I9_) | 111581 | 265626 |
| ANTIDEPRESSANTS | Depression medications | Comorbidities of Neurological endpoints | 106785 | 88536 |
| O15_DELIV_SPONT | Single spontaneous delivery | XV Pregnancy, childbirth and the puerperium (O15_) | 106627 | 92306 |

| | | | | |
|---|---|---|---|---|
| M13_DORSOPATHY | Dorsopathies | XIII Diseases of the musculoskeletal system and connective tissue (M13_) | 106313 | 270964 |
| N14_FEMALEGENNONINF | Noninflammatory disorders of female genital tract | XIV Diseases of the genitourinary system (N14_) | 103306 | 107564 |
| M13_SOFTTISSUE | Soft tissue disorders | XIII Diseases of the musculoskeletal system and connective tissue (M13_) | 102065 | 275212 |
| KRA_PSY_ANYMENTAL | Any mental disorder | Psychiatric endpoints from Katri Räikkönen | 99751 | 277526 |
| M13_ARTHROSIS_INCLAVO | Artrosis, including avohilmo | XIII Diseases of the musculoskeletal system and connective tissue (M13_) | 96500 | 280777 |
| AUTOIMMUNE | Autoimmune diseases | Diseases marked as autimmune origin | 96150 | 281127 |
| J10_UPPERDIS | Other diseases of upper respiratory tract | X Diseases of the respiratory system (J10_) | 93935 | 283342 |
| FALLS | Falls/tendency to fall | Miscellaneous, not yet classified endpoints | 92857 | 284420 |
| I9_HYPTENSESS | Hypertension, essential | IX Diseases of the circulatory system (I9_) | 92462 | 265626 |
| G6_EPIPAROX | Episodal and paroxysmal disorders | VI Diseases of the nervous system (G6_) | 89440 | 287837 |
| K11_GINGIVITIS_PERIODONTAL | Gingivitis and periodontal diseases | XI Diseases of the digestive system (K11_) | 87497 | 259234 |
| K11_PULPITIS_1_ONLYAVO | Dental pulpitis 1, only avohilmo | XI Diseases of the digestive system (K11_) | 81713 | 295564 |
| M13_OTHERJOINT | Other joint disorders | XIII Diseases of the musculoskeletal system and connective tissue (M13_) | 81676 | 240862 |
| M13_DORSOPATHYOTH | Other dorsopathies, not elsewhere classified | XIII Diseases of the musculoskeletal system and connective tissue (M13_) | 79212 | 270964 |
| FG_OTHHEART | Other heart diseases | Cardiometabolic endpoints | 77711 | 191924 |
| RX_CROHN_1STLINE | First line medication for Crohn's disease | Drug purchase endpoints | 77497 | 299780 |
| C3_CANCER_EXALLC | Malignant neoplasm (controls excluding all cancers) | II Neoplasms, from cancer register (ICD-O-3) | 77325 | 287137 |
| K11_INTESTOTH | Other diseases of intestines | XI Diseases of the digestive system (K11_) | 75346 | 301931 |
| M13_ARTHROSIS | Arthrosis | XIII Diseases of the musculoskeletal system and connective tissue (M13_) | 73710 | 240862 |
| J10_UPPERINFEC | Acute upper respiratory infections | X Diseases of the respiratory system (J10_) | 69111 | 308166 |
| CARDIAC_ARRHYTM | Cardiac arrhythmias | Comorbidities of COPD | 67035 | 220224 |
| J10_LOWCHRON | Chronic lower respiratory diseases | X Diseases of the respiratory system (J10_) | 65991 | 311286 |
| RX_N05C | Use of hypnotics and sedatives | V Mental and behavioural disorders (F5_) | 65814 | 311463 |
| I9_IHD | Ischaemic heart disease, wide definition | IX Diseases of the circulatory system (I9_) | 63744 | 313533 |
| J10_INFLUPNEU | Influenza and pneumonia | X Diseases of the respiratory system (J10_) | 62604 | 314673 |
| O15_PREG_ABORT | Pregnancy with abortive outcome | XV Pregnancy, childbirth and the puerperium (O15_) | 61248 | 149622 |
| I9_CVD_HARD | Hard cardiovascular diseases | Cardiometabolic endpoints | 61240 | 316037 |
| H7_CATARACTSENILE | Senile cataract | VII Diseases of the eye and adnexa (H7_) | 59522 | 312864 |
| M13_DORSALGIA | Dorsalgia | XIII Diseases of the musculoskeletal system and connective tissue (M13_) | 59438 | 270964 |
| J10_PNEUMONIA | All pneumoniae | X Diseases of the respiratory system (J10_) | 58174 | 319103 |
| RX_CODEINE_TRAMADOL | Codeine or tramadol medication | Drug purchase endpoints | 57823 | 319454 |
| T2D | Type 2 diabetes, definitions combined | Diabetes endpoints | 57698 | 308252 |
| K11_OESSTODUO | Diseases of oesophagus, stomach and duodenum | XI Diseases of the digestive system (K11_) | 56890 | 320387 |
| E4_THYROID | Disorders of the thyroid gland | IV Endocrine, nutritional and metabolic diseases (E4_) | 56574 | 320703 |
| BRONCHITIS | Bronchitis | Asthma and related endpoints | 55222 | 322055 |
| K11_HERNIA | Hernia | XI Diseases of the digestive system (K11_) | 54691 | 322586 |
| KELA_DIAB_INSUL_EXMORE | Diabetes, insulin treatment (Kela reimbursement) (more control exclusions) | Diabetes endpoints | 53892 | 308249 |
| AUTOIMMUNE_NONTHYROID | Autoimmune diseases excluding thyroid diseases | Diseases marked as autimmune origin | 53213 | 280251 |
| I9_DISVEINLYMPH | Diseases of veins, lymphatic vessels and lymph nodes, not elsewhere classified | IX Diseases of the circulatory system (I9_) | 53156 | 324121 |
| E4_METABOLIA | Metabolic disorders | IV Endocrine, nutritional and metabolic diseases (E4_) | 53127 | 324150 |
| AUTOIMMUNE_NONTHYROID_STRICT | Autoimmune diseases excluding thyroid diseases, strict definition | Diseases marked as autimmune origin | 51863 | 275890 |
| N14_URINOTH | Other diseases of urinary system | XIV Diseases of the genitourinary system (N14_) | 49276 | 328001 |
| F5_DEPRESSION_DYSTHYMIA | Depression or dysthymia | V Mental and behavioural disorders (F5_) | 48847 | 225483 |
| O15_COMPLIC_LAB_DELIV | Complications of labour and delivery | XV Pregnancy, childbirth and the puerperium (O15_) | 48093 | 162777 |
| F5_MOOD | Mood [affective] disorders | V Mental and behavioural disorders (F5_) | 48085 | 329192 |
| I9_CORATHER | Coronary atherosclerosis | IX Diseases of the circulatory system (I9_) | 47550 | 313400 |
| N14_MALEGEN | Diseases of male genital organs | XIV Diseases of the genitourinary system (N14_) | 47110 | 119297 |
| G6_NERPLEX | Nerve, nerve root and plexus disorders | VI Diseases of the nervous system (G6_) | 46900 | 330377 |
| E4_DM2NASCOMP | Type 2 diabetes with other specified/multiple/unspecified complications | IV Endocrine, nutritional and metabolic diseases (E4_) | 46373 | 308280 |
| I9_AF | Atrial fibrillation and flutter | IX Diseases of the circulatory system (I9_) | 45766 | 191924 |
| H8_OTHEREAR | Other disorders of ear | VIII Diseases of the ear and mastoid process (H8_) | 45541 | 331736 |
| AB1_OTHER_BACTERIAL | Other bacterial diseases | I Certain infectious and parasitic diseases (AB1_) | 44934 | 332343 |
| M13_ARTHROSIS_KNEE | Gonarthrosis | XIII Diseases of the musculoskeletal system and connective tissue (M13_) | 44688 | 240862 |
| SLEEP | Sleep disorders (combined) | Neurological endpoints | 44299 | 329251 |
| I9_CHD | Major coronary heart disease event | IX Diseases of the circulatory system (I9_) | 43518 | 333759 |
| J10_CHRONTONSADEN | Chronic diseases of tonsils and adenoids | X Diseases of the respiratory system (J10_) | 43325 | 283342 |
| F5_DEPRESSIO | Depression | V Mental and behavioural disorders (F5_) | 43280 | 329192 |
| M13_SOFTTISSUEOTH | Other soft tissue disorders, not elsewhere classified | XIII Diseases of the musculoskeletal system and connective tissue (M13_) | 43190 | 275212 |
| J10_ASTHMA_EXMORE | Asthma (more control exclusions) | X Diseases of the respiratory system (J10_) | 42163 | 202399 |
| O15_MATERN_CARE | Maternal care related to the fetus and amniotic cavity and possible delivery problems | XV Pregnancy, childbirth and the puerperium (O15_) | 41941 | 168929 |
| M13_LOWBACKPAINORANDSCIATICA | Lower back pain or/and sciatica | XIII Diseases of the musculoskeletal system and connective tissue (M13_) | 41815 | 335462 |
| G6_SLEEPAPNO_INCLAVO | Sleep apnoea, including avohilmo | VI Diseases of the nervous system (G6_) | 41704 | 335573 |
| J10_ASTHMACOPDKELA | Asthma/COPD (KELA code 203) | X Diseases of the respiratory system (J10_) | 41495 | 311286 |
| E4_HYTHY_AI_STRICT | Hypothyroidism, strict autoimmune | IV Endocrine, nutritional and metabolic diseases (E4_) | 40926 | 274069 |
| AB1_INTESTINAL_INFECTIONS | Intestinal infectious diseases | I Certain infectious and parasitic diseases (AB1_) | 40881 | 336396 |
| L12_DERMATITISECZEMA | Dermatitis and eczema | XII Diseases of the skin and subcutaneous tissue (L12_) | 40688 | 336589 |
| DEATH | Any death | Common endpoint | 40455 | 336822 |
| KRA_PSY_ANXIETY_EXMORE | Anxiety disorders (more control exclusions) | Psychiatric endpoints from Katri Räikkönen | 40191 | 277526 |
| C_STROKE | STROKE | Comorbidities of Diabetes | 39818 | 271817 |
| MIGRAINE_TRIPTAN | Migraine, single triptan purchase ok & required. ICD-code if available is included | Neurological endpoints | 39387 | 337890 |
| G6_SLEEPAPNO | Sleep apnoea | VI Diseases of the nervous system (G6_) | 38998 | 336659 |
| T2D_WIDE | Type 2 diabetes, wide definition | Diabetes endpoints | 38657 | 310131 |

| | | | | |
|---|---|---|---|---|
| N14_RENALTUB | Renal tubulo-interstitial diseases | XIV Diseases of the genitourinary system (N14_) | 38335 | 338942 |
| M13_SPONDYLOPATHY | Spondylopathies | XIII Diseases of the musculoskeletal system and connective tissue (M13_) | 38276 | 270964 |
| E4_LIPOPROT | Disorders of lipoprotein metabolism and other lipidaemias | IV Endocrine, nutritional and metabolic diseases (E4_) | 37742 | 324150 |
| M13_INTERVERTEB | Other intervertebral disc disorders | XIII Diseases of the musculoskeletal system and connective tissue (M13_) | 37636 | 270964 |
| K11_CHOLELITH | Cholelithiasis | XI Diseases of the digestive system (K11_) | 37041 | 330903 |
| N14_MESNRUIRREG | Excessive, frequent and irregular menstruation | XIV Diseases of the genitourinary system (N14_) | 36824 | 107564 |
| J10_ACUTEUPPERINFEC | Acute upper respiratory infections of multiple and unspecified sites | X Diseases of the respiratory system (J10_) | 36445 | 308166 |
| O15_ABORT_MEDICAL | Medical abortion | XV Pregnancy, childbirth and the puerperium (O15_) | 36232 | 149622 |
| K11_CONSTIPATION | Constipation | XI Diseases of the digestive system (K11_) | 36022 | 341255 |
| BLEEDING | Bleeding | Miscellaneous, not yet classified endpoints | 35917 | 341360 |
| M13_KNEEDERANGEMENTS | Internal derangement of knee | XIII Diseases of the musculoskeletal system and connective tissue (M13_) | 35511 | 240862 |
| M13_SHOULDER | Shoulder lesions | XIII Diseases of the musculoskeletal system and connective tissue (M13_) | 34594 | 275212 |
| I9_ANGINA | Angina pectoris | IX Diseases of the circulatory system (I9_) | 34456 | 313400 |
| J10_ASTHMA_MAIN_EXMORE | Asthma (only as main-diagnosis) (more control exclusions) | X Diseases of the respiratory system (J10_) | 34343 | 202399 |
| N14_URETHRAOTH | Other disorders of urethra and urinary system | XIV Diseases of the genitourinary system (N14_) | 32905 | 328001 |
| H7_CHOROIDRETINA | Disorders of choroid and retina | VII Diseases of the eye and adnexa (H7_) | 32708 | 344569 |
| H7_LIDLACRIMALORBIT | Disorders of eyelid, lacrimal system and orbit | VII Diseases of the eye and adnexa (H7_) | 32593 | 344684 |
| H8_HL_SEN_NAS | Sensorineural hearing loss | VIII Diseases of the ear and mastoid process (H8_) | 32487 | 331736 |
| K11_HERING | Inguinal hernia | XI Diseases of the digestive system (K11_) | 32335 | 322586 |
| M13_POLYARTHROPATHIES | Polyarthropathies | XIII Diseases of the musculoskeletal system and connective tissue (M13_) | 32199 | 240862 |
| CD2_BENIGN_LEIOMYOMA_UTERI | Leiomyoma of uterus | II Neoplasms from hospital discharges (CD2_) | 31661 | 179209 |
| E4_PCOS_CONCORTIUM | Polycystic ovarian syndrome, consortium definition | IV Endocrine, nutritional and metabolic diseases (E4_) | 31548 | 179322 |
| O15_PREG_OTHER_MAT_DISORD | Other maternal disorders predominantly related to pregnancy | XV Pregnancy, childbirth and the puerperium (O15_) | 30971 | 179899 |
| K11_DIVERTIC | Diverticular disease of intestine | XI Diseases of the digestive system (K11_) | 30649 | 301931 |
| M13_LIMBPAIN | Pain in limb | XIII Diseases of the musculoskeletal system and connective tissue (M13_) | 30485 | 275212 |
| K11_PARODON_OPER | Parodontitis or operation codes | XI Diseases of the digestive system (K11_) | 30377 | 346900 |
| N14_PROSTHYPERPLA | Hyperplasia of prostate | XIV Diseases of the genitourinary system (N14_) | 30066 | 119297 |
| I9_VARICVE | Varicose veins | IX Diseases of the circulatory system (I9_) | 29539 | 324121 |
| AB1_GASTROENTERITIS_NOS | Diarrhoea and gastroenteritis of presumed infectious origin | I Certain infectious and parasitic diseases (AB1_) | 29401 | 336396 |
| M13_LOWBACKPAIN | Low back pain | XIII Diseases of the musculoskeletal system and connective tissue (M13_) | 29329 | 270964 |
| H7_CONJUNCTIVITIS | Conjunctivitis | VII Diseases of the eye and adnexa (H7_) | 28895 | 345122 |
| K11_APPENDACUT | Acute appendicitis | XI Diseases of the digestive system (K11_) | 28745 | 346283 |
| I9_OTHARR | Other arrhythmias | IX Diseases of the circulatory system (I9_) | 28079 | 286109 |
| K11_PARODON_OPER_DENTAL_OPER | Parodontitis or/and operation codes | XI Diseases of the digestive system (K11_) | 27833 | 349444 |
| JOINTPAIN | Pain in joint | XIII Diseases of the musculoskeletal system and connective tissue (M13_) | 27688 | 205355 |
| J10_TONSILLECTOMY | Tonsillectomy | X Diseases of the respiratory system (J10_) | 27449 | 349828 |
| D3_ANAEMIA | Anaemias | III Diseases of the blood and blood-forming organs and certain disorders involving the immune mechanism (D3_) | 27371 | 88536 |
| I9_HEARTFAIL | Heart failure,strict | IX Diseases of the circulatory system (I9_) | 27304 | 349973 |
| I9_HEARTFAIL_EXMORE | Heart failure,strict (more control exclusions) | IX Diseases of the circulatory system (I9_) | 27304 | 328105 |
| G6_OTHNEU | Other neurological diseases | VI Diseases of the nervous system (G6_) | 27026 | 350251 |
| I9_HEARTFAIL_ALLCAUSE | All-cause Heart Failure | IX Diseases of the circulatory system (I9_) | 26872 | 349361 |
| K11_CHOLECYSTECTOMY | Cholecystectomy | XI Diseases of the digestive system (K11_) | 26778 | 350499 |
| K11_REFLUX | Gastro-oesophageal reflux disease | Interstitial lung disease endpoints | 26184 | 320387 |
| K11_APPENDECTOMY | Appendectomy | XI Diseases of the digestive system (K11_) | 26013 | 351264 |
| E4_HYPERCHOL | Pure hypercholesterolaemia | IV Endocrine, nutritional and metabolic diseases (E4_) | 25928 | 324150 |
| I9_HEARTFAIL_AND_ANTIHYPERT | Heart failure and antihypertensive medication | Cardiometabolic endpoints | 25827 | 177731 |
| I9_STR | Stroke, excluding SAH | IX Diseases of the circulatory system (I9_) | 25398 | 339920 |
| K11_CARIES_2 | Dental caries 2 | XI Diseases of the digestive system (K11_) | 25095 | 352182 |
| H8_MIDDLEMASTOID | Diseases of middle ear and mastoid | VIII Diseases of the ear and mastoid process (H8_) | 24706 | 352571 |
| F5_ALLANXIOUS | All anxiety disorders | V Mental and behavioural disorders (F5_) | 24662 | 337577 |
| L12_OTHERSKINSUBCUTIS | Other disorders of skin and subcutaneous tissue | XII Diseases of the skin and subcutaneous tissue (L12_) | 24189 | 353088 |
| I9_MI_STRICT | Myocardial infarction, strict | IX Diseases of the circulatory system (I9_) | 24185 | 313400 |
| M13_MENISCUSDERANGEMENTS | Meniscus derangement | XIII Diseases of the musculoskeletal system and connective tissue (M13_) | 24158 | 240862 |
| M13_ROTATORCUFF | Rotator cuff syndrome | XIII Diseases of the musculoskeletal system and connective tissue (M13_) | 24061 | 275212 |
| N14_PYELONEPHR | Acute tubulo-interstitial nephritis | XIV Diseases of the genitourinary system (N14_) | 23912 | 338942 |
| H7_RETINALDISOTH | Other retinal disorders | VII Diseases of the eye and adnexa (H7_) | 23610 | 344569 |
| KRA_PSY_SUBSTANCE_EXMORE | Substance abuse (more control exclusions) | Psychiatric endpoints from Katri Räikkönen | 23194 | 277526 |
| I9_REVASC | Coronary revascularization (ANGIO or CABG) | IX Diseases of the circulatory system (I9_) | 23139 | 313400 |
| N14_FEMALEGENINF | Inflammatory diseases of female pelvic organs | XIV Diseases of the genitourinary system (N14_) | 22575 | 188295 |
| G6_CARPTU | Carpal tunnel syndrome | VI Diseases of the nervous system (G6_) | 22426 | 330377 |
| M13_ARTHTROSIS_COX | Coxarthrosis, | XIII Diseases of the musculoskeletal system and connective tissue (M13_) | 22254 | 240862 |
| FG_DOAAC | Diseases of arteries, arterioles and capillaries (FINNGEN) | IX Diseases of the circulatory system (I9_) | 22224 | 191924 |
| APPENDACUT_NOCOMPLIC | Acute appendicitis, no complications | XI Diseases of the digestive system (K11_) | 21998 | 346283 |
| AUD_SWEDISH | Alcohol use disorder, Swedish definition | Alcohol related diseases | 21996 | 355281 |
| L12_DERMATITISNAS | Other and unspecified dermatitis | XII Diseases of the skin and subcutaneous tissue (L12_) | 21951 | 336589 |
| ASTHMA_INFECTIONS | Asthma-related infections | Asthma and related endpoints | 21927 | 335114 |
| I9_AF_REIMB | Atrial fibrillation and flutter with reimbursement | IX Diseases of the circulatory system (I9_) | 21700 | 191924 |
| N14_MENORRHAGIA | Menorrhagia | XIV Diseases of the genitourinary system (N14_) | 21496 | 107564 |
| ASTHMA_NAS | Asthma, unspecified (mode) | Asthma and related endpoints | 21392 | 210122 |
| E4_OBESITY | Obesity | IV Endocrine, nutritional and metabolic diseases (E4_) | 21375 | 355786 |
| L12_INFECT_SKIN | Infections of the skin and subcutaneous tissue | XII Diseases of the skin and subcutaneous tissue (L12_) | 21308 | 355969 |
| H7_ALLERGICCONJUNCTIVITIS | Allergic conjunctivitis | VII Diseases of the eye and adnexa (H7_) | 20958 | 356319 |
| C3_SKIN_EXALLC | Malignant neoplasm of skin (controls excluding all cancers) | II Neoplasms, from cancer register (ICD-O-3) | 20951 | 287137 |

| | | | | |
|---|---|---|---|---|
| M13_ARTHROSIS_OTH | Other arthrosis | XIII Diseases of the musculoskeletal system and connective tissue (M13_) | 20949 | 240862 |
| I9_VHD | Valvular heart disease excluding rheumatic fever | IX Diseases of the circulatory system (I9_) | 20929 | 286109 |
| J10_LOWERINF | Other acute lower respiratory infections | X Diseases of the respiratory system (J10_) | 20873 | 356404 |
| I9_NONRHEVALV | Non-rheumatic valve diseases | IX Diseases of the circulatory system (I9_) | 20772 | 286109 |
| N14_OVARYCYST | Ovarian cyst | XIV Diseases of the genitourinary system (N14_) | 20750 | 107564 |
| K11_REIMB_202 | KELA_REIMBURSEMENT_202 | Gastrointestinal endpoints | 20730 | 356547 |
| J10_SINUSITIS | Acute sinusitis | X Diseases of the respiratory system (J10_) | 20520 | 308166 |
| M13_SYNOTEND | Disorders of synovium and tendon | XIII Diseases of the musculoskeletal system and connective tissue (M13_) | 20492 | 275212 |
| AB1_ERYSIPELAS | Erysipelas | I Certain infectious and parasitic diseases (AB1_) | 20470 | 332343 |
| RX_CROHN_2NDLINE | Second line medication for Crohn's disease | Drug purchase endpoints | 20353 | 356924 |
| CD2_BENIGN_COLORECANI | Benign neoplasm of colon, rectum, anus and anal canal | II Neoplasms from hospital discharges (CD2_) | 20336 | 356941 |
| M13_ARTHTROSIS_COX_PRIM_ICD10 | Coxarthrosis, primary | XIII Diseases of the musculoskeletal system and connective tissue (M13_) | 20056 | 357221 |
| ST19_FRACT_LOWER_LEG_INCLU_ANKLE | Fracture of lower leg, including ankle | XIX Injury, poisoning and certain other consequences of external causes (ST19_) | 19994 | 323307 |
| M13_SPONDYLOSIS | Spondylosis | XIII Diseases of the musculoskeletal system and connective tissue (M13_) | 19892 | 270964 |
| ST19_FRACT_FOREA | Fracture of forearm | XIX Injury, poisoning and certain other consequences of external causes (ST19_) | 19577 | 351196 |
| J10_VOCALLARYNX | Diseases of vocal cords and larynx+other diseases of upper respiratory tract, no elsewhere classified | X Diseases of the respiratory system (J10_) | 19530 | 283342 |
| C3_BASAL_CELL_CARCINOMA_INCLAVO_EXALLC | Basal cell carcinoma, including avohilmo (controls excluding all cancers) | II Neoplasms, from cancer register (ICD-O-3) | 19522 | 286768 |
| H7_SCLERACORNEA | Disorders of sclera, cornea, iris and ciliary body | VII Diseases of the eye and adnexa (H7_) | 19463 | 357814 |
| I9_VTE | Venous thromboembolism | IX Diseases of the circulatory system (I9_) | 19372 | 357905 |
| I9_K_CARDIAC | Death due to cardiac causes | IX Diseases of the circulatory system (I9_) | 19295 | 357982 |
| M13_OSTEOCHONDRO | Osteopathies and chondropathies | XIII Diseases of the musculoskeletal system and connective tissue (M13_) | 19263 | 358014 |
| M13_ARTHROSIS_KNEE_PRIM_KNEESURG | Gonarthrosis, primary, with knee surgery | XIII Diseases of the musculoskeletal system and connective tissue (M13_) | 19126 | 337196 |
| M13_SPINSTENOSIS | Spinal stenosis | XIII Diseases of the musculoskeletal system and connective tissue (M13_) | 18989 | 270964 |
| C3_BASAL_CELL_CARCINOMA_EXALLC | Basal cell carcinoma (controls excluding all cancers) | II Neoplasms, from cancer register (ICD-O-3) | 18982 | 287137 |
| H7_GLAUCOMA | Glaucoma | VII Diseases of the eye and adnexa (H7_) | 18902 | 358375 |
| AB1_BACTINF_NOS | Bacterial infection, other or unspecified | I Certain infectious and parasitic diseases (AB1_) | 18576 | 332343 |
| M13_SCIATICA | Sciatica+with lumbago | XIII Diseases of the musculoskeletal system and connective tissue (M13_) | 18569 | 270964 |
| G6_MIGRAINE | Migraine | VI Diseases of the nervous system (G6_) | 18477 | 287837 |
| I9_TIA | Transient ischemic attack | IX Diseases of the circulatory system (I9_) | 18398 | 342294 |
| RX_GLUCOSAMINE | Glucosamine medication | Drug purchase endpoints | 18341 | 358936 |
| J10_COPD | COPD | X Diseases of the respiratory system (J10_) | 18266 | 311286 |
| N14_CYSTITIS | Cystitis | XIV Diseases of the genitourinary system (N14_) | 18080 | 328001 |
| C3_OTHER_SKIN_EXALLC | Other malignant neoplasms of skin (=non-melanoma skin cancer) (controls excluding all cancers) | II Neoplasms, from cancer register (ICD-O-3) | 17958 | 287137 |
| F5_DEMENTIA_INCLAVO | Dementia, including avohilmo | V Mental and behavioural disorders (F5_) | 17901 | 359376 |
| H7_CATARACTOTHER | Other cataract | VII Diseases of the eye and adnexa (H7_) | 17699 | 312864 |
| K11_ENERCOLNONINF | Noninfective enteritis and colitis | XI Diseases of the digestive system (K11_) | 17350 | 359927 |
| N14_MENSIRREG | Irregular menses | XIV Diseases of the genitourinary system (N14_) | 17228 | 179322 |
| F5_DEPRESSION_RECURRENT | Recurrent or chronic depression | V Mental and behavioural disorders (F5_) | 17227 | 228647 |
| N14_FEMGENPROL | Female genital prolapse | XIV Diseases of the genitourinary system (N14_) | 17150 | 107564 |
| PRIM_KNEEARTHROSIS | Primary gonarthrosis, bilateral | Rheuma endpoints | 17009 | 332589 |
| O15_ABORT_SPONTAN | Spontaneous abortion | XV Pregnancy, childbirth and the puerperium (O15_) | 16906 | 149622 |
| N14_BREAST | Disorders of breast | XIV Diseases of the genitourinary system (N14_) | 16897 | 193973 |
| F5_ANXIETY | Other anxiety disorders | V Mental and behavioural disorders (F5_) | 16887 | 337577 |
| I9_HEARTFAIL_AND_OVERWEIGHT | Heart failure and bmi 25plus | Cardiometabolic endpoints | 16707 | 154410 |
| F5_DEMENTIA | Dementia | V Mental and behavioural disorders (F5_) | 16499 | 356660 |
| CD2_BENIGN_COLON | Benign neoplasm: Colon | II Neoplasms from hospital discharges (CD2_) | 16445 | 360832 |
| J10_CHRONSINUSITIS | Chronic sinusitis | X Diseases of the respiratory system (J10_) | 16395 | 283342 |
| DM_SEVERAL_COMPLICATIONS | Diabetes, several complications | Diabetes endpoints | 16320 | 271817 |
| K11_OTHFUNC | Other functional intestinal disorders | XI Diseases of the digestive system (K11_) | 16311 | 301931 |
| J10_PNEUMOBACT | Bacterial pneumoniae | X Diseases of the respiratory system (J10_) | 16244 | 314673 |
| K11_OTHDIG | Other diseases of the digestive system | XI Diseases of the digestive system (K11_) | 16222 | 361055 |
| L12_DISORDSKINAPPENDIX | Disorders of skin appendages | XII Diseases of the skin and subcutaneous tissue (L12_) | 16137 | 361140 |
| H7_OCUMUSCLE | Disorders of ocular muscles, binocular movement, accommodation and refraction | VII Diseases of the eye and adnexa (H7_) | 16040 | 361237 |
| ASTHMA_ACUTE_RESPIRATORY_INFECTIONS | Asthma-related acute respiratory infections | Asthma and related endpoints | 16018 | 335114 |
| G6_HEADACHE | Other headache syndromes | VI Diseases of the nervous system (G6_) | 15851 | 287837 |
| AUD | Alcohol use disorder, ICD-based | V Mental and behavioural disorders (F5_) | 15715 | 361562 |
| H7_VISUDISTURB | Visual disturbances | VII Diseases of the eye and adnexa (H7_) | 15707 | 360091 |
| C3_BREAST_EXALLC | Malignant neoplasm of breast (controls excluding all cancers) | II Neoplasms, from cancer register (ICD-O-3) | 15680 | 167189 |
| F5_KELAMENT | Kela-code for severe mental illness | V Mental and behavioural disorders (F5_) | 15451 | 360114 |
| J10_PLEURA | Other diseases of pleura | X Diseases of the respiratory system (J10_) | 15441 | 361836 |
| J10_BRONCHITIS | Acute bronchitis | X Diseases of the respiratory system (J10_) | 15393 | 356404 |
| ABDOM_HERNIA | Hernia of abdominal wall | XI Diseases of the digestive system (K11_) | 15338 | 361939 |
| E4_ENDOGLAND | Disorders of other endocrine glands | IV Endocrine, nutritional and metabolic diseases (E4_) | 15289 | 361988 |
| H7_VITRBODYGLOBE | Disorders of vitreous body | VII Diseases of the eye and adnexa (H7_) | 15177 | 361552 |
| H7_MACULADEGEN | Degeneration of macula and posterior pole | VII Diseases of the eye and adnexa (H7_) | 15137 | 344569 |
| N14_ENDOMETRIOSIS | Endometriosis | XIV Diseases of the genitourinary system (N14_) | 15088 | 107564 |
| E4_OBESITYCAL | Obesity due to excess calories | IV Endocrine, nutritional and metabolic diseases (E4_) | 15045 | 355902 |
| I9_ATHSCLE | Atherosclerosis, excluding cerebral, coronary and PAD | IX Diseases of the circulatory system (I9_) | 15002 | 349539 |
| M13_ARTHTROSIS_COX_PRIM_HIPSURG | Coxarthrosis, primary, with hip surgery | XIII Diseases of the musculoskeletal system and connective tissue (M13_) | 14990 | 357221 |
| H8_VERTIGO | Disorders of vestibular function (Vertigo) | VIII Diseases of the ear and mastoid process (H8_) | 14918 | 359094 |
| O15_HYPTENSPREG | Pregnancy hypertension | XV Pregnancy, childbirth and the puerperium (O15_) | 14727 | 196143 |
| M13_HALLUXVALGUS | Hallux valgus (acquired) | XIII Diseases of the musculoskeletal system and connective tissue (M13_) | 14726 | 240862 |
| I9_ANGIO | Coronary angioplasty | IX Diseases of the circulatory system (I9_) | 14723 | 313400 |
| H8_MED_SUPP | Suppurative and unspecified otitis media | VIII Diseases of the ear and mastoid process (H8_) | 14406 | 352571 |
| KRA_PSY_DEMENTIA_EXMORE | Any dementia (more control exclusions) | Psychiatric endpoints from Katri Räikkönen | 14367 | 277526 |

| | | | | |
|---|---|---|---|---|
| H7_EYELIDDIS | Other disorders of eyelid | VII Diseases of the eye and adnexa (H7_) | 14347 | 344684 |
| N14_POLYPFEMGEN | Polyp of the female genital tract | XIV Diseases of the genitourinary system (N14_) | 14324 | 107564 |
| O15_FALSE_LAB | False labour | XV Pregnancy, childbirth and the puerperium (O15_) | 14282 | 168929 |
| AB1_VIRAL_SKIN_MUCOUS_MEMBRANE | Viral infections characterized by skin and mucous membrane lesions | I Certain infectious and parasitic diseases (AB1_) | 14280 | 362997 |
| N14_RENFAIL | Renal failure | XIV Diseases of the genitourinary system (N14_) | 14100 | 363177 |
| M13_DORSALGIANAS | Other/unspecified dorsalgia | XIII Diseases of the musculoskeletal system and connective tissue (M13_) | 14075 | 270964 |
| F5_STRESSOTH | Other reaction to severe stress, and adjustment disorders | V Mental and behavioural disorders (F5_) | 13913 | 337577 |
| D3_ANAEMIA_IRONDEF | Iron deficiency anaemia | III Diseases of the blood and blood-forming organs and certain disorders involving the immune mechanism (D3_) | 13689 | 360528 |
| D3_ANAEMIANAS | Other and unspecified anaemias | III Diseases of the blood and blood-forming organs and certain disorders involving the immune mechanism (D3_) | 13600 | 362319 |
| L12_ATOPIC | Atopic dermatitis | XII Diseases of the skin and subcutaneous tissue (L12_) | 13473 | 336589 |
| M13_CERVICDISC | Cervical disc disorders | XIII Diseases of the musculoskeletal system and connective tissue (M13_) | 13394 | 270964 |
| G6_AD_WIDE | Alzheimer’s disease, wide definition | VI Diseases of the nervous system (G6_) | 13393 | 363884 |
| H7_LACRIMALSYSTEM | Disorders of lacrimal system | VII Diseases of the eye and adnexa (H7_) | 13326 | 344684 |
| I9_UAP | Unstable angina pectoris | IX Diseases of the circulatory system (I9_) | 13304 | 333375 |
| TEMPOROMANDIB_INCLAVO | Temporomandibular joint disorders, including avohilmo | XI Diseases of the digestive system (K11_) | 13282 | 363995 |
| C3_PROSTATE_EXALLC | Malignant neoplasm of prostate (controls excluding all cancers) | II Neoplasms, from cancer register (ICD-O-3) | 13216 | 119948 |
| L12_PAPULOSQUAMOUS | Papulosquamous disorders | XII Diseases of the skin and subcutaneous tissue (L12_) | 13206 | 364071 |
| J10_TONSILLITIS | Other and unspecified tonsillitis | X Diseases of the respiratory system (J10_) | 13190 | 308166 |
| N14_FEMALEINFERT | Female infertility | XIV Diseases of the genitourinary system (N14_) | 13142 | 107564 |
| NEURODEGOTH | Other degenerative diseases of the nervous system | Comorbidities of Gastrointestinal endpoints | 13086 | 212179 |
| KRA_PSY_SCHIZODEL_EXMORE | Schizophrenia or delusion (more control exclusions) | Psychiatric endpoints from Katri Räikkönen | 13061 | 277526 |
| GEST_DIABETES | Gestational diabetes (for exclusion) | Diabetes endpoints | 13039 | 197831 |
| I9_HEARTFAIL_AND_CHD | Heart failure and coronary heart disease | Cardiometabolic endpoints | 12872 | 333759 |
| I9_VEINSOTH | Other disorders of veins | IX Diseases of the circulatory system (I9_) | 12630 | 324121 |
| O15_DELIV_CAESAR | Single delivery by caesarean section | XV Pregnancy, childbirth and the puerperium (O15_) | 12596 | 92306 |
| M13_RHEUMA | Rheumatoid arthritis | XIII Diseases of the musculoskeletal system and connective tissue (M13_) | 12555 | 240862 |
| CD2_BENIGN_SKIN | Other benign neoplasms of skin | II Neoplasms from hospital discharges (CD2_) | 12341 | 364936 |
| E4_HYPERLIPNAS | Hyperlipidaemia, other/unspecified | IV Endocrine, nutritional and metabolic diseases (E4_) | 12304 | 324150 |
| AB1_OTHER_SEPSIS | Other septicaemia | I Certain infectious and parasitic diseases (AB1_) | 12301 | 332343 |
| O15_OTHER_MATERN_DIS_ELSEWHERE | Other maternal diseases classifiable elsewhere but complicating pregnancy, childbirth and the puerperium | XV Pregnancy, childbirth and the puerperium (O15_) | 12297 | 198180 |
| K11_PULP_PERIAPICAL | Diseases of pulp and periapical tissues | XI Diseases of the digestive system (K11_) | 12078 | 259234 |
| ASTHMA_PNEUMONIA | Asthma-related pneumonia | Asthma and related endpoints | 12044 | 335114 |
| HYPOTHY_REIMB | Hypothyroidism, drug reimbursement | IV Endocrine, nutritional and metabolic diseases (E4_) | 12040 | 88531 |
| H7_VITROTH | Other and unspecified disorders of vitreous body | VII Diseases of the eye and adnexa (H7_) | 12004 | 361552 |
| K11_DIAHER | Diaphragmatic hernia | XI Diseases of the digestive system (K11_) | 11757 | 322586 |
| ST19_FRACT_SHOUL_UPPER_ARM | Fracture of shoulder and upper arm | XIX Injury, poisoning and certain other consequences of external causes (ST19_) | 11750 | 345173 |
| M13_SYSTCONNECT | Systemic connective tissue disorders | XIII Diseases of the musculoskeletal system and connective tissue (M13_) | 11744 | 365533 |
| G6_EPLEPSY | Epilepsy | VI Diseases of the nervous system (G6_) | 11740 | 287837 |
| O15_CONCEPT_ABNORM | Other abnormal products of conception | XV Pregnancy, childbirth and the puerperium (O15_) | 11716 | 149622 |
| J10_SEPTDEV | Deviated nasal septum | X Diseases of the respiratory system (J10_) | 11680 | 283342 |
| Z21_PROCREATIVE_MANAG | Procreative management | XXI Factors influencing health status and contact with health services (Z21_) | 11666 | 300151 |
| R18_RETEN_URINE | Retention of urine | XVIII Symptoms, signs and abnormal clinical and laboratory findings, not elsewhere classified (R18_) | 11657 | 343006 |
| H7_KERATITIS | Keratitis | VII Diseases of the eye and adnexa (H7_) | 11611 | 357814 |
| N14_POSTMENBLEED | Postmenopausal bleeding | XIV Diseases of the genitourinary system (N14_) | 11582 | 107564 |
| I9_SEQULAE | Sequelae of cerebrovascular disease | IX Diseases of the circulatory system (I9_) | 11408 | 342673 |
| ST19_FRACT_WRIST_HAND_LEVEL | Fracture at wrist and hand level | XIX Injury, poisoning and certain other consequences of external causes (ST19_) | 11394 | 336418 |
| N14_FIOTHNAS | Female infertility, cervigal, vaginal, other or unspecified origin | XIV Diseases of the genitourinary system (N14_) | 11348 | 107564 |
| H7_RETINALDETACH | Retinal detachments and breaks | VII Diseases of the eye and adnexa (H7_) | 11230 | 344569 |
| L12_ACTINKERA | Actinic keratosis | XII Diseases of the skin and subcutaneous tissue (L12_) | 11081 | 364831 |
| RHEU_ARTHRITIS_OTH | Other arthritis (FG) | Rheuma endpoints | 11033 | 285035 |
| E4_OBESITYNAS | Obesity, other/unspecified | IV Endocrine, nutritional and metabolic diseases (E4_) | 11025 | 355902 |
| ALLERG_RHINITIS | Allergic rhinitis | Comorbidities of Asthma | 11009 | 359149 |
| I9_CABG | Coronary artery bypass grafting | IX Diseases of the circulatory system (I9_) | 10938 | 313400 |
| AB1_SEXUAL_TRANSMISSION | Infections with a predominantly sexual mode of transmission | I Certain infectious and parasitic diseases (AB1_) | 10937 | 366340 |
| J10_CHRONRHINITIS | Chronic rhinitis, nasopharyngitis and pharyngitis | X Diseases of the respiratory system (J10_) | 10868 | 283342 |
| K11_LIVER | Diseases of liver | XI Diseases of the digestive system (K11_) | 10827 | 366450 |
| KRA_PSY_ANYMENTAL_SUICID_PREG_NERV_EXMORE | Any mental disorder, or suicide (or attempt), or psychic disorders complicating pregnancy, partum or puerperium or nerve system disorders (more control exclusions) | Psychiatric endpoints from Katri Räikkönen | 10811 | 276857 |
| H7_GLAUCSUSP | Glaucoma suspect | VII Diseases of the eye and adnexa (H7_) | 10758 | 358375 |
| K11_OTHDISDIG | Other diseases of the digestive system | XI Diseases of the digestive system (K11_) | 10589 | 361055 |
| M13_ENTESOPATHYOTH | Other enthesopathies | XIII Diseases of the musculoskeletal system and connective tissue (M13_) | 10581 | 275212 |
| DM_RETINOPATHY_EXMORE | Diabetic retinopathy (more control exclusions) | Diabetes endpoints | 10413 | 308633 |
| COPD_LATER | Later onset COPD | COPD and related endpoints | 10404 | 161813 |
| F5_BEHAVE | Behavioural syndromes associated with physiological disturbances and physical factors | V Mental and behavioural disorders (F5_) | 10401 | 366876 |
| N14_ABNORMALBLEED | Other abnormal uterine and caginal bleeding | XIV Diseases of the genitourinary system (N14_) | 10319 | 107564 |
| ASTHMA_OBESITY | Obesity related asthma | Comorbidities of Asthma | 10306 | 335114 |
| DENTAL_TMD | TMD related pain | Dental endpoints | 10303 | 366974 |
| O15_PREEC_OR_FETGRO | Pre-eclampsia or poor fetal growth | XV Pregnancy, childbirth and the puerperium (O15_) | 10297 | 200573 |
| H8_EXTERNAL | Diseases of external ear | VIII Diseases of the ear and mastoid process (H8_) | 10272 | 367005 |
| D3_ANAEMIA_IRONDEF_NAS | Other and unspecified iron deficiency | III Diseases of the blood and blood-forming organs and certain disorders involving the immune mechanism (D3_) | 10208 | 360528 |
| L12_URTICARIA | Urticaria | XII Diseases of the skin and subcutaneous tissue (L12_) | 10175 | 364583 |
| KRA_PSY_PERSON_EXMORE | Personality disorders (more control exclusions) | Psychiatric endpoints from Katri Räikkönen | 10012 | 277522 |
| I9_CONDUCTIO | Conduction disorders | IX Diseases of the circulatory system (I9_) | 9949 | 286109 |
| K11_OTHANRECT | Other diseases of anus and rectum | XI Diseases of the digestive system (K11_) | 9932 | 301931 |

| | | | | |
|---|---|---|---|---|
| I9_NONISCHCARDMYOP | Non-ischemic cardiomyopathy | IX Diseases of the circulatory system (I9_) | 9926 | 303607 |
| F5_ALCOHOL_DEPENDENCE | Alcohol dependence | V Mental and behavioural disorders (F5_) | 9876 | 353355 |
| K11_DENTOFACIAL_ANOMALIES | Dentofacial anomalies [including malocclusion] | XI Diseases of the digestive system (K11_) | 9866 | 259234 |
| H8_SUP_ACUTE | Acute suppurative otitis media | VIII Diseases of the ear and mastoid process (H8_) | 9850 | 352571 |
| G6_XTRAPYR | Extrapyramidal and movement disorders | VI Diseases of the nervous system (G6_) | 9833 | 367444 |
| AB1_OTHER_VIRAL | Other viral diseases | I Certain infectious and parasitic diseases (AB1_) | 9805 | 367472 |
| O15_MATERN_CARE_OTHER | Maternal care for other known or suspected fetal problems | XV Pregnancy, childbirth and the puerperium (O15_) | 9783 | 168929 |
| N14_CALCUKIDUR | Calculus of kidney and ureter | XIV Diseases of the genitourinary system (N14_) | 9713 | 366693 |
| C3_BREAST_ERPLUS_EXALLC | Malignant neoplasm of breast, HER-positive (controls excluding all cancers) | II Neoplasms, from cancer register (ICD-O-3) | 9698 | 167017 |
| E4_FLUIDELECTRO | Other disorders of fluid, electrolyte and acid-base balance | IV Endocrine, nutritional and metabolic diseases (E4_) | 9694 | 324150 |
| M13_DEFORMDORSO | Deforming dorsopathies | XIII Diseases of the musculoskeletal system and connective tissue (M13_) | 9672 | 270964 |
| ALLERG_ASTHMA | Allergic asthma (mode) | Asthma and related endpoints | 9631 | 210122 |
| I9_PAROXTAC | Paroxysmal tachycardia | IX Diseases of the circulatory system (I9_) | 9604 | 191924 |
| K11_CHRONGASTR | Chronic gastritis | XI Diseases of the digestive system (K11_) | 9570 | 320387 |
| DM_RETINA_PROLIF | Proliferative diabetic retinopathy | Diabetes endpoints | 9511 | 362581 |
| E4_NONTOXIC_THYROID | Nontoxic goitre/thyroid nodule | IV Endocrine, nutritional and metabolic diseases (E4_) | 9485 | 367792 |
| O15_HAEMORRH_EARLY_PREG | Haemorrhage in early pregnancy | XV Pregnancy, childbirth and the puerperium (O15_) | 9433 | 179899 |
| CD2_BENIGN_LIPOMATOUS | Benign lipomatous neoplasm | II Neoplasms from hospital discharges (CD2_) | 9431 | 367846 |
| K11_IBS | Irritable bowel syndrome | XI Diseases of the digestive system (K11_) | 9323 | 301931 |
| G6_ALZHEIMER | Alzheimer disease | VI Diseases of the nervous system (G6_) | 9301 | 367976 |
| L12_PSORIASIS | Psoriasis | XII Diseases of the skin and subcutaneous tissue (L12_) | 9267 | 364071 |
| I9_PULMEMB | Pulmonary embolism | IX Diseases of the circulatory system (I9_) | 9243 | 367108 |
| RHEUMA_SEROPOS_WIDE | Seropositive rheumatoid arthritis, wide | Rheuma endpoints | 9243 | 368029 |
| N14_FEMALE_GENITAL_DYSPLASIA_ALL | All dysplastic lesions of the cervix uteri, vagina or vulva | XIV Diseases of the genitourinary system (N14_) | 9227 | 201643 |
| K11_GASTRODUOULC | Gastroduodenal ulcer | XI Diseases of the digestive system (K11_) | 9216 | 320387 |
| I9_CAVS_OPERATED | Calcific aortic valvular stenosis, operated | IX Diseases of the circulatory system (I9_) | 9153 | 368124 |
| O15_MATERN_CARE_PELVIC_ABNORM | Maternal care for known or suspected abnormality of pelvic organs | XV Pregnancy, childbirth and the puerperium (O15_) | 9149 | 168929 |
| RHEUMA_SEROPOS_OTH | Other/unspecified seropositive rheumatoid arthritis | Rheuma endpoints | 9139 | 368029 |
| I9_PHLETHROMBDVTLOW | DVT of lower extremities | IX Diseases of the circulatory system (I9_) | 9109 | 324121 |
| ST19_FRACT_RIBS_STERNUM_THORACIC_SPINE | Fracture of rib(s), sternum and thoracic spine | XIX Injury, poisoning and certain other consequences of external causes (ST19_) | 9094 | 362260 |
| N14_CHRONKIDNEYDIS | Chronic kidney disease | XIV Diseases of the genitourinary system (N14_) | 9073 | 363177 |
| DENTAL_TMD_FIBRO | TMD muscular pain linked with fibromyalgia | Dental endpoints | 8995 | 368282 |
| VWXY20_SUICI_OTHER_INTENTI_SELF_H | Suicide or other Intentional self-harm | Psychiatric endpoints from Katri Räikkönen | 8978 | 368299 |
| T1D_WIDE | Type 1 diabetes, wide definition | Diabetes endpoints | 8967 | 308373 |
| F5_ALCOHOLAC | Acute alcohol intoxication | V Mental and behavioural disorders (F5_) | 8957 | 353355 |
| H7_AMD | Age-related macular degeneration (whether dry or wet) | VII Diseases of the eye and adnexa (H7_) | 8913 | 348936 |
| H7_CONJUNCTIVITISACUNONATOPIC | Conjunctivitis (acute, non atopic) | VII Diseases of the eye and adnexa (H7_) | 8907 | 345122 |
| CD2_BENIGN_MELANOCYTIC | Melanocytic naevi | II Neoplasms from hospital discharges (CD2_) | 8900 | 368377 |
| K11_FISTULA | Colonic or urogenital fistula | XI Diseases of the digestive system (K11_) | 8879 | 368398 |
| K11_FUNCDYSP | Functional dyspepsia | XI Diseases of the digestive system (K11_) | 8875 | 320387 |
| H8_OTHHEARINGLOSS | Other hearing loss | VIII Diseases of the ear and mastoid process (H8_) | 8833 | 331736 |
| ST19_FRACT_FEMUR | Fracture of femur | XIX Injury, poisoning and certain other consequences of external causes (ST19_) | 8766 | 360928 |
| N14_OTHNONINFUTER | Other noninflammatory disorders of uterus, except cervix | XIV Diseases of the genitourinary system (N14_) | 8752 | 107564 |
| OTHER_SYSTCON_FG | Other systemic involvement of connective tissue (FG) | Rheuma endpoints | 8695 | 285035 |
| M13_IMPINGEMENT | Impingement syndrome of shoulder | XIII Diseases of the musculoskeletal system and connective tissue (M13_) | 8676 | 275212 |
| K11_ILEUS | Paralytic ileus and intestinal obstruction | XI Diseases of the digestive system (K11_) | 8660 | 301931 |
| O15_COMPLIC_PUERP | Complications predominantly related to the puerperium | XV Pregnancy, childbirth and the puerperium (O15_) | 8603 | 202267 |
| PRIM_COXARTHROSIS | Primary coxarthrosis, bilateral | Rheuma endpoints | 8597 | 355023 |
| K11_HAEMORR | Haemmorrhoids and perianal venous thrombosis | XI Diseases of the digestive system (K11_) | 8583 | 301931 |
| J10_INFLUENZA | All influenza | X Diseases of the respiratory system (J10_) | 8555 | 314673 |
| H7_BLEPHAROCHALASIS | Blepharochalasis | VII Diseases of the eye and adnexa (H7_) | 8547 | 344684 |
| K11_UMBHER | Umbilical hernia | XI Diseases of the digestive system (K11_) | 8510 | 322586 |
| O15_PRETERM | Preterm labour and delivery | XV Pregnancy, childbirth and the puerperium (O15_) | 8507 | 162777 |
| O15_GESTAT_HYPERT | Gestational [pregnancy-induced] hypertension | XV Pregnancy, childbirth and the puerperium (O15_) | 8502 | 194266 |
| M13_GOUT | Gout | XIII Diseases of the musculoskeletal system and connective tissue (M13_) | 8489 | 240862 |
| I9_HYPTENSHD | Hypertensive Heart Disease | IX Diseases of the circulatory system (I9_) | 8387 | 265626 |
| M13_EARLY_LUMBAR_PROLAPSE | Early lumbar disc prolapse, operated | XIII Diseases of the musculoskeletal system and connective tissue (M13_) | 8351 | 368926 |
| O15_LABOUR_FETAL_STRESS | Labour and delivery complicated by fetal stress [distress] | XV Pregnancy, childbirth and the puerperium (O15_) | 8324 | 162777 |
| H8_NONSUPPNAS | Nonsuppurative otitis media | VIII Diseases of the ear and mastoid process (H8_) | 8315 | 352571 |
| H8_BPV | Benign paroxysmal vertigo | VIII Diseases of the ear and mastoid process (H8_) | 8280 | 359094 |
| O15_POSTPART_HEAMORRH | Postpartum haemorrhage | XV Pregnancy, childbirth and the puerperium (O15_) | 8249 | 162777 |
| O15_POSTPART_HEAMORRH_ALLW | Postpartum haemorrhage | XV Pregnancy, childbirth and the puerperium (O15_) | 8249 | 202621 |
| O15_POSTPART_HEAMORRH_LB | Postpartum haemorrhage | XV Pregnancy, childbirth and the puerperium (O15_) | 8249 | 146016 |
| M13_SOFTOVERUSE | Soft tissue disorders related to use, overuse and pressure | XIII Diseases of the musculoskeletal system and connective tissue (M13_) | 8236 | 275212 |
| AB1_MYCOSES | Mycoses | I Certain infectious and parasitic diseases (AB1_) | 8179 | 369098 |
| THYROTOXICOSIS | Thyrotoxicosis | Comorbidities of Gastrointestinal endpoints | 8173 | 367407 |
| L12_ABSCESS_CUT | Cutaneous abscess, furuncle and carbuncle | XII Diseases of the skin and subcutaneous tissue (L12_) | 8157 | 355969 |
| K11_EMBIMPACT_TEETH_INCLAVO | Embedded and impacted teeth , including avohilmo | XI Diseases of the digestive system (K11_) | 8139 | 369138 |
| N14_CERVICAL_DYSPLASIA_ALL | All dysplastic lesions of the cervix uteri | XIV Diseases of the genitourinary system (N14_) | 8025 | 202845 |
| K11_OTHENTERCOL | Other noninfective gastroenteritis and colitis | XI Diseases of the digestive system (K11_) | 7988 | 359927 |

| | | | | |
|---|---|---|---|---|
| M13_FIBROBLASTIC | Fibroblastic disorders | XIII Diseases of the musculoskeletal system and connective tissue (M13_) | 7952 | 275212 |
| G6_MIGRAINE_WITH_AURA | Migraine with aura | VI Diseases of the nervous system (G6_) | 7917 | 287837 |
| O15_LABOUR_LONG | Long labour | XV Pregnancy, childbirth and the puerperium (O15_) | 7866 | 162777 |
| DM_KETOACIDOSIS | Diabetic ketoacidosis | Diabetes endpoints | 7841 | 271817 |
| H7_GLAUCOMA_POAG | Primary open-angle glaucoma, strict | VII Diseases of the eye and adnexa (H7_) | 7756 | 358375 |
| H7_REFRAACCOMMODIS | Disorders of refraction and accommodation | VII Diseases of the eye and adnexa (H7_) | 7737 | 361237 |
| K11_CARIES_DENTIN | Dentinal caries | XI Diseases of the digestive system (K11_) | 7680 | 259234 |
| O15_DELIV_FORCEP_VAC | Single delivery by forceps and vacuum extractor | XV Pregnancy, childbirth and the puerperium (O15_) | 7630 | 92306 |
| K11_VENTHER | Ventral hernia | XI Diseases of the digestive system (K11_) | 7628 | 322586 |
| K11_IBD_STRICT | Inflammatory bowel disease, strict (require KELA) | XI Diseases of the digestive system (K11_) | 7625 | 369652 |
| K11_KELAIBD | IBD patients in KELA-register | XI Diseases of the digestive system (K11_) | 7625 | 359927 |
| AB1_VIRAL_OTHER_INTEST_INFECTIONS | Viral and other specified intestinal infections | I Certain infectious and parasitic diseases (AB1_) | 7613 | 336396 |
| ST19_FRACT_FOOT_ANKLE | Fracture of foot, except ankle | XIX Injury, poisoning and certain other consequences of external causes (ST19_) | 7593 | 351392 |
| N14_ENDOMETRIOSIS_ASRM_STAGE3_4 | Endometriosis ASRM stages 3,4 | XIV Diseases of the genitourinary system (N14_) | 7574 | 203296 |
| CD2_PRIMARY_LYMPHOID_HEMATOPOIETIC_EXALLC | Primary_lymphoid and hematopoietic malignant neoplasms (controls excluding all cancers) | II Neoplasms from hospital discharges (CD2_) | 7519 | 299952 |
| K11_OTHGASTR | Other gastritis (incl. Duodenitis) | XI Diseases of the digestive system (K11_) | 7514 | 320387 |
| J10_PERITONSABSC | Peritonsillar abscess | X Diseases of the respiratory system (J10_) | 7510 | 283342 |
| F5_PSYCHOTH | Other and unspecified nonorganic psychotic disorders | V Mental and behavioural disorders (F5_) | 7508 | 364160 |
| E4_DMNAS | Unspecified diabetes | IV Endocrine, nutritional and metabolic diseases (E4_) | 7455 | 308280 |
| O15_LABOUR_MALPOS | Obstructed labour due to malposition and malpresentation of fetus | XV Pregnancy, childbirth and the puerperium (O15_) | 7414 | 162777 |
| I9_AORTANEUR | Aortic aneurysm | IX Diseases of the circulatory system (I9_) | 7395 | 349539 |
| M13_CERVICOBRACHSDR | Cervicobrachial syndrome | XIII Diseases of the musculoskeletal system and connective tissue (M13_) | 7353 | 270964 |
| DM_HYPOGLYC | Diabetic hypoglycemia | Diabetes endpoints | 7332 | 271817 |
| H7_VISUDISTURBSUB | Subjective visual disturbances | VII Diseases of the eye and adnexa (H7_) | 7313 | 360091 |
| H7_IRIDOCYCLITIS | Iridocyclitis | VII Diseases of the eye and adnexa (H7_) | 7306 | 357814 |
| M13_OSTEOPOROSIS | Osteoporosis | XIII Diseases of the musculoskeletal system and connective tissue (M13_) | 7300 | 358014 |
| M13_GANGLION | Ganglion | XIII Diseases of the musculoskeletal system and connective tissue (M13_) | 7270 | 275212 |
| CPAP | Continuous positive airway pressure | Interstitial lung disease endpoints | 7247 | 164315 |
| O15_PRE_OR_ECLAMPSIA | Pre-eclampsia or eclampsia | XV Pregnancy, childbirth and the puerperium (O15_) | 7212 | 194266 |
| H7_STRABOTH | Other strabismus | VII Diseases of the eye and adnexa (H7_) | 7179 | 361237 |
| H8_TINNITUS | Tinnitus | VIII Diseases of the ear and mastoid process (H8_) | 7175 | 331736 |
| O15_MEMBR_PREMAT_RUPT | Premature rupture of membranes | XV Pregnancy, childbirth and the puerperium (O15_) | 7147 | 168929 |
| NONALLERG_ASTHMA_EXMORE | Non-allergic asthma (mode) (more control exclusions) | Asthma and related endpoints | 7143 | 202399 |
| M13_ENTESOPATHYLOW | Enthesopathies of lower limb, excluding foot | XIII Diseases of the musculoskeletal system and connective tissue (M13_) | 7125 | 270964 |
| COPD_EARLY | Early onset COPD | COPD and related endpoints | 7079 | 359794 |
| I9_PHLETHROM | Phlebitis and thrombophlebitis (not including DVT) | IX Diseases of the circulatory system (I9_) | 7037 | 324121 |
| M13_MYALGIA | Myalgia | XIII Diseases of the musculoskeletal system and connective tissue (M13_) | 7024 | 275212 |
| CD2_BENIGN_COLON_NOS | Benign neoplasm: Colon, unspecified | II Neoplasms from hospital discharges (CD2_) | 7013 | 370264 |
| F5_BIPO | Bipolar affective disorders | V Mental and behavioural disorders (F5_) | 7006 | 329192 |
| N14_FEMGENPAIN | Pain and other conditions associated with female genital organs and menstrual cycle | XIV Diseases of the genitourinary system (N14_) | 6976 | 107564 |
| ST19_FRACT_SKULL_FACIAL_BONES | Fracture of skull and facial bones | XIX Injury, poisoning and certain other consequences of external causes (ST19_) | 6907 | 329318 |
| D3_BLOODOTHER | Other diseases of blood and blood-forming organs | III Diseases of the blood and blood-forming organs and certain disorders involving the immune mechanism (D3_) | 6877 | 370400 |
| N14_PHIMOSIS | Redundant prepuce, phimosis and paraphimosis | XIV Diseases of the genitourinary system (N14_) | 6859 | 119297 |
| K11_DISORD_TEETH | Other disorders of teeth and supporting structures | XI Diseases of the digestive system (K11_) | 6850 | 259234 |
| O15_PREG_MATERN_CARE | Maternal care for other conditions predominantly related to pregnancy | XV Pregnancy, childbirth and the puerperium (O15_) | 6845 | 179899 |
| FE | Focal epilepsy | VI Diseases of the nervous system (G6_) | 6842 | 365537 |
| H7_RETINOPATHYDIAB | Diabetic retinopathy | VII Diseases of the eye and adnexa (H7_) | 6818 | 344569 |
| F5_PERSMOOD | Persistent mood disorders | V Mental and behavioural disorders (F5_) | 6756 | 329192 |
| APPENDACUT_COMPLIC | Acute appendicitis, with complications | XI Diseases of the digestive system (K11_) | 6747 | 346283 |
| G6_MIGRAINE_NO_AURA | Migraine without aura | VI Diseases of the nervous system (G6_) | 6730 | 287837 |
| TRAUMBRAIN_NONCONCUS | severe traumatic brain injury, does not include concussion | Neurological endpoints | 6687 | 370590 |
| M13_CERVICALGIA | Cervicalgia | XIII Diseases of the musculoskeletal system and connective tissue (M13_) | 6684 | 270964 |
| L12_CELLULITIS | Cellulitis | XII Diseases of the skin and subcutaneous tissue (L12_) | 6683 | 355969 |
| O15_PREECLAMPS | Pre-eclampsia | XV Pregnancy, childbirth and the puerperium (O15_) | 6663 | 194266 |
| F5_SUBSNOALCO | Substance use, excluding alcohol | V Mental and behavioural disorders (F5_) | 6605 | 353355 |
| M13_OTHERBONE | Other disorders of bone | XIII Diseases of the musculoskeletal system and connective tissue (M13_) | 6568 | 358014 |
| H8_EXTOTITIS | Otitis externa | VIII Diseases of the ear and mastoid process (H8_) | 6547 | 367005 |
| H7_IRIDOCYC_ANTER | Anterior Iridocyclitis | VII Diseases of the eye and adnexa (H7_) | 6536 | 370741 |
| I9_INTRACRA | Nontraumatic intracranial haemmorrhage | IX Diseases of the circulatory system (I9_) | 6530 | 342673 |
| F5_SCHZPHR | Schizophrenia | V Mental and behavioural disorders (F5_) | 6515 | 364160 |
| C3_COLORECTAL_EXALLC | Colorectal cancer (controls excluding all cancers) | II Neoplasms, from cancer register (ICD-O-3) | 6509 | 287137 |
| AD_LO_EXMORE | Alzheimer's disease (Late onset) (more control exclusions) | Neurological endpoints | 6489 | 170429 |
| K11_PARODON_PERIAPIC_CHRONIC | Chronic periapical parodontitis | XI Diseases of the digestive system (K11_) | 6466 | 259234 |
| RX_INFERTILITY | Medical treatment for female infertility | Drug purchase endpoints | 6446 | 204424 |
| J10_RESPOTHER | Other diseases of the respiratory system | X Diseases of the respiratory system (J10_) | 6381 | 370896 |
| L12_URTICA_NAS | Other and unspecified urticaria | XII Diseases of the skin and subcutaneous tissue (L12_) | 6381 | 364583 |
| E4_PARATHYRO | Disorders of parathyroid gland | IV Endocrine, nutritional and metabolic diseases (E4_) | 6362 | 361988 |
| M13_PATELLAOTH | Other disorders of patella | XIII Diseases of the musculoskeletal system and connective tissue (M13_) | 6333 | 240862 |
| SPONDYLOARTHRITIS | Spondyloarthritis | Comorbidities of Interstitial lung disease endpoints | 6333 | 341455 |
| NIV | Non-invasive ventilation | Interstitial lung disease endpoints | 6315 | 164315 |
| ALCOHOLACUTE10 | Acute alcohol intoxication | Alcohol related diseases | 6289 | 369838 |

| | | | | |
|---|---|---|---|---|
| K11_SALIVARY | Diseases of salivary glands | XI Diseases of the digestive system (K11_) | 6269 | 259234 |
| K11_ANOMALI_DENTAL_ARCH_RELATIONS1_INCLAVO | Anomalies of dental arch relationship, including avohilmo | XI Diseases of the digestive system (K11_) | 6260 | 371017 |
| J10_NASALPOLYP | Nasal polyp | X Diseases of the respiratory system (J10_) | 6255 | 283342 |
| E4_DM1NASCOMP | Type 1 diabetes with other specified/multiple/unspecified complications | IV Endocrine, nutritional and metabolic diseases (E4_) | 6234 | 308280 |
| K11_ACUTPANC | Acute pancreatitis | XI Diseases of the digestive system (K11_) | 6223 | 330903 |
| FE_STRICT | Focal epilepsy, strict definition | VI Diseases of the nervous system (G6_) | 6213 | 365534 |
| ST19_FRACT_LUMBAR_SPINE_PELVIS | Fracture of lumbar spine and pelvis | XIX Injury, poisoning and certain other consequences of external causes (ST19_) | 6209 | 364504 |
| M13_ARTHROSIS_POLY | Polyarthrosis | XIII Diseases of the musculoskeletal system and connective tissue (M13_) | 6185 | 240862 |
| H7_IRIDOACUTE | Acute and subacute iridocyclitis | VII Diseases of the eye and adnexa (H7_) | 6166 | 357814 |
| F5_BEHEMOCHILD | Behavioural and emotional disorders with onset usually occurring in childhood and adolescence | V Mental and behavioural disorders (F5_) | 6160 | 371117 |
| RHEUMA_OTHER_WIDE | Other (seronegative) rheumatoid arthritis, wide | Rheuma endpoints | 6153 | 371124 |
| E4_GOITREMULTINOD | Nontoxic multinodular goitre | IV Endocrine, nutritional and metabolic diseases (E4_) | 6149 | 320703 |
| POLLENALLERG | Pollen allergy | Comorbidities of Asthma | 6118 | 368409 |
| N14_HYPERTROPHYBREAST_BOTH | Hypertrophy of breast in women and men | XIV Diseases of the genitourinary system (N14_) | 6105 | 359487 |
| DRY_AMD | Dry age-related macular degeneration (includes geographic atrophy) | VII Diseases of the eye and adnexa (H7_) | 6065 | 251042 |
| D3_IMMUNEMECHANISM | Certain disorders involving the immune mechanism | III Diseases of the blood and blood-forming organs and certain disorders involving the immune mechanism (D3_) | 6022 | 371255 |
| N14_FEMALE_GENITAL_HSIL | HSIL lesion of the cervix uteri, vagina or vulva | XIV Diseases of the genitourinary system (N14_) | 5996 | 204874 |
| C3_BREAST_ERNEG_EXALLC | Malignant neoplasm of breast, HER-negative (controls excluding all cancers) | II Neoplasms, from cancer register (ICD-O-3) | 5965 | 167017 |
| O15_MAT_CARE_MALPRESENT | Maternal care for known or suspected malpresentation of fetus | XV Pregnancy, childbirth and the puerperium (O15_) | 5957 | 168929 |
| K11_GULC | Gastric ulcer | XI Diseases of the digestive system (K11_) | 5935 | 320387 |
| K11_OTHILEUS | Other or unspecified ileus, impaction or obstruction | XI Diseases of the digestive system (K11_) | 5925 | 301931 |
| N14_ACUTERENFAIL | Acute renal failure | XIV Diseases of the genitourinary system (N14_) | 5908 | 363177 |
| I9_CARDMYO | Cardiomyopathy | IX Diseases of the circulatory system (I9_) | 5874 | 286109 |
| N14_ENDOMETRIOSIS_OVARY | Endometriosis of ovary | XIV Diseases of the genitourinary system (N14_) | 5867 | 107564 |
| ASTHMA_CHILD_EXMORE | Childhood asthma (age<16) (more control exclusions) | Asthma and related endpoints | 5865 | 202399 |
| AB1_VIRAL_NOS | Other viral diseases, not elsewhere classified | I Certain infectious and parasitic diseases (AB1_) | 5864 | 367472 |
| C3_BRONCHUS_LUNG_EXALLC | Malignant neoplasm of bronchus and lung (controls excluding all cancers) | II Neoplasms, from cancer register (ICD-O-3) | 5842 | 287137 |
| L12_SEBORRKERAT | Seborrhoeic keratosis | XII Diseases of the skin and subcutaneous tissue (L12_) | 5835 | 353088 |
| VWXY20_INTENTI_SELF_P_EXPOS_OTHER_UNSPE_CHEMIC_NOXIO_SUBST | Intentional self-poisoning by and exposure to other and unspecified chemicals and noxious substances | XX External causes of morbidity and mortality (VWXY20_) | 5811 | 371466 |
| K11_ORAL_LICHEN_PLANUS_WIDE | Oral lichen ruber planus, wide definition | XI Diseases of the digestive system (K11_) | 5791 | 371486 |
| ABDOM_HERNIA_POSTOP | Hernia of abdominal wall, postoperative | XI Diseases of the digestive system (K11_) | 5775 | 210314 |
| D3_COAGDEF_PURPUR_HAEMORRHAGIC | Coagulation defects, purpura and other haemorrhagic conditions | III Diseases of the blood and blood-forming organs and certain disorders involving the immune mechanism (D3_) | 5773 | 371504 |
| N14_ENDOMETRIOSIS_ASRM_STAGE1_2 | Endometriosis ASRM stages 1,2 | XIV Diseases of the genitourinary system (N14_) | 5769 | 205101 |
| N14_SALPHOOPH | Salpingitis and oophoritis | XIV Diseases of the genitourinary system (N14_) | 5768 | 188295 |
| M13_SPONDYLOLISTHESIS | Spondylolisthesis/Spondylolysis | XIII Diseases of the musculoskeletal system and connective tissue (M13_) | 5766 | 270964 |
| L12_PSORI_VULG | Psoriasis vulgaris | XII Diseases of the skin and subcutaneous tissue (L12_) | 5759 | 364071 |
| AB1_SEXUAL_TRANSMISSION_NOS | Other predominantly sexually transmitted diseases, not elsewhere classified | I Certain infectious and parasitic diseases (AB1_) | 5753 | 366340 |
| TEMPOROMANDIB | Temporomandibular joint disorders | XI Diseases of the digestive system (K11_) | 5668 | 205355 |
| O15_PREG_ECTOP | Ectopic pregnancy | XV Pregnancy, childbirth and the puerperium (O15_) | 5648 | 149622 |
| N14_GLOMERULAR | Glomerular diseases | XIV Diseases of the genitourinary system (N14_) | 5642 | 371635 |
| AB1_BACT_INTEST_OTH | Other bacterial intestinal infections | I Certain infectious and parasitic diseases (AB1_) | 5631 | 336396 |
| N14_ENDOMETRIOSIS_PELVICPERITONEUM | Endometriosis of pelvic peritoneum | XIV Diseases of the genitourinary system (N14_) | 5628 | 107564 |
| M13_HAMMERTOE | Other hammer toe(s) (acquired) | XIII Diseases of the musculoskeletal system and connective tissue (M13_) | 5625 | 240862 |
| CD2_BENIGN_RECTUM_ANAL | Benign neoplasm: Rectum, anus and anal canal | II Neoplasms from hospital discharges (CD2_) | 5613 | 371664 |
| E4_HYPERPARA | Hyperparathyroidism | IV Endocrine, nutritional and metabolic diseases (E4_) | 5590 | 361988 |
| G6_POLYOTHUNS | Other and unspecified polyneuropathies, also in other diseases | VI Diseases of the nervous system (G6_) | 5553 | 370790 |
| M13_RADICULOPATHY | Radiculopathy | XIII Diseases of the musculoskeletal system and connective tissue (M13_) | 5550 | 270964 |
| M13_ADHCAPSULITIS | Adhesive capsulitis of shoulder | XIII Diseases of the musculoskeletal system and connective tissue (M13_) | 5538 | 275212 |
| I9_AVBLOCK | AV-block | IX Diseases of the circulatory system (I9_) | 5536 | 286109 |
| K11_EROSION_INCLAVO | Erosion of teeth, including avohilmo | XI Diseases of the digestive system (K11_) | 5521 | 371756 |
| H7_CORNULCER | Corneal ulcer | VII Diseases of the eye and adnexa (H7_) | 5476 | 357814 |
| L12_FOLLICULARCYST | Follicular cysts of skin and subcutaneous tissue | XII Diseases of the skin and subcutaneous tissue (L12_) | 5459 | 361140 |
| N14_CERVICAL_HSIL | HSIL lesion of the cervix uteri | XIV Diseases of the genitourinary system (N14_) | 5459 | 205411 |
| K11_IMPACTED_TEETH_INCLAVO | Impacted teeth , including avohilmo | XI Diseases of the digestive system (K11_) | 5435 | 371842 |
| F5_ALZHDEMENT | Dementia in Alzheimer disease | V Mental and behavioural disorders (F5_) | 5422 | 356660 |
| C3_COLORECTAL_ADENO_EXALLC | Colorectal adenocarcinoma (controls excluding all cancers) | II Neoplasms, from cancer register (ICD-O-3) | 5378 | 287137 |
| I9_SAHANEUR | Aneurysms, operations, SAH | IX Diseases of the circulatory system (I9_) | 5342 | 342673 |
| CHILDHOOD_ALLERGY | Childhood allergy (age < 16) | Asthma and related endpoints | 5337 | 371940 |
| CD2_BENIGN_OVARY | Benign neoplasm of ovary | II Neoplasms from hospital discharges (CD2_) | 5308 | 205562 |
| G6_ROOTPLEXOTHUNS | Other and unspecified nerve root and plexus disorders, also in other diseases | VI Diseases of the nervous system (G6_) | 5294 | 330377 |
| GOUT_NOS | Gout, unspecified | Rheuma endpoints | 5292 | 368788 |
| KRA_PSY_SLEEP_NONORG_EXMORE | Nonorganic sleeping disorders (more control exclusions) | Psychiatric endpoints from Katri Räikkönen | 5254 | 277526 |
| N14_HYDROCELE | Hydrocele | XIV Diseases of the genitourinary system (N14_) | 5221 | 119297 |
| N14_HYPERTROPHYBREAST | Hypertrophy of breast | XIV Diseases of the genitourinary system (N14_) | 5212 | 193973 |
| E4_DM1OPTH | Type 1 diabetes with ophthalmic complications | IV Endocrine, nutritional and metabolic diseases (E4_) | 5202 | 308280 |
| F5_SOMATOFORM | Somatoform disorder | V Mental and behavioural disorders (F5_) | 5164 | 337577 |

| | | | | |
|---|---|---|---|---|
| E4_HYPOOSMNAT | Hypo-osmolality and hyponatraemia | IV Endocrine, nutritional and metabolic diseases (E4_) | 5137 | 324150 |
| INFLUENZA | All influenza (not pneumonia) | Asthma and related endpoints | 5136 | 314673 |
| M13_DUPUTRYEN | Palmar fascial fibromatosis [Dupuytren] | XIII Diseases of the musculoskeletal system and connective tissue (M13_) | 5128 | 275212[1] |
| N14_OTHFEMPELINF | Other female pelvic inflammatory diseases | XIV Diseases of the genitourinary system (N14_) | 5117 | 188295 |
| VOCALCORDDYS | Vocal cord dysfunction | COPD and related endpoints | 5087 | 139795 |
| CD2_BENIGN_OTHERDIGESTIVE | Benign neoplasm of other and ill-defined parts of digestive system | II Neoplasms from hospital discharges (CD2_) | 5062 | 372215 |
| K11_DISLIVOTH | Other diseases of liver | XI Diseases of the digestive system (K11_) | 5052 | 366450 |
| K11_UC_STRICT2 | Ulcerative colitis (strict definition, require KELA, min 2 HDR) | XI Diseases of the digestive system (K11_) | 5034 | 371530 |
| L12_LICHENPLANUS_INCLAVO | Lichen planus, including avohilmo | XII Diseases of the skin and subcutaneous tissue (L12_) | 5015 | 372262 |

c) **4 DEs**: We downloaded and manually harmonized (e.g., GRCh 37, flip the effect allele, etc.) the 4 DE GWAS summary statistics from the PGC website (https://pgc.unc.edu/for-researchers/download-results/).

| Trait | Data set | URL | PubMed ID | Ancestry | Num of Cases | Num of Controls |
|---|---|---|---|---|---|---|
| AD | PGC | https://ctg.cncr.nl/software/summary_statistics | 34493870 | European | 90338 | 1036225 |
| ADHD | PGC | https://figshare.com/articles/dataset/adhd2022/22564390 | 36702997 | European | 38691 | 1868432 |
| BIP | PGC | https://figshare.com/articles/dataset/PGC3_bipolar_disorder_GWAS_summary_statistics/14102594 | 34002096 | European | 41917 | 371549 |
| SCZ | PGC | https://figshare.com/articles/dataset/scz2022/19426775 | 35396580 | European | 76755 | 243649 |

**eTable 2: The significant genetic correlation results for both the LDSC and GNOVA methods.**

To present results in **Fig. 2**, we ensured that common significant results survived the Bonferroni correction in both the LDSC and GNOVA methods. A large proportion of tests were not converged using the HDL method.

| MAE | DE | gc_mean | gc_ldsc_se | P_ldsc | gc_gnova | P_gnova |
|---|---|---|---|---|---|---|
| Brain_age_gap | RX_ANTIHYP | 0.1444 | 0.0326 | 9.32E-06 | 0.106669 | 7.2E-07 |
| Cardiovascular_age_gap | RX_ANTIHYP | 0.4188 | 0.0308 | 4.54E-42 | 0.372279 | 2.4E-44 |
| Pulmonary_age_gap | RX_ANTIHYP | 0.1335 | 0.0271 | 8.07E-07 | 0.125543 | 2.86E-10 |
| fmri | RX_ANTIHYP | 0.2778 | 0.0654 | 2.18E-05 | 0.220129 | 3.78E-09 |
| Cardiovascular_age_gap | FG_CVD | 0.2782 | 0.0338 | 1.98E-16 | 0.263942 | 4.33E-27 |
| Pulmonary_age_gap | FG_CVD | 0.1578 | 0.0313 | 4.8E-07 | 0.116153 | 3.69E-08 |
| Cardiovascular_age_gap | RX_STATIN | 0.1801 | 0.0415 | 1.42E-05 | 0.162826 | 2.45E-15 |
| Metabolic_age_gap | RX_STATIN | 0.3658 | 0.0806 | 5.62E-06 | 0.364051 | 7.71E-21 |
| Pulmonary_age_gap | PULM_INFECTIONS | 0.1786 | 0.0377 | 2.17E-06 | 0.185103 | 3.61E-13 |
| Brain_age_gap | I9_HYPTENS | 0.143 | 0.0321 | 8.2E-06 | 0.121863 | 4.23E-08 |
| Cardiovascular_age_gap | I9_HYPTENS | 0.4429 | 0.0319 | 6E-44 | 0.388782 | 1.23E-39 |
| Pulmonary_age_gap | I9_HYPTENS | 0.1723 | 0.0299 | 8.24E-09 | 0.147139 | 1.5E-09 |
| fmri | I9_HYPTENS | 0.2818 | 0.0657 | 1.78E-05 | 0.220309 | 8.31E-09 |
| Musculoskeletal_age_gap | KRA_PSY_ANYMENTAL | 0.1588 | 0.0362 | 1.18E-05 | 0.129086 | 2.8E-08 |
| Renal_age_gap | M13_ARTHROSIS_INCLAVO | 0.1435 | 0.0327 | 1.14E-05 | 0.119596 | 1.26E-06 |
| Cardiovascular_age_gap | I9_HYPTENSESS | 0.4403 | 0.0322 | 1.73E-42 | 0.389667 | 2.43E-39 |
| Pulmonary_age_gap | I9_HYPTENSESS | 0.1722 | 0.0304 | 1.55E-08 | 0.150207 | 1.8E-09 |
| Cardiovascular_age_gap | FG_OTHHEART | 0.2447 | 0.0364 | 1.73E-11 | 0.22568 | 2.96E-22 |
| Pulmonary_age_gap | RX_CROHN_1STLINE | 0.3045 | 0.0331 | 3.58E-20 | 0.252513 | 7.41E-17 |
| Cardiovascular_age_gap | CARDIAC_ARRHYTM | 0.1665 | 0.0377 | 9.97E-06 | 0.126846 | 3.83E-07 |
| Pulmonary_age_gap | J10_LOWCHRON | 0.5025 | 0.0295 | 2.98E-65 | 0.429621 | 1.11E-28 |
| Cardiovascular_age_gap | I9_IHD | 0.2631 | 0.0375 | 2.35E-12 | 0.225973 | 6.29E-19 |
| Pulmonary_age_gap | I9_IHD | 0.1585 | 0.0341 | 3.43E-06 | 0.12014 | 2.42E-08 |
| Musculoskeletal_age_gap | O15_PREG_ABORT | 0.2258 | 0.0528 | 1.9E-05 | 0.163454 | 8.7E-09 |
| Cardiovascular_age_gap | I9_CVD_HARD | 0.1926 | 0.0396 | 1.17E-06 | 0.189534 | 3.79E-13 |
| Cardiovascular_age_gap | T2D | 0.1655 | 0.0311 | 1.07E-07 | 0.153936 | 8.39E-13 |
| Metabolic_age_gap | T2D | 0.3991 | 0.0478 | 7.11E-17 | 0.296483 | 2.76E-34 |
| Pulmonary_age_gap | T2D | 0.1443 | 0.0282 | 3.2E-07 | 0.117147 | 4.83E-10 |
| Pulmonary_age_gap | BRONCHITIS | 0.2434 | 0.0531 | 4.65E-06 | 0.183546 | 4.71E-11 |
| Cardiovascular_age_gap | KELA_DIAB_INSUL_EXMORE | 0.1681 | 0.0316 | 1.04E-07 | 0.161105 | 6.61E-10 |
| Metabolic_age_gap | KELA_DIAB_INSUL_EXMORE | 0.3506 | 0.0492 | 1.02E-12 | 0.267648 | 1.13E-29 |
| Pulmonary_age_gap | KELA_DIAB_INSUL_EXMORE | 0.1785 | 0.0327 | 4.67E-08 | 0.142112 | 7.37E-06 |
| Cardiovascular_age_gap | E4_METABOLIA | 0.2464 | 0.0543 | 5.68E-06 | 0.196128 | 2.6E-10 |
| C256_225 | F5_DEPRESSION_DYSTHYMIA | -0.2291 | 0.0536 | 1.92E-05 | -0.18465 | 4.57E-06 |
| Cardiovascular_age_gap | I9_CORATHER | 0.221 | 0.0361 | 8.83E-10 | 0.193513 | 5.5E-16 |
| Cardiovascular_age_gap | E4_DM2NASCOMP | 0.1562 | 0.0313 | 5.92E-07 | 0.154115 | 8.98E-12 |
| Metabolic_age_gap | E4_DM2NASCOMP | 0.3854 | 0.0473 | 3.67E-16 | 0.289669 | 7.06E-34 |
| Pulmonary_age_gap | E4_DM2NASCOMP | 0.1616 | 0.0293 | 3.6E-08 | 0.125003 | 1.11E-09 |
| Cardiovascular_age_gap | I9_AF | 0.2226 | 0.0368 | 1.39E-09 | 0.183633 | 3.28E-18 |
| Renal_age_gap | M13_ARTHROSIS_KNEE | 0.1579 | 0.0339 | 3.1E-06 | 0.127658 | 2.14E-07 |
| Cardiovascular_age_gap | I9_CHD | 0.2004 | 0.037 | 6.27E-08 | 0.180157 | 1.75E-13 |
| C512_453 | M13_SOFTTISSUEOTH | -0.2099 | 0.0493 | 2.04E-05 | -0.14428 | 1.88E-06 |
| C1024_312 | M13_SOFTTISSUEOTH | 0.3249 | 0.0726 | 7.72E-06 | 0.162773 | 1.81E-05 |
| Pulmonary_age_gap | J10_ASTHMA_EXMORE | 0.446 | 0.0318 | 9.4E-45 | 0.382263 | 1.08E-24 |
| C1024_635 | G6_SLEEPAPNO_INCLAVO | -0.2023 | 0.0416 | 1.17E-06 | -0.11426 | 7.25E-06 |
| C1024_764 | G6_SLEEPAPNO_INCLAVO | -0.1958 | 0.0431 | 5.48E-06 | -0.1163 | 2.68E-06 |
| Pulmonary_age_gap | J10_ASTHMACOPDKELA | 0.5099 | 0.0308 | 9.84E-62 | 0.430036 | 7.06E-28 |
| Pulmonary_age_gap | AB1_INTESTINAL_INFECTIONS | 0.2283 | 0.0482 | 2.18E-06 | 0.1755 | 5.01E-07 |
| C512_304 | KRA_PSY_ANXIETY_EXMORE | 0.1768 | 0.0382 | 3.74E-06 | 0.103642 | 1.4E-05 |
| C1024_304 | KRA_PSY_ANXIETY_EXMORE | 0.1839 | 0.0391 | 2.57E-06 | 0.108724 | 8.28E-06 |
| C1024_556 | KRA_PSY_ANXIETY_EXMORE | 0.1852 | 0.0381 | 1.17E-06 | 0.108821 | 2.37E-05 |
| Musculoskeletal_age_gap | KRA_PSY_ANXIETY_EXMORE | 0.1337 | 0.0314 | 2.08E-05 | 0.118202 | 1.48E-08 |
| Cardiovascular_age_gap | C_STROKE | 0.2158 | 0.0498 | 1.48E-05 | 0.196617 | 1.2E-08 |
| C1024_635 | G6_SLEEPAPNO | -0.1978 | 0.0424 | 3.01E-06 | -0.11072 | 1.42E-05 |
| C1024_764 | G6_SLEEPAPNO | -0.1873 | 0.0436 | 1.7E-05 | -0.10791 | 1.58E-05 |
| Cardiovascular_age_gap | T2D_WIDE | 0.1686 | 0.0346 | 1.06E-06 | 0.156349 | 2.55E-11 |
| Metabolic_age_gap | T2D_WIDE | 0.3755 | 0.0467 | 9.21E-16 | 0.27996 | 1.1E-28 |
| Pulmonary_age_gap | T2D_WIDE | 0.1545 | 0.032 | 1.33E-06 | 0.128957 | 1.6E-09 |
| C256_123 | M13_SPONDYLOPATHY | 0.1992 | 0.0413 | 1.38E-06 | 0.115212 | 1.44E-05 |
| C512_123 | M13_SPONDYLOPATHY | 0.1973 | 0.0414 | 1.86E-06 | 0.115928 | 1.76E-05 |
| C1024_123 | M13_SPONDYLOPATHY | 0.1963 | 0.0416 | 2.34E-06 | 0.117235 | 1.68E-05 |
| Musculoskeletal_age_gap | O15_ABORT_MEDICAL | 0.2216 | 0.0428 | 2.27E-07 | 0.155748 | 8.65E-10 |
| Pulmonary_age_gap | O15_ABORT_MEDICAL | 0.1346 | 0.0319 | 2.37E-05 | 0.129189 | 3.11E-09 |
| Cardiovascular_age_gap | I9_ANGINA | 0.2532 | 0.0366 | 4.77E-12 | 0.22528 | 2.09E-19 |
| Pulmonary_age_gap | J10_ASTHMA_MAIN_EXMORE | 0.4359 | 0.0333 | 3.82E-39 | 0.370781 | 8.17E-23 |
| Pulmonary_age_gap | AB1_GASTROENTERITIS_NOS | 0.2299 | 0.0531 | 1.47E-05 | 0.177975 | 6.05E-06 |
| C512_312 | JOINTPAIN | 0.2784 | 0.0646 | 1.63E-05 | 0.158634 | 8.13E-06 |
| C1024_312 | JOINTPAIN | 0.3206 | 0.0646 | 6.95E-07 | 0.157464 | 1.73E-06 |
| Cardiovascular_age_gap | D3_ANAEMIA | 0.2253 | 0.0461 | 1.03E-06 | 0.252342 | 2.59E-12 |
| Cardiovascular_age_gap | I9_HEARTFAIL | 0.2657 | 0.0483 | 3.87E-08 | 0.201462 | 1.79E-12 |
| Cardiovascular_age_gap | I9_HEARTFAIL_EXMORE | 0.2729 | 0.0446 | 9.59E-10 | 0.214822 | 3.55E-14 |
| Pulmonary_age_gap | I9_HEARTFAIL_EXMORE | 0.1717 | 0.04 | 1.77E-05 | 0.139845 | 4.38E-07 |
| C1024_556 | G6_OTHNEU | 0.3281 | 0.0725 | 6.02E-06 | 0.194371 | 1.94E-06 |
| Cardiovascular_age_gap | I9_HEARTFAIL_ALLCAUSE | 0.2555 | 0.0484 | 1.32E-07 | 0.195834 | 9.09E-12 |
| Musculoskeletal_age_gap | K11_REFLUX | 0.2132 | 0.0469 | 5.46E-06 | 0.141282 | 4.86E-07 |
| Cardiovascular_age_gap | I9_HEARTFAIL_AND_ANTIHYPERT | 0.3984 | 0.0372 | 8.68E-27 | 0.355048 | 6.75E-38 |

| | | | | | | |
|---|---|---|---|---|---|---|
| Pulmonary_age_gap | I9_HEARTFAIL_AND_ANTIHYPERT | 0.1664 | 0.0324 | 2.76E-07 | 0.143174 | 3.26E-10 |
| Cardiovascular_age_gap | I9_MI_STRICT | 0.1989 | 0.042 | 2.21E-06 | 0.180497 | 4.04E-11 |
| Musculoskeletal_age_gap | KRA_PSY_SUBSTANCE_EXMORE | 0.1711 | 0.0402 | 2.05E-05 | 0.133814 | 1.26E-08 |
| Pulmonary_age_gap | KRA_PSY_SUBSTANCE_EXMORE | 0.1952 | 0.0345 | 1.54E-08 | 0.169022 | 5.42E-10 |
| Cardiovascular_age_gap | I9_REVASC | 0.1672 | 0.0348 | 1.53E-06 | 0.15883 | 1.84E-12 |
| Cardiovascular_age_gap | FG_DOAAC | 0.2052 | 0.0421 | 1.09E-06 | 0.207028 | 3.81E-13 |
| Pulmonary_age_gap | AUD_SWEDISH | 0.148 | 0.0346 | 1.87E-05 | 0.137955 | 3.42E-08 |
| Pulmonary_age_gap | ASTHMA_INFECTIONS | 0.4334 | 0.0354 | 1.93E-34 | 0.387842 | 1.08E-26 |
| Cardiovascular_age_gap | I9_AF_REIMB | 0.2063 | 0.0367 | 1.91E-08 | 0.176208 | 4.7E-16 |
| Pulmonary_age_gap | ASTHMA_NAS | 0.4255 | 0.0406 | 1.06E-25 | 0.378746 | 2.47E-32 |
| C1024_808 | E4_OBESITY | 0.1749 | 0.0388 | 6.69E-06 | 0.136357 | 2.08E-07 |
| Pulmonary_age_gap | E4_OBESITY | 0.1166 | 0.0272 | 1.84E-05 | 0.120343 | 5.05E-10 |
| Renal_age_gap | E4_OBESITY | 0.158 | 0.0276 | 9.86E-09 | 0.173367 | 4.94E-16 |
| Pulmonary_age_gap | J10_LOWERINF | 0.4304 | 0.0858 | 5.19E-07 | 0.319966 | 1.4E-11 |
| C256_123 | M13_SPONDYLOSIS | 0.2377 | 0.0444 | 8.32E-08 | 0.134184 | 5.04E-06 |
| C512_123 | M13_SPONDYLOSIS | 0.2383 | 0.0448 | 1.04E-07 | 0.135421 | 5.97E-06 |
| C1024_123 | M13_SPONDYLOSIS | 0.2355 | 0.0447 | 1.4E-07 | 0.13631 | 5.87E-06 |
| Renal_age_gap | M13_ARTHROSIS_KNEE_PRIM_KNEESURG | 0.1602 | 0.0365 | 1.16E-05 | 0.132186 | 1.4E-08 |
| Pulmonary_age_gap | J10_COPD | 0.4517 | 0.0345 | 4.31E-39 | 0.40364 | 5.23E-32 |
| C1024_241 | CD2_BENIGN_COLON | 0.3576 | 0.0796 | 7.01E-06 | 0.187234 | 5.16E-06 |
| Metabolic_age_gap | DM_SEVERAL_COMPLICATIONS | 0.305 | 0.0596 | 3.11E-07 | 0.235762 | 4.1E-19 |
| Pulmonary_age_gap | ASTHMA_ACUTE_RESPIRATORY_INFECTIONS | 0.3792 | 0.039 | 2.27E-22 | 0.352279 | 8.58E-22 |
| Pulmonary_age_gap | J10_BRONCHITIS | 0.4361 | 0.0968 | 6.57E-06 | 0.316815 | 6.14E-14 |
| Pulmonary_age_gap | E4_OBESITYCAL | 0.1395 | 0.0302 | 3.75E-06 | 0.129223 | 2.37E-11 |
| Renal_age_gap | E4_OBESITYCAL | 0.1919 | 0.0319 | 1.84E-09 | 0.18948 | 1.26E-15 |
| C1024_713 | I9_ATHSCLE | 0.2317 | 0.0521 | 8.84E-06 | 0.179845 | 2.02E-07 |
| Cardiovascular_age_gap | I9_ATHSCLE | 0.2435 | 0.0502 | 1.24E-06 | 0.233697 | 1.95E-11 |
| Cardiovascular_age_gap | O15_HYPTENSPREG | 0.322 | 0.0472 | 9.3E-12 | 0.283797 | 4.42E-16 |
| Musculoskeletal_age_gap | O15_FALSE_LAB | 0.3161 | 0.0662 | 1.79E-06 | 0.188508 | 1.15E-08 |
| C512_123 | M13_DORSALGIANAS | 0.2932 | 0.0585 | 5.47E-07 | 0.160357 | 1.94E-05 |
| C1024_123 | M13_DORSALGIANAS | 0.2889 | 0.0586 | 8.21E-07 | 0.159974 | 2.27E-05 |
| Cardiovascular_age_gap | I9_UAP | 0.2161 | 0.0417 | 2.13E-07 | 0.20669 | 2.41E-16 |
| Cardiovascular_age_gap | I9_HEARTFAIL_AND_CHD | 0.264 | 0.0438 | 1.61E-09 | 0.193557 | 4.02E-15 |
| Pulmonary_age_gap | ASTHMA_PNEUMONIA | 0.4945 | 0.0441 | 3.51E-29 | 0.413686 | 3.22E-27 |
| Pulmonary_age_gap | COPD_LATER | 0.481 | 0.0556 | 5.28E-18 | 0.380091 | 4.14E-19 |
| Pulmonary_age_gap | ASTHMA_OBESITY | 0.3285 | 0.0418 | 3.73E-15 | 0.303436 | 1.08E-21 |
| Pulmonary_age_gap | ALLERG_ASTHMA | 0.407 | 0.0412 | 4.58E-23 | 0.325213 | 8.35E-15 |
| Cardiovascular_age_gap | O15_GESTAT_HYPERT | 0.3418 | 0.0511 | 2.23E-11 | 0.301109 | 8.44E-15 |
| Cardiovascular_age_gap | I9_HYPTENSHD | 0.4689 | 0.0495 | 2.52E-21 | 0.381895 | 2.15E-27 |
| Cardiovascular_age_gap | I9_AORTANEUR | -0.2826 | 0.0618 | 4.85E-06 | -0.21045 | 2.11E-09 |
| Cardiovascular_age_gap | O15_PRE_OR_ECLAMPSIA | 0.2644 | 0.0598 | 9.91E-06 | 0.241041 | 3.51E-10 |
| Pulmonary_age_gap | NONALLERG_ASTHMA_EXMORE | 0.5206 | 0.0705 | 1.53E-13 | 0.382707 | 5.73E-21 |
| Pulmonary_age_gap | COPD_EARLY | 0.4711 | 0.039 | 1.56E-33 | 0.417978 | 7.94E-36 |
| Cardiovascular_age_gap | O15_PREECLAMPS | 0.2847 | 0.0602 | 2.25E-06 | 0.258085 | 1.48E-10 |
| Pulmonary_age_gap | F5_SUBSNOALCO | 0.2322 | 0.0514 | 6.13E-06 | 0.204602 | 5.19E-11 |
| Pulmonary_age_gap | ASTHMA_CHILD_EXMORE | 0.3362 | 0.0558 | 1.64E-09 | 0.257418 | 4.78E-09 |
| C1024_1005 | TEMPOROMANDIB | 0.4156 | 0.0963 | 1.58E-05 | 0.220543 | 7.42E-06 |
| C256_123 | M13_RADICULOPATHY | 0.3281 | 0.0777 | 2.44E-05 | 0.189334 | 8.38E-06 |
| Pulmonary_age_gap | CHILDHOOD_ALLERGY | 0.3364 | 0.0775 | 1.43E-05 | 0.177896 | 3.09E-07 |
| C512_239 | BIP | -0.1329 | 0.0311 | 1.9E-05 | -0.10786 | 3.69E-06 |
| C512_368 | BIP | -0.1619 | 0.0352 | 4.24E-06 | -0.13001 | 4.51E-06 |
| C1024_114 | BIP | -0.1488 | 0.0313 | 1.95E-06 | -0.11544 | 1.73E-06 |
| C1024_191 | BIP | -0.1416 | 0.033 | 1.81E-05 | -0.10807 | 5.01E-06 |
| C1024_239 | BIP | -0.1308 | 0.0295 | 9.13E-06 | -0.11066 | 2.62E-06 |
| C1024_844 | BIP | -0.1396 | 0.0315 | 9.38E-06 | -0.11634 | 5.3E-07 |
| C1024_1012 | BIP | -0.1347 | 0.0313 | 1.73E-05 | -0.10447 | 1.7E-05 |

**eTable 3: The significant MR results from the 2024 MAEs to the 525 DEs**

After two layers of Bonferroni correction, we obtained 39 significant results for the causal relationship from the 2024 MAEs to the 525 DEs.

| outcome | exposure | method | N_snp | b | b_se | P-value | or | or_lci95 | or_uci95 |
|---|---|---|---|---|---|---|---|---|---|
| MIGRAINE_TRIPTAN | C32_4 | Inverse variance weighted | 41 | 0.121628 | 0.028871 | 2.52E-05 | 1.129334 | 1.067204 | 1.195082 |
| MIGRAINE_TRIPTAN | C64_4 | Inverse variance weighted | 42 | 0.121874 | 0.028143 | 1.49E-05 | 1.129612 | 1.06899 | 1.193671 |
| N14_URETHRAOTH | C1024_342 | Inverse variance weighted | 8 | 0.284623 | 0.063331 | 6.98E-06 | 1.329261 | 1.17409 | 1.504939 |
| KRA_PSY_ANYMENTAL | C1024_430 | Inverse variance weighted | 12 | -0.12949 | 0.028259 | 4.6E-06 | 0.878545 | 0.831207 | 0.928578 |
| KELA_DIAB_INSUL_EXMORE | C1024_448 | Inverse variance weighted | 8 | -0.23423 | 0.052857 | 9.37E-06 | 0.791183 | 0.713319 | 0.877546 |
| K11_VENTHER | C1024_573 | Inverse variance weighted | 10 | 0.46266 | 0.114794 | 5.57E-05 | 1.588293 | 1.268282 | 1.989048 |
| K11_VENTHER | C1024_684 | Inverse variance weighted | 18 | 0.360381 | 0.067875 | 1.1E-07 | 1.433876 | 1.255265 | 1.637901 |
| I9_DISVEINLYMPH | C1024_726 | Inverse variance weighted | 9 | -0.18175 | 0.041443 | 1.16E-05 | 0.833806 | 0.768754 | 0.904362 |
| RX_ANTIHYP | Cardiovascular_age_gap | Inverse variance weighted | 37 | 0.5164 | 0.107829 | 1.68E-06 | 1.675983 | 1.3567 | 2.070405 |
| RX_STATIN | Cardiovascular_age_gap | Inverse variance weighted | 37 | 0.296236 | 0.057916 | 3.14E-07 | 1.344787 | 1.200479 | 1.506443 |
| I9_IHD | Cardiovascular_age_gap | Inverse variance weighted | 37 | 0.495007 | 0.093224 | 1.1E-07 | 1.64051 | 1.366549 | 1.969393 |
| I9_CVD_HARD | Cardiovascular_age_gap | Inverse variance weighted | 37 | 0.388983 | 0.085245 | 5.04E-06 | 1.47548 | 1.248452 | 1.743794 |
| E4_METABOLIA | Cardiovascular_age_gap | Inverse variance weighted | 37 | 0.231611 | 0.057193 | 5.13E-05 | 1.260629 | 1.126947 | 1.410169 |
| I9_CORATHER | Cardiovascular_age_gap | Inverse variance weighted | 37 | 0.55913 | 0.112018 | 5.99E-07 | 1.749149 | 1.40435 | 2.178604 |
| I9_CHD | Cardiovascular_age_gap | Inverse variance weighted | 37 | 0.498727 | 0.110528 | 6.41E-06 | 1.646624 | 1.325902 | 2.044925 |
| I9_ANGINA | Cardiovascular_age_gap | Inverse variance weighted | 37 | 0.586462 | 0.120701 | 1.18E-06 | 1.797618 | 1.418908 | 2.277406 |
| I9_HEARTFAIL_AND_ANTIHYPERT | Cardiovascular_age_gap | Inverse variance weighted | 37 | 0.677346 | 0.132275 | 3.04E-07 | 1.968647 | 1.519054 | 2.551305 |
| I9_MI_STRICT | Cardiovascular_age_gap | Inverse variance weighted | 37 | 0.536533 | 0.12364 | 1.43E-05 | 1.710068 | 1.342051 | 2.179002 |
| I9_REVASC | Cardiovascular_age_gap | Inverse variance weighted | 37 | 0.759398 | 0.16355 | 3.43E-06 | 2.13699 | 1.550908 | 2.94455 |
| I9_ANGIO | Cardiovascular_age_gap | Inverse variance weighted | 37 | 0.691944 | 0.149555 | 3.72E-06 | 1.997596 | 1.490059 | 2.678008 |
| I9_UAP | Cardiovascular_age_gap | Inverse variance weighted | 37 | 0.607329 | 0.137226 | 9.61E-06 | 1.835522 | 1.402655 | 2.401974 |
| I9_CABG | Cardiovascular_age_gap | Inverse variance weighted | 37 | 0.872591 | 0.206066 | 2.29E-05 | 2.393104 | 1.597918 | 3.584007 |
| RX_STATIN | Metabolic_age_gap | Inverse variance weighted | 67 | 0.630057 | 0.133458 | 2.35E-06 | 1.877718 | 1.445534 | 2.439116 |
| E4_LIPOPROT | Metabolic_age_gap | Inverse variance weighted | 67 | 0.418715 | 0.103112 | 4.89E-05 | 1.520008 | 1.241867 | 1.860443 |
| E4_HYPERCHOL | Metabolic_age_gap | Inverse variance weighted | 67 | 0.460656 | 0.110526 | 3.08E-05 | 1.585113 | 1.276375 | 1.96853 |
| RX_CROHN_1STLINE | Pulmonary_age_gap | Inverse variance weighted | 59 | 0.162068 | 0.033818 | 1.65E-06 | 1.17594 | 1.100522 | 1.256526 |
| J10_LOWCHRON | Pulmonary_age_gap | Inverse variance weighted | 59 | 0.47783 | 0.044435 | 5.7E-27 | 1.612572 | 1.478071 | 1.759311 |
| J10_ASTHMA_EXMORE | Pulmonary_age_gap | Inverse variance weighted | 59 | 0.515135 | 0.054713 | 4.72E-21 | 1.673864 | 1.503654 | 1.863342 |
| J10_ASTHMACOPDKELA | Pulmonary_age_gap | Inverse variance weighted | 59 | 0.62383 | 0.061108 | 1.81E-24 | 1.866061 | 1.655427 | 2.103495 |
| J10_ASTHMA_MAIN_EXMORE | Pulmonary_age_gap | Inverse variance weighted | 59 | 0.539305 | 0.06128 | 1.36E-18 | 1.714814 | 1.520739 | 1.933656 |
| ASTHMA_INFECTIONS | Pulmonary_age_gap | Inverse variance weighted | 59 | 0.524477 | 0.057305 | 5.57E-20 | 1.689575 | 1.510075 | 1.890413 |
| ASTHMA_NAS | Pulmonary_age_gap | Inverse variance weighted | 59 | 0.526937 | 0.06143 | 9.67E-18 | 1.693736 | 1.501603 | 1.910453 |
| J10_COPD | Pulmonary_age_gap | Inverse variance weighted | 59 | 0.572531 | 0.062029 | 2.71E-20 | 1.772749 | 1.569808 | 2.001924 |
| ASTHMA_ACUTE_RESPIRATORY_INFECTIONS | Pulmonary_age_gap | Inverse variance weighted | 59 | 0.538785 | 0.069642 | 1.02E-14 | 1.713923 | 1.495241 | 1.964588 |
| ASTHMA_PNEUMONIA | Pulmonary_age_gap | Inverse variance weighted | 59 | 0.516694 | 0.067221 | 1.51E-14 | 1.676476 | 1.469526 | 1.91257 |
| COPD_LATER | Pulmonary_age_gap | Inverse variance weighted | 59 | 0.433138 | 0.07697 | 1.83E-08 | 1.542089 | 1.326146 | 1.793196 |
| ALLERG_ASTHMA | Pulmonary_age_gap | Inverse variance weighted | 59 | 0.486942 | 0.086125 | 1.57E-08 | 1.627332 | 1.374563 | 1.926582 |
| COPD_EARLY | Pulmonary_age_gap | Inverse variance weighted | 59 | 0.705393 | 0.096803 | 3.17E-13 | 2.024642 | 1.674743 | 2.447645 |
| SCZ | C1024_598 | Inverse variance weighted | 7 | -0.37 | 0.07 | 9.89E-8 | 0.68770 | 0.59922 | 0.78924 |

278

**eTable 4: The significant MR results from the 525 Des to the 2024 MAEs**

After two layers of Bonferroni correction, we obtained 47 significant results for the causal relationship from the 525 DEs to the 2024 MAEs.

| outcome | exposure | method | N_snp | b | b_se | P-value | or | or_lci95 | or_uci95 |
|---|---|---|---|---|---|---|---|---|---|
| C512_6 | M13_SPONDYLOPATHY | Inverse variance weighted | 18 | 0.142042 | 0.034195 | 3.27E-05 | 1.152625 | 1.077906 | 1.232523 |
| C1024_43 | F5_DEMENTIA_INCLAVO | Inverse variance weighted | 8 | 0.075009 | 0.017305 | 1.46E-05 | 1.077894 | 1.041948 | 1.115081 |
| C128_13 | F5_DEMENTIA | Inverse variance weighted | 8 | 0.071625 | 0.01722 | 3.19E-05 | 1.074252 | 1.038601 | 1.111127 |
| C512_489 | F5_DEMENTIA | Inverse variance weighted | 8 | 0.076354 | 0.017746 | 1.69E-05 | 1.079345 | 1.042448 | 1.117549 |
| C1024_43 | F5_DEMENTIA | Inverse variance weighted | 8 | 0.077896 | 0.017868 | 0.000013 | 1.08101 | 1.043808 | 1.119538 |
| C1024_847 | F5_DEMENTIA | Inverse variance weighted | 8 | -0.07945 | 0.017734 | 7.47E-06 | 0.923625 | 0.892072 | 0.956294 |
| C1024_881 | F5_DEMENTIA | Inverse variance weighted | 8 | -0.08244 | 0.02008 | 4.04E-05 | 0.920868 | 0.885329 | 0.957834 |
| C512_335 | G6_AD_WIDE | Inverse variance weighted | 10 | -0.06586 | 0.014882 | 9.61E-06 | 0.936258 | 0.909344 | 0.963969 |
| C1024_335 | G6_AD_WIDE | Inverse variance weighted | 10 | -0.06828 | 0.014882 | 4.47E-06 | 0.933999 | 0.90715 | 0.961643 |
| C1024_714 | RHEUMA_SEROPOS_OTH | Inverse variance weighted | 24 | -0.04246 | 0.010166 | 2.96E-05 | 0.95843 | 0.939521 | 0.977719 |
| C512_342 | M13_GOUT | Inverse variance weighted | 7 | -0.07598 | 0.015492 | 9.38E-07 | 0.926837 | 0.899118 | 0.955412 |
| C512_296 | E4_DM1NASCOMP | Inverse variance weighted | 20 | -0.03075 | 0.006313 | 1.11E-06 | 0.969719 | 0.957794 | 0.981792 |
| C512_316 | E4_DM1NASCOMP | Inverse variance weighted | 20 | -0.04029 | 0.007216 | 2.36E-08 | 0.960514 | 0.947025 | 0.974195 |
| C1024_316 | E4_DM1NASCOMP | Inverse variance weighted | 20 | -0.03974 | 0.006949 | 1.07E-08 | 0.961036 | 0.948035 | 0.974215 |
| C1024_777 | E4_DM1NASCOMP | Inverse variance weighted | 20 | -0.03707 | 0.006776 | 4.49E-08 | 0.963611 | 0.950898 | 0.976494 |
| C512_142 | D3_IMMUNEMECHANISM | Inverse variance weighted | 10 | -0.08011 | 0.017363 | 3.96E-06 | 0.923019 | 0.892136 | 0.954971 |
| Brain_age_gap | G6_AD_WIDE | Inverse variance weighted | 8 | 0.096317 | 0.016834 | 1.05E-08 | 1.101108 | 1.065371 | 1.138044 |
| Cardiovascular_age_gap | RX_ANTIHYP | Inverse variance weighted | 87 | 0.23742 | 0.023887 | 2.81E-23 | 1.267974 | 1.209978 | 1.32875 |
| Cardiovascular_age_gap | FG_CVD | Inverse variance weighted | 34 | 0.249638 | 0.053537 | 3.12E-06 | 1.283561 | 1.155699 | 1.425569 |
| Cardiovascular_age_gap | I9_HYPTENS | Inverse variance weighted | 110 | 0.210271 | 0.018144 | 4.67E-31 | 1.234013 | 1.190901 | 1.278686 |
| Cardiovascular_age_gap | I9_HYPTENSESS | Inverse variance weighted | 78 | 0.204266 | 0.023308 | 1.89E-18 | 1.226624 | 1.171849 | 1.283959 |
| Cardiovascular_age_gap | O15_HYPTENSPREG | Inverse variance weighted | 7 | 0.212637 | 0.030338 | 2.4E-12 | 1.236936 | 1.165528 | 1.312718 |
| Immune_age_gap | E4_LIPOPROT | Inverse variance weighted | 25 | -0.07225 | 0.017714 | 4.53E-05 | 0.930297 | 0.898552 | 0.963164 |
| Immune_age_gap | G6_AD_WIDE | Inverse variance weighted | 8 | -0.03673 | 0.008493 | 1.53E-05 | 0.963936 | 0.948022 | 0.980117 |
| Metabolic_age_gap | RX_STATIN | Inverse variance weighted | 87 | 0.195489 | 0.024735 | 2.72E-15 | 1.215906 | 1.158364 | 1.276307 |
| Metabolic_age_gap | T2D | Inverse variance weighted | 92 | 0.135659 | 0.022415 | 1.43E-09 | 1.145292 | 1.096063 | 1.196731 |
| Metabolic_age_gap | KELA_DIAB_INSUL_EXMORE | Inverse variance weighted | 87 | 0.130886 | 0.020905 | 3.83E-10 | 1.139838 | 1.094078 | 1.187512 |
| Metabolic_age_gap | E4_METABOLIA | Inverse variance weighted | 19 | 0.393877 | 0.085355 | 3.94E-06 | 1.482718 | 1.254304 | 1.752728 |
| Metabolic_age_gap | E4_DM2NASCOMP | Inverse variance weighted | 91 | 0.137762 | 0.020165 | 8.39E-12 | 1.147702 | 1.103225 | 1.193972 |
| Metabolic_age_gap | T2D_WIDE | Inverse variance weighted | 62 | 0.134072 | 0.027315 | 9.18E-07 | 1.143476 | 1.083866 | 1.206363 |
| Metabolic_age_gap | E4_LIPOPROT | Inverse variance weighted | 25 | 0.288242 | 0.051362 | 2E-08 | 1.33408 | 1.206318 | 1.475372 |
| Metabolic_age_gap | E4_HYPERCHOL | Inverse variance weighted | 21 | 0.285819 | 0.049144 | 6.03E-09 | 1.330852 | 1.208642 | 1.465419 |
| Metabolic_age_gap | G6_AD_WIDE | Inverse variance weighted | 8 | 0.066829 | 0.012495 | 8.88E-08 | 1.069112 | 1.043247 | 1.095619 |
| Metabolic_age_gap | E4_HYPERLIPNAS | Inverse variance weighted | 9 | 0.33123 | 0.057931 | 1.08E-08 | 1.39268 | 1.243194 | 1.560139 |
| Musculoskeletal_age_gap | M13_RHEUMA | Inverse variance weighted | 18 | 0.032774 | 0.007718 | 2.17E-05 | 1.033317 | 1.017804 | 1.049067 |
| Pulmonary_age_gap | J10_LOWCHRON | Inverse variance weighted | 23 | 0.157539 | 0.034114 | 3.87E-06 | 1.170627 | 1.094914 | 1.251575 |
| Pulmonary_age_gap | J10_ASTHMA_EXMORE | Inverse variance weighted | 28 | 0.147994 | 0.030992 | 1.79E-06 | 1.159506 | 1.09117 | 1.232122 |
| Pulmonary_age_gap | J10_ASTHMACOPDKELA | Inverse variance weighted | 34 | 0.134479 | 0.024261 | 2.97E-08 | 1.143941 | 1.090818 | 1.199651 |
| Pulmonary_age_gap | J10_ASTHMA_MAIN_EXMORE | Inverse variance weighted | 23 | 0.128606 | 0.01813 | 1.31E-12 | 1.137242 | 1.09754 | 1.17838 |

| | | | | | | | | | |
|---|---|---|---|---|---|---|---|---|---|
| Pulmonary_age_gap | ASTHMA_INFECTIONS | Inverse variance weighted | 13 | 0.101217 | 0.019429 | 1.89E-07 | 1.106517 | 1.065172 | 1.149467 |
| Pulmonary_age_gap | ASTHMA_ACUTE_RESPIRATORY_INFECTIONS | Inverse variance weighted | 9 | 0.128295 | 0.02248 | 1.15E-08 | 1.136889 | 1.087884 | 1.188101 |
| Pulmonary_age_gap | ALLERG_ASTHMA | Inverse variance weighted | 14 | 0.092517 | 0.015499 | 2.38E-09 | 1.096931 | 1.064111 | 1.130764 |
| Renal_age_gap | E4_OBESITY | Inverse variance weighted | 19 | 0.104542 | 0.018812 | 2.74E-08 | 1.110202 | 1.070012 | 1.151902 |
| Renal_age_gap | ABDOM_HERNIA | Inverse variance weighted | 11 | 0.080233 | 0.018786 | 1.95E-05 | 1.083539 | 1.044369 | 1.124178 |
| Renal_age_gap | E4_OBESITYCAL | Inverse variance weighted | 10 | 0.111586 | 0.014416 | 9.89E-15 | 1.11805 | 1.086902 | 1.150091 |
| Renal_age_gap | G6_AD_WIDE | Inverse variance weighted | 8 | -0.03673 | 0.008957 | 4.12E-05 | 0.963937 | 0.947162 | 0.981008 |
| Brain_age_gap | AD | Inverse variance weighted | 20 | 0.060638 | 0.014905 | 4.74E-05 | 1.062514 | 1.031923 | 1.094013 |

### eTable 5: The 59 phenotypes used in the PWAS analysis

| Type | Category | Phenotype | Field ID |
|---|---|---|---|
| numeric | cognitive_function | number_of_puzzles_correct_f21004 | 21004 |
| numeric | cognitive_function | number_of_symbol_digit_matches_made_correctly_f23324 | 23324 |
| numeric | cognitive_function | number_of_puzzles_correctly_solved_f6373 | 6373 |
| numeric | cognitive_function | duration_to_complete_numeric_path_trail_1_f6348 | 6348 |
| numeric | cognitive_function | duration_to_complete_alphanumeric_path_trail_2_f6350 | 6350 |
| numeric | cognitive_function | fluid_intelligence_score_f20016 | 20016 |
| numeric | cognitive_function | maximum_digits_remembered_correctly_f4282 | 4282 |
| numeric | cognitive_function | mean_time_to_correctly_identify_matches_f20023 | 20023 |
| numeric | Lifestyle_and_environment | coffee_intake_f1498 | 1498 |
| numeric | Lifestyle_and_environment | fresh_fruit_intake_f1309 | 1309 |
| ordinal | Lifestyle_and_environment | oily_fish_intake_f1329 | 1329 |
| ordinal | Lifestyle_and_environment | processed_meat_intake_f1349 | 1349 |
| ordinal | Lifestyle_and_environment | cheese_intake_f1408 | 1408 |
| ordinal | Lifestyle_and_environment | salt_added_to_food_f1478 | 1478 |
| numeric | Lifestyle_and_environment | tea_intake_f1488 | 1488 |
| ordinal | Lifestyle_and_environment | smoking_status_f20116 | 20116 |
| ordinal | Lifestyle_and_environment | weekly_usage_of_mobile_phone_in_last_3_months_f1120 | 1120 |
| ordinal | Lifestyle_and_environment | sleeplessness_insomnia_f1200 | 1200 |
| ordinal | Lifestyle_and_environment | nap_during_day_f1190 | 1190 |
| numeric | Lifestyle_and_environment | sleep_duration_f1160 | 1160 |
| ordinal | Lifestyle_and_environment | plays_computer_games_f2237 | 2237 |
| numeric | Lifestyle_and_environment | time_spend_outdoors_in_summer_f1050 | 1050 |
| binary | Lifestyle_and_environment | alcohol_drinker_status_f20117 | 20117 |
| numeric | Lifestyle_and_environment | time_spent_using_computer_f1080 | 1080 |
| categoric | Mental_health | bipolar_and_major_depression_status_f20126 | 20126 |
| numeric | Mental_health | neuroticism_score_f20127 | 20127 |
| binary | Mental_health | probable_recurrent_major_depression_moderate_f20124 | 20124 |
| binary | Mental_health | probable_recurrent_major_depression_severe_f20125 | 20125 |
| binary | Mental_health | mood_swings_f1920 | 1920 |
| binary | Mental_health | miserableness_f1930 | 1930 |
| binary | Mental_health | irritability_f1940 | 1940 |
| binary | Mental_health | sensitivity_hurt_feelings_f1950 | 1950 |
| binary | Mental_health | fedup_feelings_f1960 | 1960 |
| binary | Mental_health | nervous_feelings_f1970 | 1970 |
| binary | Mental_health | worrier_anxious_feelings_f1980 | 1980 |
| binary | Mental_health | tense_highly_strung_f1990 | 1990 |
| binary | Mental_health | worry_too_long_after_embarrassment_f2000 | 2000 |
| binary | Mental_health | suffer_from_nerves_f2010 | 2010 |
| binary | Mental_health | guilty_feelings_f2030 | 2030 |
| binary | Mental_health | risk_taking_f2040 | 2040 |
| ordinal | Mental_health | workjob_satisfaction_f4537 | 4537 |
| ordinal | Mental_health | health_satisfaction_f4548 | 4548 |
| ordinal | Mental_health | family_relationship_satisfaction_f4559 | 4559 |
| ordinal | Mental_health | friendships_satisfaction_f4570 | 4570 |
| ordinal | Mental_health | financial_situation_satisfaction_f4581 | 4581 |
| ordinal | Mental_health | frequency_of_depressed_mood_in_last_2_weeks_f2050 | 2050 |
| ordinal | Mental_health | frequency_of_unenthusiasm_disinterest_in_last_2_weeks_f2060 | 2060 |
| ordinal | Mental_health | frequency_of_tenseness_restlessness_in_last_2_weeks_f2070 | 2070 |
| ordinal | Mental_health | frequency_of_tiredness_lethargy_in_last_2_weeks_f2080 | 2080 |
| binary | Mental_health | seen_doctor_gp_for_nerves_anxiety_tension_or_depression_f2090 | 2090 |
| binary | Mental_health | seen_a_psychiatrist_for_nerves_anxiety_tension_or_depression_f2100 | 2100 |
| binary | Mental_health | ever_depressed_for_a_whole_week_f4598 | 4598 |
| numeric | Mental_health | longest_period_of_depression_f4609 | 4609 |
| numeric | Mental_health | number_of_depression_episodes_f4620 | 4620 |
| binary | Mental_health | ever_unenthusiasticdisinterested_for_a_whole_week_f4631 | 4631 |
| binary | Mental_health | ever_manichyper_for_2_days_f4642 | 4642 |
| binary | Mental_health | ever_highly_irritableargumentative_for_2_days_f4653 | 4653 |
| numeric | Mental_health | longest_period_of_unenthusiasm_disinterest_f5375 | 5375 |
| numeric | Mental_health | number_of_unenthusiasticdisinterested_episodes_f5386 | 5386 |

**eFile1-9: Files contain large-size table results**

Due to the large size of these files, we share the data via Google Docs during peer review: https://drive.google.com/drive/folders/18IZxaL85VETSfvutiP9rJ0Y_xLFFdCNn?usp=sharing.
